# Genome-wide association study on longitudinal and cross-sectional traits of child health and development in a Japanese population

**DOI:** 10.1101/2025.02.18.25322458

**Authors:** Natsuhiko Kumasaka, Hidetoshi Mezawa, Tatsuhiko Naito, Kei Sakamoto, Kazuhisa Akiba, Atsushi Hattori, Mikko Baur, Tomohiko Isobe, Miori Sato, Mayako Saito-Abe, Aya Yoshikawa, Limin Yang, Daisuke Harama, Yumiko Miyaji, Minaho Nishizato, Makiko Sekiyama, Miyuki Iwai-Shimada, Yayoi Kobayashi, Mai Takagi, Nozomi Tatsuta, Yu Taniguchi, Kenji Matsumoto, Kazuhiko Nakabayashi, Masayo Kagami, Emiko Noguchi, Akira Hata, Kenichiro Hata, Atsushi Tajima, Shi Mingyang, Masaru Koido, Yoichiro Kamatani, Go Sato, Koichi Matsuda, BioBank Japan, Yosuke Kawai, Katsushi Tokunaga, National Center Biobank Network, Nicolas Fragoso-Bargas, Marc Vaudel, Stefan Johansson, Reiko Kishi, Chiharu Ota, Koichi Hashimoto, Chisato Mori, Shuichi Ito, Ryoji Shinohara, Hidekuni Inadera, Takeo Nakayama, Ryo Kawasaki, Yasuhiro Takeshima, Seiji Kageyama, Akemi Morita, Narufumi Suganuma, Shouichi Ohga, Takahiko Katoh, Michihiro Kamijima, Kiwako Yamamoto-Hanada, Akihiro Umezawa, Maki Fukami, Shin Yamazaki, Yukinori Okada, Shoji F. Nakayama, Yukihiro Ohya

**Affiliations:** Medical Support Center for the Japan Environment and Children’s Study, National Center for Child Health and Development, Tokyo, Japan; Human Genome Center, Institute of Medical Science, The University of Tokyo, Tokyo, Japan; Mohn Center for Diabetes Precision Medicine, Department of Clinical Science, University of Bergen, Bergen, Norway; Programme Office for the Japan Environment and Children’s Study, National Institute for Environmental Studies, Tsukuba, Japan; Department of Allergy and Clinical Immunology, National Center for Child Health and Development Research Institute, Tokyo, Japan; Department of Maternal-Fetal Biology, National Center for Child Health and Development Research Institute, Tokyo, Japan; Department of Molecular Endocrinology, National Center for Child Health and Development Research Institute, Tokyo, Japan; Institute of Medicine, University of Tsukuba, Tsukuba, Japan; Graduate School of Medicine, Chiba University, Chiba, Japan / Chiba Foundation for Health Promotion & Disease Prevention, Chiba, Japan; Graduate School of Medicine, Gunma University, Maebashi, Japan; Graduate School of Advanced Preventive Medical Sciences, Kanazawa University, Kanazawa, Japan; Graduate School of Frontier Sciences, The University of Tokyo, Tokyo, Japan; Graduate School of Medicine, The University of Tokyo, Tokyo, Japan; Genome Medical Science Project, National Center for Global Health and Medicine Research Institute, Tokyo, Japan; Department of Pediatrics and Adolescent Medicine, Haukeland University Hospital, Bergen, Norway; Center for Environmental and Health Sciences, Hokkaido University, Sapporo, Japan; Graduate School of Medicine, Tohoku University, Sendai, Japan; School of Medicine, Fukushima Medical University, Fukushima, Japan; School of Medicine, Chiba University, Chiba, Japan; School of Medicine, Yokohama City University, Yokohama, Japan; Center for Birth Cohort Studies, University of Yamanashi, Chuo, Japan; School of Medicine, University of Toyama, Toyama, Japan; Graduate School of Medicine, Kyoto University, Kyoto, Japan; Graduate School of Medicine, University of Osaka, Suita, Japan; Pediatrics, Hyogo Medical University, Nishinomiya, Japan; Faculty of Medicine, Tottori University, Yonago, Japan; Kochi Medical School, Kochi University, Nankoku, Japan; Graduate School of Medical Sciences, Kyushu University, Fukuoka, Japan; Faculty of Life Sciences, Kumamoto University, Kumamoto, Japan; Graduate School of Medical Sciences, Nagoya City University, Nagoya, Japan; Medical Research Center for Japan Environment and Children’s Study, National Center for Child Health and Development Research Institute, Tokyo, Japan

## Abstract

Understanding the influence of both genetics and environment on human health, especially early in life, is essential for shaping long-term health. Here, we utilize a nationwide prospective birth cohort, the Japan Environment and Children’s Study (JECS), to conduct a large-scale population-based genetic study using biannual questionnaire surveys and biological and physical measurements collected from both parents and their children up to the age of 4, since the participant mothers were pregnant. Analyses of genome-wide genotyping for 80,639 child participants with parental consent and sufficient DNA from cord blood samples represent the genetic diversity of the general population in Japan. Systematic genome-wide association studies of 1,255 child health and developmental traits (including, e.g., food allergy, anthropometry measurements or ASQ-3 developmental screening) and parental environmental exposure traits (including, e.g., mercury or PFAS exposure) identify 6,073 common genomic loci (2,332 of which passed the phenome-wide significance threshold *P* = 4. 0 × 10^−11^), of which 18-20% of loci represent novel associations not previously reported, including loci with potential therapeutic relevance. Additional longitudinal GWAS of BMI, using Gaussian process regression, identified 158 independent loci, of which 41 were not identified by age-stratified cross-sectional GWAS. These associations can also be used to predict an individual’s growth trajectory based on their genetic background. In addition, proxy-based genetic correlation analysis identified composite genetic signals linking maternal environmental exposures during pregnancy and child traits after birth. Together with studies of genetic risk factors and environmental exposures, across time and multiple outcomes, these demonstrate the uniqueness and value of the JECS data.

## Introduction

A birth cohort study is the ideal genetic research platform for studying genetic variations in human health and development over the life course^1^. While the number of cases is not as large as in a disease case-control study, it is possible to study a wide range of phenotypes, especially in infant and childhood participants, or environmental exposures as intermediate phenotypes that may account for missing heritability in complex diseases^2^. This platform, particularly once parental genotypes are available, will enable partitioning of genetic effects on birth phenotypes into maternal and fetal components^3^. Furthermore, a biobank alliance between a birth cohort and a case-control study allows us to project the adult disease risks in early life outcomes for early intervention or a potential route to new therapies and diagnostics^4–6^.

The Japan Environment and Children’s Study (JECS) is a large-scale birth cohort study funded by the Ministry of the Environment, Japan, to evaluate the effects of environmental chemicals on children’s health and development^7^. Over 100,000 pregnant women were enrolled at 15 geographically different Regional Centres across Japan, representing the comprehensive genetic diversity of the Japanese population. Detailed data from questionnaires, biological and physical measurements have been collected from both parents and their children since the participant mothers were pregnant, with additional surveys every six months being conducted in, on average, 80% of the child participants until the age of 4. Furthermore, JECS is actively engaged in the collection of samples and data pertaining to the pervasive exposure to environmental pollutants, including heavy metals^8,9^ and per- and polyfluoroalkyl substances (PFAS)^10,11^.

Genome-wide association studies (GWAS) are powerful tools for uncovering genetic factors behind complex traits and diseases^12^, enabling better diagnosis^13^, prevention^14^, and treatment^15^. As of 2024, over 691,532 associations across 36,643 traits have been cataloged^12^. However, despite the broad success of GWAS, the genetic basis of many phenotypes remains incompletely characterized, especially for less-studied traits and early-life outcomes. We here present the results of GWAS investigating a wide range of traits related to child health and development, as well as parental environmental exposures. Subsequently, cross-trait LD score regression (LDSC) analysis was conducted for a cross-sectional analysis of child phenotypes, such as various allergic traits. The analysis suggests pleiotropic associations in similar traits and highlights an East Asian-specific variant of potential therapeutic relevance. Furthermore, in a novel and distinctive attempt, we elucidate age-specific genetic associations with the anthropometric measurement BMI using a Gaussian process regression as an example of a longitudinal GWAS. This analysis uncovers novel genetic associations in a Japanese population and creates personalized growth curves, which could significantly improve the assessment and management of growth disorders and obesity. Lastly, cross-trait LDSC analysis revealed proxy-based genetic correlations for some trait pairs (e.g. maternal IgE and childhood allergy) between maternal exposures and offspring traits. The findings of these analyses, including GWAS summary statistics for 1,255 traits are available via the JECS PheWeb (https://jecs-pheweb.ncchd.go.jp/).

### The JECS genetic study of 100K children born in Japan

JECS is a nationwide government-funded birth cohort study in Japan. The study design of JECS has been published elsewhere^7,16^. Briefly, 103,099 pregnancies were enrolled at 15 Regional Centres across Japan through Co-operating Health Care Providers and/or local government offices between 2011 and 2014 (**Fig. 1a**, **Methods**). The previous publication reported that the children in JECS covered approximately 45% of the total live births in the study area^16^. Of these, 80,639 child participants with parental consent and sufficient DNA from the cord blood samples have been genotyped using the Illumina Asian Screening Array (**Methods**). The result yielded 617,557 genome-wide SNP variants after fixing strand orientations and matching alleles to the reference sequence (GRCh38) (**Methods**). A total of 109 samples with low DNA quality, as defined by a sample call rate less than 0.98 (**Fig. 1b**), were excluded from further analysis.

**Figure 1.**
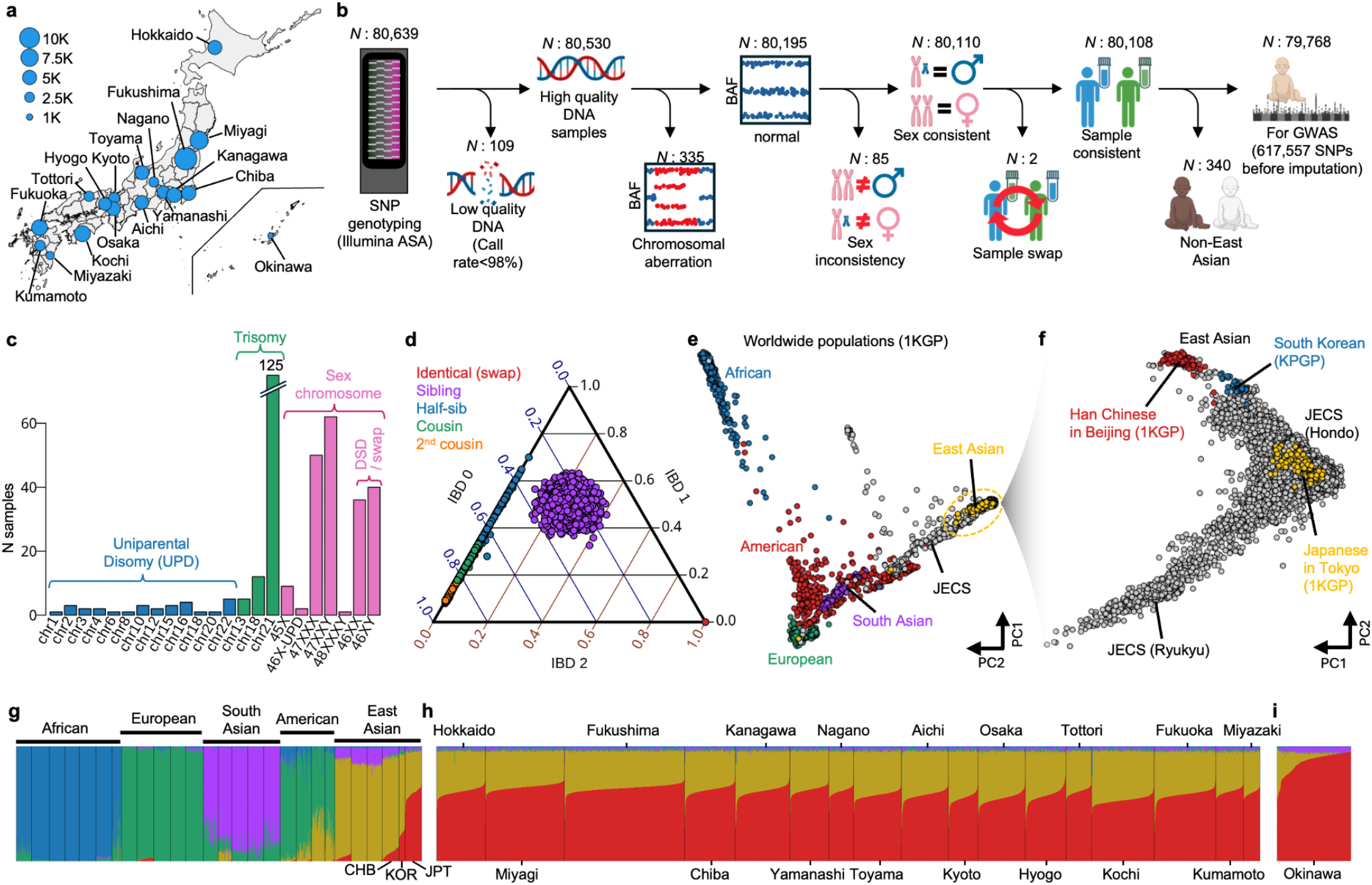
Overview of the JECS genetic study. **a.** Geographic distribution and demographic statistics of the 80,639 participants involved in the JECS genetic study. Dot size on the map indicates the number of participants in each region. **b.** Sample quality control flow for GWAS. Samples were selected by call rate (CR), B allele frequency (BAF) distributions, inferred sex based on genotype data and genotype principal components (**Methods**). **c.** Number of chromosomal aberrations for each chromosome. **d.** Ternary plot shows proportions of identity-by-descent (IBD) segments for each pair of participating children. **e.** The first two genotype principal components of JECS samples with worldwide populations obtained from the 1000 Genomes Project (1KGP) and Korean Personal Genome Project (KPGP). Dots are colored by different populations (gray: JECS; blue: African; green: European; red: American; purple: South Asian; yellow: East Asian). **f.** The first two principal components of East Asian populations (gray: JECS; red: Han Chinese in Beijing; blue: South Korean; yellow: Japanese in Tokyo). **g-i.** The stacked bar plots show the result of ADMIXTURE analysis for worldwide populations in 1KGP with KPGP (*N*=2,587) and JECS participants including non-East Asian populations (*N*=80,108). The result is split into three panels, showing the populations from 1KGP with KPGP (panel g), participants enrolled in mainland Japan (Hondo; *N*=79,353; panel h) and those enrolled in Okinawa Regional Centre (Ryukyu Islands; *N*=755; panel i). The y-scales differ between the panels: the panel g and i are 10 times wider than panel h, to clearly show the ancestry differences. JPT: Japanese in Tokyo, Japan; CHB: Han Chinese in Beijing, China; and KOR: South Korean (KPGP).

Subsequently, chromosomal aberrations were identified from the Illumina B allele frequency of signal intensity data (**Methods**) to yield a total of 335 participants exhibiting one or more autosomes that are uniparental disomy or trisomy (**Fig. 1b,c**). Additionally, some participants exhibited loss of the X/Y chromosome, as well as duplication of the X chromosome (**Fig. 1c**). Following the selection of normal karyotypes, we examined discrepancies between the inferred sex from the genotype and that from the questionnaire, resulting in the removal of 85 samples (**Fig. 1b**). Given that we collected 100K samples from a general Japanese population, it is plausible that some of the sex-inconsistent samples may represent disorders of sexual development^17^ (DSDs) rather than sample swapping. Note that the potential sample swapping was not enriched at certain Regional Centres (the minimum Fisher’s exact test *P*-value was 0.60 at a Regional Centre after Bonferroni correction).

Following the removal of participants containing chromosomal aberrations, MoChA^18–20^ was additionally used to identify participants with mosaic chromosomal alterations (mCA). This analysis revealed a prevalence of 1.9% for autosomal mCA and 1.1% and 1.6% for mosaic loss of the X and Y chromosomes, respectively (**Methods**; **Extended Data Fig. 1a-c**). For comparison, the corresponding frequencies in an adult population obtained from the BioBank Japan (BBJ), combining the first and second BBJ cohorts, were, on average, 11.3%, 37.0% and 25.3% (**Methods**). Note that we did not remove any participants carrying mCA because the mCA events were relatively small and the cell fractions were low (**Extended Data Fig. 1b**).

We then employed KING^21^ to examine the relatedness of our samples (**Methods**). The analysis revealed a high degree of relatedness in the data, with 4,131 pairs of half- and full siblings, and 9,628 pairs of cousins (**Fig. 1d**). We also found the number of related samples tended to be lower in urban areas and higher in rural areas of Japan (**Extended Data Fig. 1d, e**). This discrepancy is likely due to the effective sample size in each area. Additionally, two samples were identified as genetically identical (**Fig. 1d**), which is unlikely to occur due to the sampling scheme of cord blood (**Methods**). Therefore, these samples were removed due to a potential sample swap (**Fig. 1b**).

Finally, a population stratification analysis was performed using principal component analysis^22^ and ADMIXTURE^23^ (**Methods**). Due to the recruitment criteria (at least one parent must be literate in Japanese), some participants may not be of Japanese origin. In fact, 340 non-East Asian children were found to have African, South American, South Asian and/or European ancestries to varying degrees (**Fig. 1e**, **Extended Data Fig. 1f**). Following the removal of the aforementioned samples, the first two principal components demonstrated the existence of genetic diversity from the mainland Japanese (Hondo) to the East Asian continental population (including Chinese individuals from Beijing and South Koreans) and the Japanese Ryukyu ancestry (**Fig. 1f**), which can be seen to reflect the dual structure of the Japanese population previously reported^24,25^. Notably, compared to the other East Asian populations (**Fig. 1g**), the JECS participants exhibited a higher proportion of the Japanese-specific ancestry (red component in **Fig. 1h,i**), which was particularly high among participants enrolled at the Okinawa Regional Centre (**Fig. 1i**). However, this increase is gradual rather than discrete; the ancestry proportions shift continuously along the geographical gradient from Hondo to Okinawa without forming a clearly distinct cluster (**Fig. 1f and i**). In other words, the Japanese population structure appears clinal, reflecting continuous genetic variation rather than sharp categorical boundaries between Ryukyu and Hondo. In fact, in previous large-scale GWAS conducted in BBJ, Japanese participants have commonly been analyzed as a single ancestry group, while subtle population structure was controlled by excluding non–East Asian outliers and including genetic principal components as covariates^26–29^.

A total of 79,768 samples have been determined to meet the quality control standards (**Fig. 1b**). It should be noted that JECS collected the cord blood sample during delivery, and only one sample was taken, even in cases of multiple births (**Methods**). While there were 502 twins and 4 triplets in the genotyped samples, one child was genotyped while the other other child / co-twin (or the other two children in a triplet) was not. In contrast, phenotypes were observed for each child independently, irrespective of whether they were twins or triplets, and the sample size was not negligible (1,014 twin and triplet children). Therefore, we assigned the same genotype data from one twin to the co-twin(s) when a multiple birth was recorded and the reported sex of the child matched the genotyped sample. This resulted in 80,278 children, including those from multiple births whose sexes are consistent with their genotypes, for downstream analysis. We acknowledge that this approach may introduce some degree of non-independence due to shared genetic and environmental factors among siblings, which was formally evaluated in the sensitivity analysis section below.

### GWAS identify common non-coding genetic variants as major genetic determinants of child health and development

Large-scale GWAS of more than 80K children were then conducted on 1,046 child traits selected from questionnaires, biological and physical measurements for children up to the age of four (see **Supplementary Table 1**). Some traits were not available for GWAS in their original form; therefore, transformed or combined traits were generated from the original phenotype data (**Methods**). To increase the power and reduce the type I error in GWAS, we filtered out traits with a case-control ratio of less than 1% for binary traits and a complexity (an index of how many identical values/answers in the trait) of less than 5% for quantitative traits (see **Methods** for more details). The test statistics are often inflated when samples from related individuals are analyzed in a GWAS. To address this issue, we employed REGENIE^30^ to account for the relatedness between samples (**Methods**).

In addition, proxy-based GWAS was performed on an extra 209 parental environmental exposure traits (see **Supplementary Table 1**). Here the directly-observed parental traits were regressed onto the child (proxy) genotypes to assess genetic associations, proposed as GWAS-by-proxy^31^, a viable approach given the observed correlation between child and parent genotypes (**Methods**). Although this approach was initially used in a case-control study^31^, this framework can be extended beyond case–control traits to quantitative traits of various environmental exposures, because the core idea is that an offspring genotype provides noisy information on the parental genotype, regardless of the scale of the phenotype. The expected effect estimate in this design is approximately half of the direct effect that would be obtained using parental genotypes, assuming no additional indirect effects or assortative mating. We emphasize that our primary aim was locus discovery and empirical prioritization of parental exposure-related traits, rather than precise effect-size estimation. Because the genetic contribution to many pregnancy exposure traits is unknown a priori, we applied GWAS-by-proxy as a systematic, hypothesis-generating screen to identify traits with detectable genetic signals for future exposure-genetics analyses.

For each trait, 17,454,650 common variants with a minor allele frequency (MAF) greater than 0.1% and an imputation quality score (*R*^2^) greater than 0.3 were tested (**Methods**). In total, 6,073 GWAS loci (including 821 secondary hits in the same region; **Methods**) were identified across 776 traits (**Supplementary Table 2; Extended Data Fig. 2a**), with the index variant of each locus exhibiting a *P*-value of less than the common genome-wide threshold (*P* = 0.5 × 10^−8^), of which 2,332 loci across 288 traits passed a more stringent threshold of *P*-values after Bonferroni correction for the 1,255 traits (phenome-wide significant *P* = 5. 0 × 10^−8^/1255 = 4. 0 × 10^−11^). We initially examined the number of GWAS loci identified for each trait as the sample size varied (**Fig. 2a**). As expected, the number of loci demonstrated an increase in conjunction with both sample size and the additive SNP heritability *h*^2^ (**Fig. 2a**, **Extended Data Fig. 2b**; **Methods**). It should be noted that over 90% of the traits included in the study were analyzed using sample sizes greater than 60,000 children, and 34% of the traits had additive SNP heritability greater than 5% (**Supplementary Table 1**). We then examined the data to identify highly heritable trait categories and found that the majority of biospecimen and anthropometry measurements identified at least one genetic association, such as height at 4 years old (**Fig. 2b**, **Extended Data Fig. 2c**).

**Figure 2.**
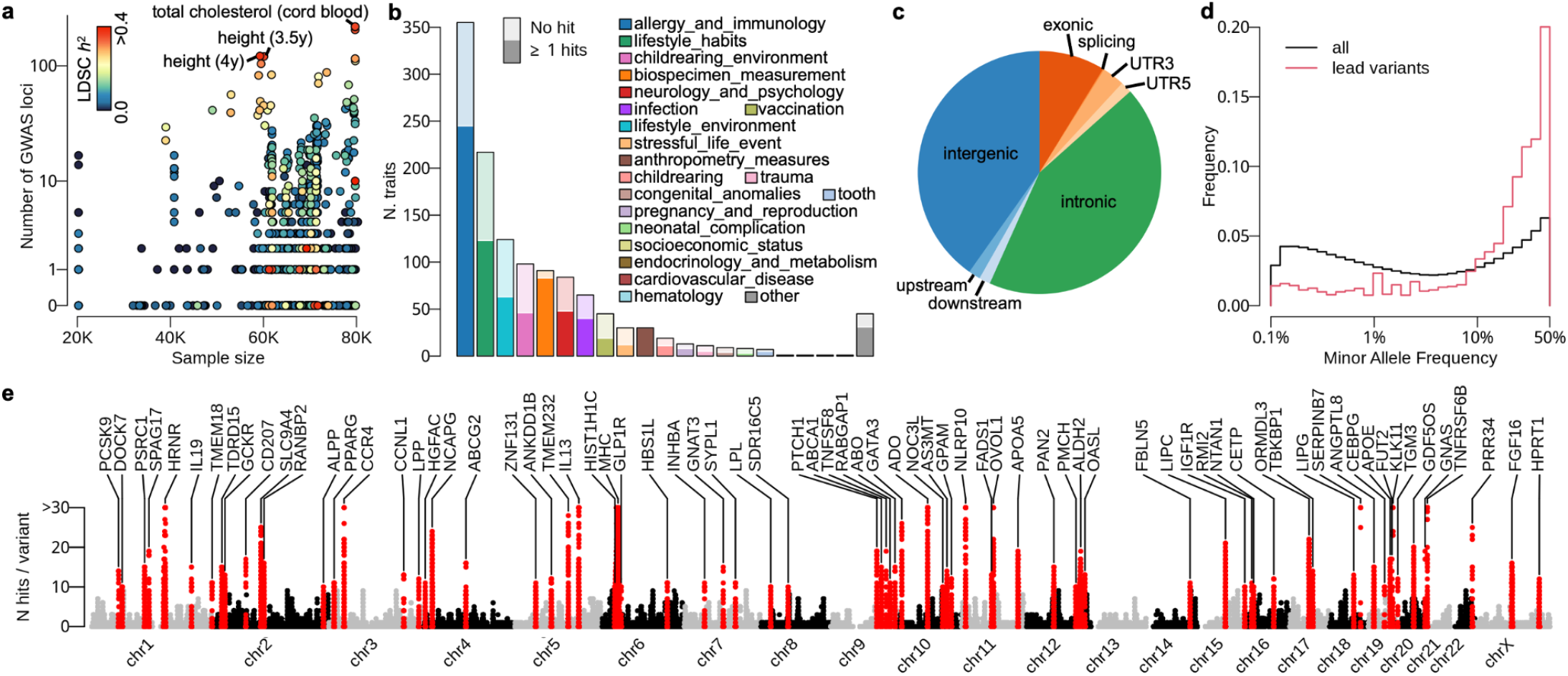
Characteristics of genetic loci and index variants identified by the JECS GWAS. **a.** Scatterplot shows the number of GWAS loci for each trait as a function of sample size. Each dot is colored by the additive SNP heritability estimation from the LD Score Regression (LDSC). **b.** The number of traits in different trait categories segregated by the existence of significant associations. Bars are split and darker colors show the number of traits with one or more genome-wide significant associations (*P* = 5. 0 × 10^−8^). **c.** Pie chart shows the proportion of variant locations relative to the Ensembl gene (version 113; GRCh38). **d.** Distribution of the minor allele frequency of the identified GWAS index variants against those of the entire tested variants. **e**. Summary of the genetic associations discovered through GWAS of 1,255 traits in JECS. Manhattan-like plot shows the number of GWAS hits (*P* = 5. 0 × 10^−8^) across 1,255 traits at each variant with the nearest gene annotation. Peaks are defined by variants shared across 10 or more traits.

According to the Ensembl gene annotation (version 113; GRCh38), only 9% of GWAS index variants were found in the coding sequences (**Fig. 2c**). Of these, 468 index variants (7.7% of all) potentially altered protein sequences (**Extended Data Fig. 2d**). Consequently, over 90% of variants were identified in non-coding regions (**Fig. 2c**), indicating that the discovered variants are likely to be involved in the regulatory cascades of neighboring genes, thereby influencing phenotypes. Within the range of variants tested (MAF > 0.1%), index variants were significantly enriched toward the common end of the allele frequency spectrum (MAF>10%) relative to the background distribution (**Fig. 2d**; Kolmogorov–Smirnov test *P* < 10^−16^), consistent with greater statistical power and imputation accuracy for common variants.

While common complex traits are highly polygenic, the number of variants is finite, which results in a single variant affecting multiple traits, a phenomenon known as pleiotropy. The landscape of pleiotropy in European and Japanese populations has been intensively studied, and these regions are recognized as being under recent selection pressure^32^. Here we also defined the degree of pleiotropy as the number of significant associations per variant (**Fig. 2e**), and replicated various known regions under selection pressure^32^, including the major histocompatibility complex (MHC), ABO, ALDH2, and APOE loci^32^.

Overall, our discovered loci spanned approximately 13% of the entire genome when taking the 200 kb cis-window centered on each index variant. We used our imputed genotype data to calculate the LD *r*^2^ index between the index variants of our GWAS and the entire variants reported in the GWAS catalog (**Methods**), regardless of traits (**Extended Data Fig. 2e**). We identified 1,221 out of 6,073 index variants (approximately 20% of index variants) that showed weak LD (*r*^2^ < 0.1), representing potentially novel genetic loci or distinct causal variants at known loci in a Japanese population. Alternatively, some of these loci may appear ’novel’ simply because the corresponding phenotypes have not yet been examined in GWAS that mostly focus on adult phenotypes. Note that this number doesn’t change much (1,167 or 1,123 variants) even if we set the cis-window to 500 kb or 1 Mb, respectively. We also repeated the analysis using the genotype data from the three major populations (AFR, EUR and EAS) in the 1000 Genomes Project (1KGP), and identified 1,674, 1,613 and 1,112 variants that showed weak LD (*r*^2^ < 0.1) in 1 Mb cis-window. This suggests that using a population-matched LD index is the most conservative approach and best tags previously reported GWAS hits. We note that all the tagged GWAS hits (*r*^2^ < 0.1) are also reported in **Supplementary Table 2**.

### HLA imputation contributes to improved biological insights into allergy and immunological traits

The allele-level HLA analysis is particularly informative for immune-related and allergy traits, as well as for interpreting associations in the major histocompatibility complex (MHC) region. We performed HLA imputation using DEEP*HLA^33^ and retrieved 2,103 classical HLA alleles and amino acid variants in both classical and non-classical HLA genes. We discovered 177 independent associations for the standard GWAS threshold, of which 106 passed the phenome-wide threshold (**Supplementary Table 3**). Of 183 traits with significant lead signals in the MHC region (from 28.5 to 33.5 Mb on chromosome 6), 133 traits were linked to HLA alleles (**Supplementary Table 1**).

Associations with food allergy-related traits were predominantly driven by class II HLA genes, with *HLA-DRB1* most frequently emerging as the primary signal (**Fig. 3a**). This observation underscores the central role of class II-mediated antigen presentation to CD4+ T cells in food sensitization. In contrast, some other traits showed associations with both class I and II genes; for instance, *HLA-DRB1*, *HLA-DPB1*, and *HLA-B* were found to be independently associated with self-reported itchy rash, a proxy for eczema (**Fig. 3b**). This is in line with the established involvement of both CD4+ and CD8+ T-cell pathways in the pathogenesis of eczema or atopic dermatitis, where class I HLA-restricted effector cells contribute to cutaneous inflammation^34^.

**Figure 3.**
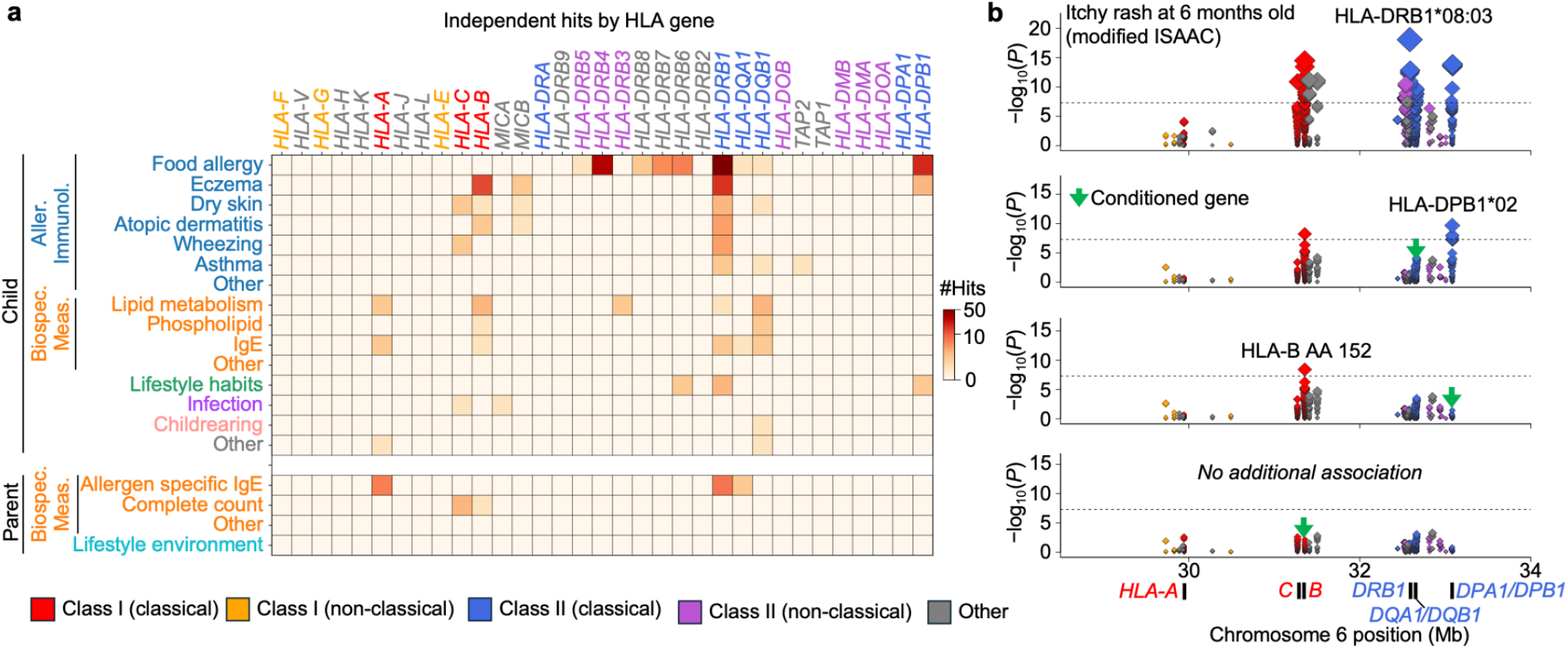
HLA imputation and fine-mapping in the MHC region. **a.** Heatmap showing the independent association counts of HLA genes and traits grouped by trait category and target. When the lead signals were *HLA-DRB2–9* variants in strong LD with *HLA-DRB1* variants (R2 > 0.9), *HLA-DRB1* was counted together. **b.** Four stacked panels showing the MHC regional association plots from the stepwise conditional regression analysis of itchy rash at 6 months old, based on the modified ISAAC questionnaire (c6m_0120001). The panels show nominal results, followed by results sequentially conditioned on *HLA-DRB1*, *HLA-DPB1*, and *HLA-B*. HLA gene labels in panel a and the variant markers in panel b are colored according to the HLA gene categories shown at the bottom.

### Sensitivity analyses addressing potential confounding from relatedness and population structure

Previous studies have shown that association test statistics can remain inflated in GWAS analyses, including those using REGENIE, when individuals sharing environmental influences are included. To assess the robustness of our results with respect to relatedness in our cohort, we performed GWAS in a subsample (*N*=70,751) excluding individuals related up to the third degree (**Methods**). In this analysis, we identified 5,126 GWAS loci (including secondary signals) across 737 traits at the conventional genome-wide significance threshold (*P* = 5. 0 × 10^−8^), of which 1,946 loci across 275 traits passed the phenome-wide significance threshold (*P* = 5. 0 × 10^−11^) (**Supplementary Table 2**). We observed high concordance with the main analysis: 82% of loci (4,193 loci) were identical (either the same index variants or proxies in LD *r*^2^ > 0.1) at the genome-wide threshold, and 97% (1,879 loci) were identical at the phenome-wide threshold (**Extended Data Fig. 3a**). The reduction in the number of detected loci was consistent with the 12% decrease in sample size (**Methods; Extended Data Fig. 3b–c**), suggesting that the observed differences are largely attributable to reduced power rather than confounding. We further compared attenuation ratios from LD score regression and observed a modest reduction (approximately 0.067 on average) after excluding related individuals (**Extended Data Fig. 3d**), indicating that shared familial effects may contribute slightly to the observed signal. However, this effect was small in magnitude, and the overall consistency of association results supports the robustness of our main findings.

Although Japan is often described as a genetically homogeneous, single-ethnicity population (**Fig. 1g–i**), lifestyle, childrearing, and social traits are strongly influenced by socio-cultural, economic, and policy factors^35,36^. Therefore, genetic associations identified in our study could, in principle, reflect genetic drift within specific ancestral subgroups. To address this possibility, we first compared genomic control lambda (*λ_GC_*) and LDSC intercepts between lifestyle-related and other traits and found no significant differences (**Extended Data Fig. 3e–f**). Next, we performed GWAS along the two major axes of ancestry variation (**Extended Data Fig. 3g**; **Methods**), capturing differences between (1) Japanese and other East Asian populations (e.g., Chinese and South Korean individuals) and (2) the Japanese Hondo and Ryukyu ancestries, as represented by the first two principal components (see **Fig. 1f**). We treated these axes as quantitative traits and conducted GWAS to identify variants associated with population structure and potential genetic drift (**Extended Data Fig. 3h–i**). Finally, we conducted genetic correlation analyses using the cross-trait LD score regression^37^ (**Methods**) and found no traits showing significant genetic correlation with these ancestry axes, even when stratifying traits into lifestyle and non-lifestyle categories (**Extended Data Fig. 3j**; **Supplementary Table 1**). These analyses suggest that the associations for lifestyle, childrearing, and social traits were not primarily driven by the major genetic ancestry axes captured by the first two principal components. However, these analyses do not rule out more local or non-genetic sources of confounding, including regional differences, socio-economic factors, healthcare access, family structure, correlated environments, or residual population structure. We therefore interpret loci for these traits as statistical genetic associations rather than evidence of direct biological effects.

### Genetic correlation analysis uncovers shared genetic architectures within/between trait categories

We then selected 585 child traits with positive SNP heritability estimates (additive SNP *h*^2^ > 0) and one or more genetic loci discovered through GWAS (**Supplementary Table 1**), and performed the genetic correlation analysis using cross-trait LD score regression^37^ (**Fig. 4**). We identified 8,057 significant genetic correlations (GCs) after multiple testing corrections (Benjamini-Hochberg *Q* < 0. 05; **Supplementary Table 4**). To define the multiple-testing burden in a broad and inclusive manner, we included all traits with positive SNP-heritability estimates (*h*^2^ *Z*-score > 0). This broader testing universe contains more trait pairs and therefore imposes a more stringent correction for declaring significant genetic correlations. Restricting the analysis to these more heritable traits (*h*^2^ *Z*-score > 2 or 4) produced stronger deviations from the null expectation in QQ plots (**Extended Data Fig. 4a**), which is better interpreted as increased power and a change in the tested trait set. Among these significant GCs, there were 7,672 GCs for which both traits had heritability *Z*-scores greater than 2, and 3,126 GCs for which both traits had heritability *Z*-scores greater than 4, consistent with the threshold recommended by LD score regression to ensure reliable genetic correlation estimates^37^. Heritability estimates, their standard errors, and corresponding *Z*-scores for all traits analyzed here are provided in both **Supplementary Table 1** and **4**.

**Figure 4.**
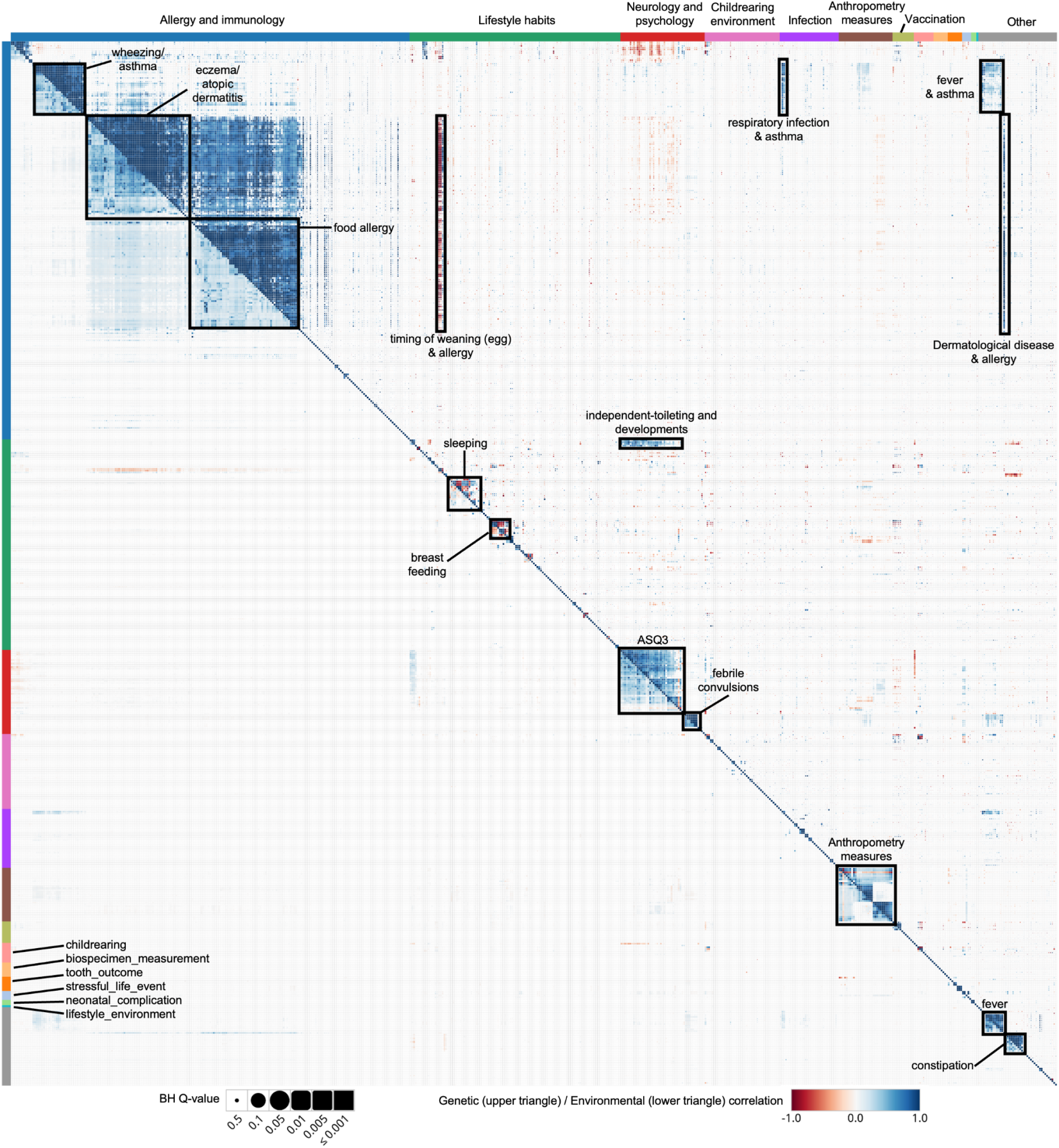
Genetic correlations (upper triangular matrix) and environmental correlations (lower triangular matrix) of 585 child traits estimated by the cross-trait LD score regression. Blue colors indicate positive correlations and red colors indicate negative correlations. The color bars at the top and left of the heatmap indicate trait categories.

Unsurprisingly, discovered GCs were significantly enriched within the same trait category (7,367 GCs; *OR* = 50. 7; *P* < 2. 2 × 10^−16^). The most frequent trait categories were “allergy and immunology” (6,312 GCs), “neurology and psychology” (472 GCs) and “anthropometry measures” (313 GCs) (**Fig. 4**). We discovered a modest number of genetic correlations between traits in different trait categories (690 GCs across trait categories below *Q* < 0. 05). For example, we identified GCs between wheezing/asthma traits and fever/infection traits (**Fig. 4**). There were also reasonable GCs between several developmental traits measured by the ASQ-3^38,39^ and independent-toileting traits (**Fig. 4**). Overall, there were no surprising pairs of traits that share genetic architectures. We also note that there was general agreement between environmental correlations (**Methods**) and genetic correlations of trait pairs (**Extended Data Fig. 4b**). The environmental correlations are not the phenotypic correlations and are computed from the output of LDSC software (see **Methods** for the details).

This analysis not only highlighted traits whose correlations were previously unknown, but also highlighted similar traits that exhibited both shared and unshared genetic loci. One such example is allergies. These often begin in early life, first manifesting as eczema in young children. The progression from eczema to the subsequent development of atopic dermatitis, food allergies, allergic rhinitis and asthma is known as the *allergic march*^40^. Indeed, a trait of itchy rash at one year old and a trait of egg allergy score at four years old showed a very high genetic correlation (*r_g_*= 0. 83; *P* = 5. 4 × 10^−30^), sharing multiple genetic loci involved in skin barrier function, such as the FLG/HRNR and SERPINB7 loci (**Extended Data Fig. 4c**). FLG loss-of-function variants are the most robust genetic risk factors for eczema and atopic dermatitis and also increase susceptibility to food allergy and childhood-onset asthma^41,42^, largely through impaired skin-barrier function and enhanced skin sensitization. Although functional variants of the FLG have been extensively studied in European and East Asian populations^43–45^, we identified a Japanese-specific nonsynonymous variant (rs540453626:G>C also known as p.S2889X) associated with both itchy rash (as a proxy for eczema) and egg allergy for the first time. The clade B serpin (SERPINB) gene cluster at 18q21.3 was previously reported by a GWAS of paediatric food allergy in a European population^46^. We have identified the SERPINB7 gene containing a novel East Asian-specific stop-gain mutation (rs142859678: C>T) that is significantly associated with both itchy rash and egg allergy (**Extended Data Fig. 4d**). It is known that the homozygous mutant of the SNP leads to an autosomal recessive inherited skin disorder, Nagashima-type palmoplantar keratosis^47^ (NPPK). This suggests that the mutant allele may also cause skin barrier defects and sensitization to various allergens, even in heterozygous individuals. Interestingly, gentamicin, an antibacterial ointment, has been proposed as a potential read-through therapy for NPPK^48–50^, although its clinical utility remains uncertain. While highly speculative, approaches that restore skin barrier function may have downstream effects on allergic sensitization in heterozygous carriers of stop-gain mutations.

Another example of genetically correlated traits is the food allergy traits between different foods (e.g., eggs and milk). The GWAS for egg allergy showed a strong association at the MHC locus (**Extended Data Fig. 4c**), consistent with previous studies showing that the MHC region represents the most robust and reproducible genetic risk locus for food allergy, although prior GWAS have been limited by relatively small sample sizes, often including only a few hundred to a few thousand cases^51^. In contrast, a trait of milk allergy at four years of age showed an association at the CTLA4 locus, but not in the MHC region (**Extended Data Fig. 4c**). Notably, the sample size in our study is more than 4-fold larger than that of previous GWAS of food allergy, including recent large-scale meta-analyses in European populations^52^ (*N*=14,234 children), thereby providing increased statistical power to detect additional loci beyond the MHC region. Recent studies have demonstrated that GWAS signals can be used to prioritize relevant cell types by integrating genetic associations with functional genomic and single-cell data^53,54^. In this context, the observed locus-specific differences raise the possibility that distinct immune cell types may contribute to the development of allergies to different foods. In line with this, scDRS^55^ analysis using PBMC data from the recent single-cell multi-omics data^56^ showed distinct enrichment patterns for egg and milk allergy across cell types. Specifically, significant enrichment in dendritic cells was observed only for egg allergy, and CD4+ T cells and monocytes showed a trend toward opposite directions of enrichment for egg and milk allergy (**Methods**; **Extended Data Fig. 4e,f**).

### GWAS of longitudinal traits reveal age-related genetic determinants

One of the potential benefits of using the birth cohort platform to study the genetics of child traits is that it allows us to track participants at multiple time points, which could help us to analyze any changes in their health and development over time^5^. One such trait that we are able to study in great detail is anthropometric traits, such as height or body mass index (BMI). As we collect the records every 6 months from birth and participants can enter them at their convenience, we have been able to retrieve the trajectory of BMI at monthly resolution (**Extended Data Fig. 5a**).

We initially performed standard GWAS of BMI at 11 age strata from birth to 4 years old with a proper adjustment by child age in months. We identified 454 loci with the common GWAS threshold (296 loci after phenome-wide Bonferroni correction at *P* = 4.5 × 10^−9^ in the 11 GWAS), comprising 197 independent genetic loci after LD pruning between index variants (*r*^2^ < 0.1 in our population) (**Fig. 5a**). By comparing with the BMI associations in the GWAS catalog, we identified 108 novel loci whose index variants are in weak LD (*r*^2^ < 0.1 with any of the BMI-associated variants previously reported. Because the overlap of loci is modest, we compared the effect sizes between the JECS and summary statistics from the GWAS results from up to 28,681 children in the Norwegian Mother, Father, and Child Cohort Study^5^ (MoBa) which were performed within the same age strata (**Methods**). Although the effect sizes are significantly correlated (Pearson *R*=0.76), most of the associations in the smaller MoBa study were below the GWAS threshold (**Extended Data Fig. 5b**). We then compared the genetic correlations between the GWAS at the 11 time points and the adult BMI GWAS obtained from BBJ^32^. We observed strong correlations within childhood GWAS, but modest correlations between childhood and adult GWAS, suggesting that the genetic determinants of childhood obesity may be less shared with adult obesity (**Extended Data Fig. 5c**).

**Figure 5.**
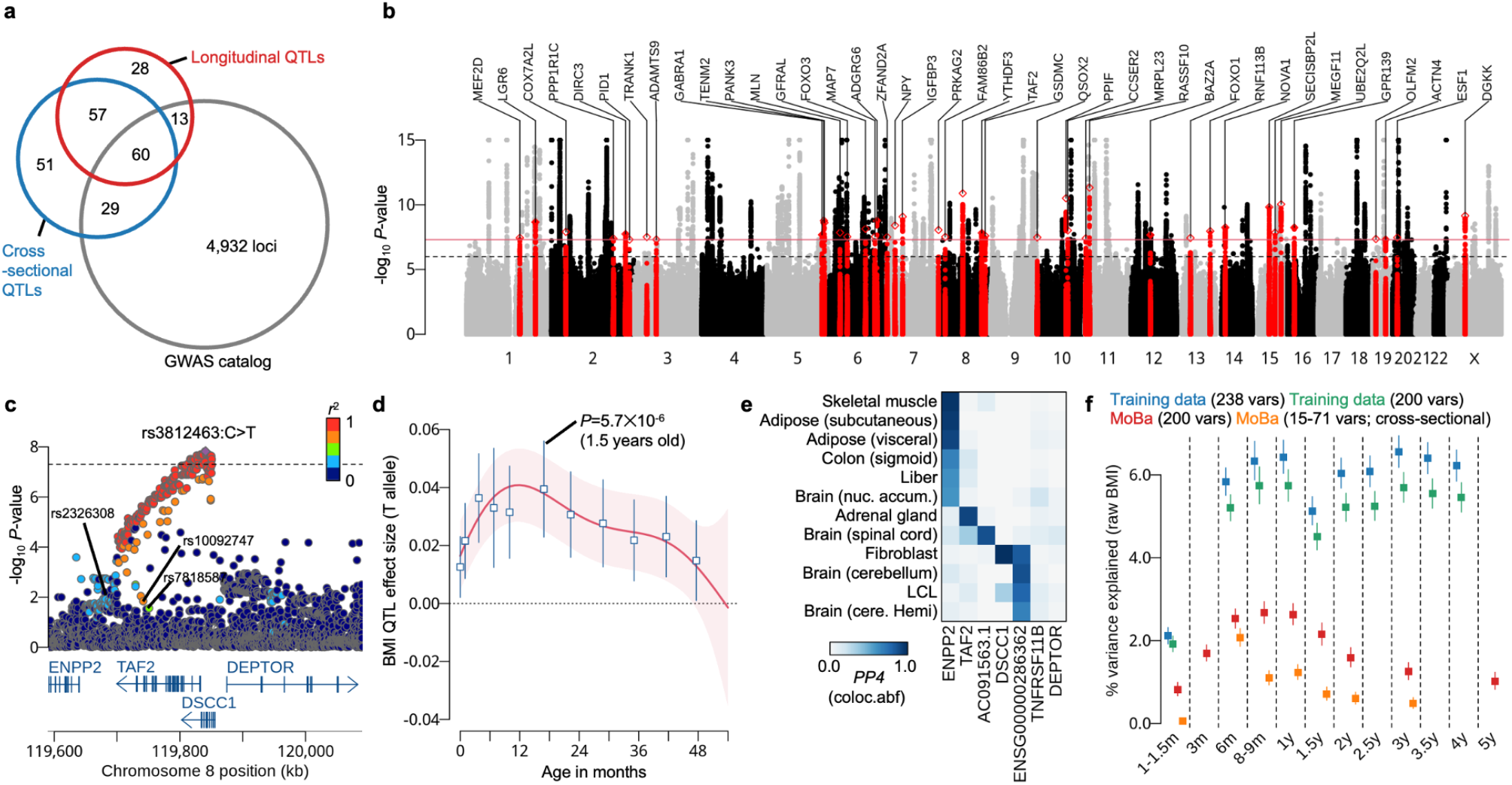
Genetic study of childhood BMI as a longitudinal trait in JECS. **a.** Venn diagram shows the genetic loci (lead variants whose LD *r*^2^ < 0.1) discovered through cross-sectional (blue; 197 loci) and longitudinal (red; 158 loci) GWAS of childhood BMI. Those loci are compared with BMI-related loci reported in the GWAS catalogue (gray). **b.** Manhattan plot shows the score test *P*-values obtained from the GWAS of the dynamic genetic effects of BMI from birth until 4 years old. The 41 independent loci only discovered in the longitudinal GWAS (panel a) are highlighted in red and the nearest genes are annotated. **c.** Locuszoom plot of the TAF2 locus shows the statistical significance of dynamic genetic effects. Points are colored by the LD index value ( ^2^) to the index variant rs3812463:C>T. The three height-associated SNPs (rs2326308:C>T, rs10092747:C>T, rs7818587:G>T) are also shown in this plot. **d.** Age-dependent genetic effect of the T allele relative to the C allele at rs3812463, estimated by Gaussian process regression. The red line and shaded area indicate the posterior mean and 95% credible interval, respectively. Cross-sectional QTL effect (computed from the raw BMI) estimates and their 95% confidence intervals across 11 age strata are overlaid as blue open squares and error bars. **e.** Heatmap shows the colocalization posterior probability (PP4) obtained from the coloc package in R. We used the summary statistics of GTEx eQTLs (V8) across 48 tissues in 1 Mb cis-window centered at each gene TSS. We found that 12 tissues showed PP4 greater than 0.5 in one of 5 different eQTL genes. We did not identify colocalized eQTL signals for TNFRSF11B or DEPTOR. **f.** Performance of the longitudinal and cross-sectional PGSs across age strata in JECS and MoBa. Points and error bars indicate the percentage of variance in raw BMI explained by each PGS and its 95% confidence interval, respectively. Blue points show the apparent in-sample performance of the dynamic PGS comprising 238 variants in the JECS discovery dataset (*N* = 79,743). Green points show the corresponding JECS estimates after restricting the PGS to the 200 variants available in MoBa. Red points show the external performance of this 200-variant dynamic PGS in MoBa (*N* = 65,455). Orange points show the performance in MoBa of age-specific PGSs constructed from the corresponding cross-sectional JECS GWAS, comprising 15–71 variants depending on the age stratum. MoBa data were used exclusively for external evaluation and were not used for locus discovery, variant selection, effect-size estimation or model tuning.

A recent review has emphasized that incorporating longitudinal trait trajectories into genetic epidemiology can provide deeper insight into the genetic architecture of trait development and disease progression^57^. Incorporating all data points simultaneously, we then performed a dynamic genetic mapping using the Gaussian process regression^58,59^ (**Fig. 5b**; **Methods**). This method explicitly models the genetic effect as a non-linear function of child age, regardless of whether the effect changes sign, changes magnitude, or appears only during part of the age range. It is more flexible than modeling only the constant mean shift of a data trajectory^60^ or a linear genetic effect along age^61^, or summarizing trajectories into a small set of derived phenotypes^62^ (e.g., mean or slope). Using this approach, we discovered an additional 41 independent genetic loci (*r*^2^ < 0. 1 with loci discovered through the cross-sectional GWAS of 11 age strata), of which 28 loci were novel and not in LD (*r*^2^ < 0. 1) with any BMI-related variant (**Fig. 5a,b**; **Supplementary Table 5**). As a proof of concept, we first examined the FTO locus, which is strongly linked to adult obesity. The FTO locus is known to be associated with infant BMI with an opposite effect direction to that in adults^63^, possibly reflecting differences in the relative proportions of brown and white adipose tissue between infants and adults^64^. We clearly showed that the QTL effect was reversed at around 3 years of age (**Extended Data Fig. 5d,e**), as previously reported^63^.

Then, we examined our novel discoveries, which were enriched for variants with significantly lower MAFs in a European population (1KGP EUR) (Kolmogorov–Smirnov test *P* = 0. 003). One of these novel associations was identified at the TAF2 locus (**Fig. 5c**). The strongest association was observed for a SNP (rs3812463:C>T) in an intron of the TAF2 gene, although 207 additional SNPs and INDELs near this SNP were in strong LD (*r*^2^ < 0. 8). The estimated effect size of the T allele over the C allele showed an age-dependent pattern, increasing in infants and decreasing in toddlers (**Fig. 5d**). This pattern was also consistent with the results from cross-sectional GWAS of raw BMI for 11 distinct age strata superimposed on the same plot (**Fig. 5d**). Effect size estimates from the cross-sectional GWAS were highly concordant with and without RINT (Pearson *r*=0.995; **Extended Data Fig. 5f,g**; **Methods**). Note that no genome-wide significant associations were observed in any cross-sectional GWAS, and the minimum *P*-value was 5. 7 × 10^−6^ at 1.5 years of age. We also found that there are several height-associated index variants (rs2326308:C>T, rs10092747:C>T, rs7818587:G>T) in this locus^65^ (**Fig. 5c**). However, the LD between the TAF2 index variant and those variants was modest (0. 25≤*r*^2^ ≤0. 64 in our population). Therefore, distinct association signals may be present, although population-specific LD patterns also allow for a shared causal variant. As there are several putative candidate genes in this locus, we checked the colocalization with eQTLs in GTEx (V8; **Data Availability**). We found that the locus is colocalized with 5 different eQTL genes in 12 different tissues (colocalization posterior probability *PP4*>0.5) (**Fig. 5e**). One of them was the ENPP2 eQTL in skeletal muscle tissue (*PP*4 = 0. 87), and the locuszoom plot was consistent with the GWAS locus (**Fig. 5c** and **Extended Data Fig. 5h**). ENPP2 encodes an enzyme that converts lysophosphatidyl -choline to lysophosphatidic acid (LPA) leading to growth factor-like responses such as stimulation of cell proliferation or chemotaxis^66^.

Together with the set of dynamic genetic effects estimated at each locus (*e.g.* TAF2 locus in **Fig. 5d**) combined with individual genotype data, a dynamic polygenic score (PGS) can be constructed along the child’s age (**Methods**). This information reveals the extent to which the genetic influence deviates from the population mean and the degree to which the genetic prediction aligns with the observed child’s growth (**Extended Data Fig. 5i-l**). In collaboration with MoBa, we carefully assessed the performance of the dynamic PGS model, comprising 238 lead variants discovered through the cross-sectional and longitudinal GWAS (**Methods**). First, we trained the dynamic PGS model using all our samples with BMI observations at least once in JECS (*N* = 79,743). We evaluated the model’s apparent in-sample performance in each age stratum of the same JECS discovery data (**Methods**). This demonstrated that the average variance in raw BMI explained by the PGS was 5.7% (ranging from 2.1% to 6.5% across the age range) in JECS (**Fig. 5f**), which should not be interpreted as an independent out-of-sample estimate of prediction accuracy. When we applied the model to an independent cohort dataset from MoBa (*N*=65,455; Norwegian children of Northern European descent), we found that the PGS prediction based on 200 variants available in MoBa (**Methods**), on average, explained 1.8% of the variance (ranging from 0.8% to 2.7% across the age range) in this European population. Notably, the PGS model still explained 1.0% (*P* = 6. 3 × 10^−81^) of the variance at age 5 in MoBa (outside the range of the trained model) (**Fig. 5f**). Note that MoBa data were used only for external evaluation; no locus discovery, variant selection, effect-size estimation, or model tuning was performed using the MoBa data. To benchmark the longitudinal approach against conventional age-specific GWAS, we additionally constructed PGSs from the cross-sectional GWAS performed at each age stratum (**Methods**). In MoBa validation, these cross-sectional PGSs explained only an average of 0.7% of BMI variance across ages (range, 0.06–2.1%) (**Fig. 5f**). We also recalculated the JECS in-sample PGS using the same 200 variants available in MoBa; excluding the 38 unavailable variants reduced the variance explained in JECS by approximately 10% (**Fig. 5f**). In all variance-explained analyses, we used only the genotype-weighted PGS and did not include the age-dependent non-genetic population-average trajectory which was specifically used for visualization in **Extended Data Fig. 5i–l**. The constructed dynamic PGS model with the full sample data is available on our GitHub page (**Code availability**).

### Sensitivity analyses of longitudinal GWAS addressing stability and reproducibility

First, to show the statistical calibration of the longitudinal GWAS model using the GP regression, we examined the score test *P*-values on the QQ-plot (**Extended Data Fig. 5m**) and computed the attenuation ratio of LDSC regression (*Ratio* = 0. 09), which was sufficiently low^67–71^, despite the high genomic control lambda (λ*_GC_* = 1. 58) (**Methods**). Next, we simulated genotypes with a MAF ranging from 0.001 to 0.5 while preserving relatedness among relatives up to the third degree (**Methods**) and confirmed that the *P*-values follow the uniform distribution (**Extended Data Fig. 5m**). We also performed a permutation test using the real genotype data while keeping the family member structures (**Methods**) and demonstrated that the *P*-values follow a uniform distribution (**Extended Data Fig. 5m**).

To further assess the robustness and interpretability of the findings, we compared the GP-based approach with two alternative analyses: a naive meta-analysis across 11 age strata (**Methods**) and a conventional age-by-genotype interaction model including linear and quadratic age terms (**Methods**). The meta-analysis across 11 age strata showed substantial inflation of the test statistics under both the null and alternative hypotheses (**Extended Data Fig. 5n**). Under the null, the genomic control factor reached λ*_GC_* = 2. 7, suggesting that failure to account adequately for within-individual correlation can result in poorly calibrated and potentially irreproducible findings.

We next evaluated the quadratic age-by-genotype interaction model, hereafter referred to as the quadratic polynomial mixed-effect model (QPMM), for the 63,640 variants identified by GP-based screening at *P* < 10⁻⁴ (**Methods**). Because genome-wide fitting of this mixed-effects model was computationally prohibitive, the QPMM was used as a targeted sensitivity analysis rather than as an independent genome-wide discovery scan. Its purpose was therefore to benchmark the GP results against a more conventional mixed-effects framework, rather than to replace the primary GP-based analysis. An analytical assessment further indicated that restricting the QPMM analysis to variants with GP *P* < 10⁻⁴ was unlikely to miss genome-wide significant QPMM associations (**Methods**). The GP and QPMM *P*-values showed strong concordance (**Extended Data Fig. 5o**), with a Pearson correlation of 0.95 on the log scale. Nevertheless, significant variants were more frequently detected by the GP approach: 6,656 variants reached genome-wide significance (P < 5 × 10⁻⁸) in both analyses, 4,782 did so only in the GP analysis, while 423 did so only in the QPMM (**Extended Data Fig. 5o**). After selecting the lead variant for each locus, 100 of the 106 loci identified by this model (94%) had already been detected by the GP-based longitudinal GWAS (**Extended Data Fig. 5p**), and 104 of the 106 loci had been detected by either the GP-based longitudinal GWAS or the cross-sectional GWAS. The QPMM can be interpreted as a restricted special case of the GP framework, in which the kernel matrix is limited to a second-degree polynomial and the covariance matrix of the latent random variables at the inducing points is fixed to the identity matrix (**Methods**). It is therefore expected to be most sensitive to associations whose age-dependent effects follow an approximately quadratic pattern (**Extended Data Fig. 5q,r**).

Finally, we generated synthetic longitudinal BMI data using the estimated population mean and variance structure, incorporating measured covariates, including simulated principal components and sex, as well as unobserved donor-specific variation modeled as a Gaussian process (**Methods**). We then added QTL effects for the 238 identified variants (**Supplementary Table 5**) by multiplying the posterior mean effect trajectory of each variant by the corresponding simulated genotype dosage for each individual, thereby generating data under a realistic alternative model (**Methods**). We applied the same analytical workflow used for the real data to a set of 1,000 variants, of which 238 had simulated QTL effects and 762 represented null variants. QQ plots under both the null and alternative models demonstrated appropriate statistical calibration and high sensitivity to the simulated QTLs (**Extended Data Fig. 5s**). In direct comparison with the QPMM approach, the GP approach also showed better classification performance (**Extended Data Fig. 5t**), with an area under the precision–recall curve of 0.69 compared with 0.60 for the QPMM approach.

### Proxy-based genetic correlations between parental exposure traits and child outcomes

Adverse environmental conditions and pollution have been identified as significant contributors to the high rates of childhood mortality, morbidity and disability^72^. Another notable benefit of JECS is its ability to assess environmental exposures of participating parents, including environmental pollutants such as heavy metals^9^ and other chemical substances^73^, as well as lifestyle factors (such as dietary habits, smoking, and alcohol consumption). GWAS-by-proxy of 209 exposure traits has identified 138 traits with at least one genome-wide association (*P* = 5. 0 × 10^−8^) (**Supplementary Table 1**; **Fig. 2e**), which could be a valuable addition to the knowledge of genetic determinants for children.

Even after accounting for the approximate four-fold reduction in effective sample size under the GWAS-by-proxy design^31^, our study detected a number of genetic associations to metal (mercury, cadmium, manganese and lead) or metalloid (selenium) exposure. The nominal sample size of 77,614 for those traits was almost 50 to 100 times larger than those studied previously^74,75^. We have observed pleiotropic effects among those traits. One of the outstanding effects was observed in ALDH2 locus (**Fig. 6a**), which is implicated in the metabolism of aldehydes, modulating drinking habits, and highly polymorphic in East Asian populations^76^. The locus was detected in maternal blood mercury and lead levels as well as subtypes of PFAS^77^. We observed the opposite effect direction between alcohol intake and blood mercury level at the missense SNP rs671 (**Fig. 6a**). The mechanism underlying this pattern remains unclear and may reflect pleiotropy, correlated behaviors, or other ALDH2-related metabolic pathways. Previous reports that alcohol consumption can increase mercury excretion in mice and humans^78,79^ provide one possible biological context, but our data do not establish a direct causal pathway from alcohol intake or ALDH2-related metabolism to lower mercury levels. For blood lead and PFAS levels, the effect directions were concordant with that for alcohol intake (**Fig. 6a**). In addition, the index variant for blood lead level differed from that for alcohol intake (**Fig. 6b–d**), suggesting that distinct mechanisms of metabolism or excretion may contribute to these associations. Furthermore, we sought to provide empirical support for the proxy-based findings at the ALDH2 locus, directly genotyped participating mothers (*N* = 63,257), including rs671 (**Methods**), and confirmed consistent effect directions and magnitudes (**Fig. 6a**). This analysis was intended as a locus-specific sensitivity check at ALDH2 and should not be interpreted as a genome-wide validation of the GWAS-by-proxy framework or as excluding false-positive associations at other loci.

**Figure 6.**
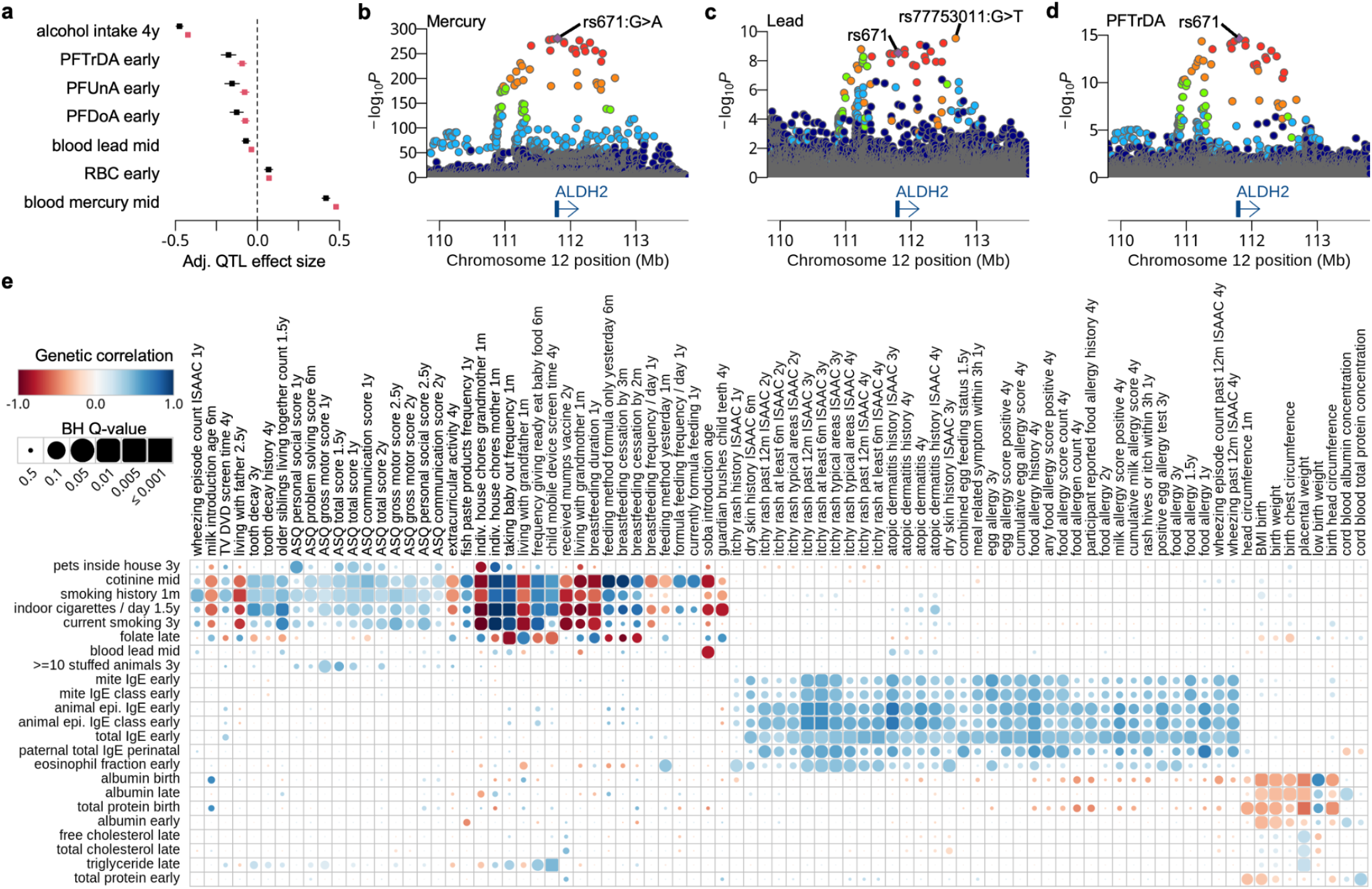
Genetics of environmental exposure traits for participating parents. **a.** QTL effect sizes of the missense SNP rs671 (G>A) at the ALDH2 locus across multiple environmental exposure traits (black: GWAS-by-proxy; red: GWAS using directly genotyped maternal data). Effect sizes are reported for the A allele relative to the G allele. Here the effect sizes from GWAS-by-proxy were rescaled (× 2) for visualization purposes to be comparable to those from GWAS using direct maternal genotyping. **b.** Locus zoom plot of the blood mercury level GWAS at the ALDH2 locus. **c.** Locus zoom plot of the blood lead level GWAS at the ALDH2 locus. **d.** Locus zoom plot of the blood PFTrDA level GWAS at the ALDH2 locus. **e.** Cross-trait LDSC analysis between parental environmental exposure traits and child health and developmental traits. Only trait combinations with Q-value<0.05 are shown in the heat map.

We then performed the cross-trait genetic correlation analysis, which revealed shared genetic architectures between exposure traits (**Extended Data Fig. 6; Supplementary Table 4**), such as positive correlations between maternal blood lead level and red blood cell counts (*r_g_*= 0. 52; *P* = 2. 0 × 10^−6^), and eosinophil percentage and various IgE levels (0. 42 ≤ *r_g_* ≤ 0. 52; *P* < 2. 0 × 10^−4^) during pregnancy. It is worth noting that paternal exposure traits (e.g., cholesterol levels) tend to be strongly clustered and not correlated well with maternal traits. This may reflect biological differences between the sexes and/or the influence of pregnancy on maternal physiology. It may also capture the contribution of fetal genetic effects specific to maternal physiology, as well as differences in trait measurement within the GWAS-by-proxy framework.

Finally, we investigated genetic correlations between parental environmental exposure traits and child health and developmental traits. Using genetic correlations obtained by the cross-trait LDSC regression, we identified several maternal IgE levels that were positively correlated with various allergic traits in children, such as itchy rash and food allergy (**Fig. 6e**). We also discovered maternal albumin and total protein levels during pregnancy, which are negatively correlated with the anthropometric traits of children at birth (**Fig. 6e**). Surprisingly, we found limited evidence for broad genetic correlation signals between most parental environmental exposure traits and child outcomes (**Supplementary Table 4**). For example, smoking exposure during pregnancy showed a genetic correlation with tooth decay at 4 years of age (*r_g_* = 0. 38; *Q* = 1. 1 × 10^−5^). However, we did not observe strong evidence that a specific genetic locus is colocalized between mother and child traits or shares a putative causal variant. These results do not provide strong evidence for a shared gene regulatory mechanism at specific loci. Moreover, genetic correlations estimated in the GWAS-by-proxy framework may be influenced by pleiotropy, fetal genetic effects, and correlated environmental or socio-behavioral factors, and therefore do not necessarily reflect direct or causal relationships between maternal exposures and child outcomes.

## Discussion

In this study, we conducted a large-scale GWAS of child health and development traits in more than 80K individuals, greatly expanding our knowledge of common genetic determinants in various early-life traits in an East Asian population. The study included 1,046 traits in children up to the age of four, as well as 209 environmental exposure traits in their parents. We identified 6,073 genetic associations across 21 different trait categories, some of which are highly pleiotropic at MHC or ALDH2 loci. In addition, we discovered a modest number of genetic correlations between children’s traits that go beyond their respective categories. The analysis also identified a stop-gain variant in *SERPINB7*, previously reported in Nagashima-type palmoplantar keratosis (NPPK), as a candidate locus shared between egg allergy and skin barrier dysfunction, suggesting a possible connection to therapeutic hypotheses.

Our analysis of longitudinal child BMI traits revealed novel and dynamic genetic associations during development, enabling us to construct individualized growth curves in the form of dynamic polygenic scores. Typically, the child’s growth is benchmarked against the population average. These personalized reference values may eventually contribute to the early detection and management of growth disorders and obesity by distinguishing between environmental influences and genetic predisposition. Recognizing such deviations could ultimately facilitate the early identification of children whose growth falls outside the genetically expected trajectory, thereby not only enabling the prediction and prevention of lifestyle-related diseases, but also prompting further evaluation for underlying etiologies, including rare congenital disorders^80,81^. This approach could also help to reduce overdiagnosis and overtreatment, as well as healthcare costs, while providing reassurance when growth is within expected ranges.

The MoBa evaluation should be interpreted as a trans-ancestry external validation of the JECS-trained dynamic PGS. Of the 238 JECS index variants, 200 were available and harmonized in MoBa, and the JECS–MoBa comparison was therefore based on this overlapping variant set. The lower variance explained in MoBa is consistent with the well-described attenuation of polygenic score transferability across populations, even for closely matched traits^82^. This attenuation may reflect differences in allele frequency, LD structure, variant tagging, phenotype measurement, age distribution, and environmental context. In addition, although JECS is large for a birth cohort, its discovery sample size remains modest (N ∼ 80,000) by contemporary GWAS/PGS standards, which may reduce effect-size precision and further limit transferability to a European population.

A further limitation of our longitudinal approach is that we did not apply established Bayesian shrinkage methods for polygenic score construction, such as PRS-CS^83^ or SBayesR^84^, to the GP-derived effects. These methods are primarily designed for cross-sectional GWAS summary statistics, where each variant is assigned a single scalar effect size for a given phenotype. In contrast, our model estimates an age-varying genetic effect as a continuous function over early childhood. Applying standard shrinkage methods at discrete ages would ignore the covariance structure across age and could break the smoothness and coherence of the estimated effect trajectories. Developing shrinkage or Bayesian polygenic prediction methods that operate directly on functional genetic effects remains an important area for future work.

A key consideration in interpreting GWAS-by-proxy results is that offspring genotypes capture a mixture of direct and indirect genetic effects. Associations with parental phenotypes may therefore reflect a combination of parental genetic effects, fetal genetic effects acting on maternal physiology, and indirect effects mediated through shared familial environments. These concerns are well established in family-based genetic studies: non-transmitted parental alleles can influence offspring phenotypes through parental environments, a process referred to as genetic nurture^85^, and population-based genetic associations may also be affected by indirect genetic effects, assortative mating, population structure, and shared familial environments^86–88^. These issues are particularly relevant for parental traits measured during pregnancy or early childhood, when parental and offspring environments are inherently shared. Consistent with these broader concerns, recent studies of GWAS-by-proxy have shown that proxy-based estimates may be biased by imperfect proxying, phenotype misclassification, residual confounding, and assortative mating^89,90^. Therefore, genetic correlations derived from this framework should be interpreted with caution and regarded as composite parent–offspring genetic signals rather than evidence for direct causal effects of maternal exposures. Nevertheless, for loci with well-established biological functions, such as ALDH2, the observed associations remain biologically informative, although contributions from fetal, indirect, or shared environmental effects cannot be excluded. Overall, GWAS-by-proxy enables large-scale hypothesis-generating analyses in the absence of parental genotypes, but does not allow definitive separation of maternal, paternal, fetal, and indirect genetic components.

Some of the traits analyzed in this study, particularly lifestyle-related, childrearing, and social traits, are strongly shaped by environmental, cultural, regional, socio-economic, healthcare-access, and family-structure factors. Although our sensitivity analyses did not provide strong evidence that these associations were primarily explained by the major genetic ancestry axes within the cohort, these analyses cannot fully exclude more local population structure, correlated environments, indirect genetic effects, or residual confounding. We therefore interpret loci identified for these phenotypes as statistical genetic associations that may reflect a mixture of direct genetic effects, correlated environments, indirect effects, and residual structure, rather than direct biological effects on the measured behaviors or social traits. These findings should be regarded as hypothesis-generating and as a resource for prioritising traits and loci for future follow-up analyses.

We also identified a limited number of genetic correlations between parental environmental exposure traits as well as maternal exposure traits during pregnancy and child traits after birth. One possible explanation for the modest number of observed correlations is that our GWAS-by-proxy approach reduces power to detect genetic associations in parental traits, approximately to one quarter in terms of chi-squared statistics under simple assumptions. Maternal genotyping is currently underway and will be completed in 2026. At that time, we will have substantially increased power to detect genetic associations of parental exposure traits, and we will also be able to disentangle genetic effects of children from those of parents on birth outcomes such as birth weight.

The analysis of mCA revealed that approximately 1% of study participants carried mCA in their cord blood samples. Although several previous studies have examined chromosomal mosaicism in early life, these investigations were limited by either small sample sizes or insufficient sensitivity. For example, Zhou et al.^91^ analyzed a total of 4,512 fetal samples derived from chorionic villus sampling, amniotic fluid, and cord blood combined, but the number of cord blood samples was considerably smaller than in our study. In addition, Li et al.^92^ defined chromosomal mosaicism using a ≥10% mosaic cell-fraction threshold, which inherently misses low–cell-fraction events. As a consequence, prior studies were unable to comprehensively characterize low-level mosaicism or to distinguish autosomal mCAs, LOX, and LOY with high precision. By using a large cohort of more than 80,000 cord blood samples and a high-resolution haplotype-based method (MoChA), our study provides the first robust, population-level estimate of neonatal mCA frequencies that is directly comparable to adult datasets such as BBJ. As mCAs are strongly associated with clonal hematopoiesis and increased risks of cancer^93^ and infertility^94^ in adulthood, it is imperative to follow up participants over the course of their lives to ascertain whether participants with mCAs in early life develop diseases in later life. However, two important caveats should be noted. First, because the prevalence of mCA in cord blood is expected to be low^91,92^, the positive predictive value of the MoChA program is likely lower than in adult samples, implying a higher false-positive rate. Second, the higher LOY prevalence observed in this study compared with previous reports^20,95,96^ is likely due to the absence of a lower cutoff for cell fraction and the inclusion of partial mCA events (**Methods**).

Our analyses primarily identified common genetic variants, which is consistent with the well-known properties of genome-wide association studies, where statistical power and imputation accuracy are generally higher for common alleles^97^. Future studies incorporating exome or whole-genome sequencing will be essential to capture the contribution of rare variants, which are often poorly tagged by genotyping arrays^98^. In addition, the integration of both short- and long-read sequencing technologies will enable more comprehensive detection of complex structural variants, which are increasingly recognized as important contributors to human traits and disease^99^. Such approaches will be particularly valuable for elucidating the genetic architecture of child health and developmental traits, which may involve a broader spectrum of genetic variation than can be captured by array-based GWAS alone.

To fully connect variants to their biological functions, we also need to fine-map each genomic locus discovered through GWAS. For example, we could use SuSiE^100^ to identify secondary hits in each locus and integrate genomic annotations, such as ChIP-seq or chromatin accessibility peaks used in sLDSC^53^ or fGWAS^101^. We could also colocalize with public eQTL databases, such as GTEx^102^, to annotate functional genes in non-coding GWAS loci. These directions remain important areas for future investigation.

JECS will continue until the participating children reach the age of 18 and beyond. The phenotype data freeze up to the age of 8 will soon be available to further increase the number of phenotypes analyzed in GWAS. We are continuing to uncover the genetic determinants of various childhood traits in early life, which may have the potential to prevent various adult diseases at a much earlier stage or even during pregnancy.

## Data Availability

The manifest file of the Illumina Asian Screening Array (ASA-24v1-0_E2.csv) was available from the product web page (https://support.illumina.com/downloads/infinium-asian-screening-array-v1-0-product-files.html). The allele frequency data of Tohoku Medical Megabank (ToMMo) samples was obtained from jMorp (https://jmorp.megabank.tohoku.ac.jp/). The latest GWAS results were downloaded from the GWAS Catalog (https://www.ebi.ac.uk/gwas/api/search/downloads/gwas_catalog_v1.0-associations_e113_r2024-11-03.tsv). Two imputation reference panels were obtained from the 1KGP (ALL.chr*.shapeit2_integrated_snvindels_v2a_27022019.GRCh38.phased.vcf.gz and 1kGP_high_coverage_Illumina.chr*.filtered.SNV_INDEL_SV_phased_panel.vcf.gz). The variant VCF of a South Korean population was obtained from the Korean Personal Genome Project (KPGP) (http://kpgp.kr/). The pre-computed LD scores using the 1000 Genomes Project (1KGP) East Asian Population (1000G_Phase3_EAS_baselineLD_v2.2_ldscores.tgz) was downloaded from ZENODO (https://zenodo.org/records/10515792). GTEx eQTL summary statistics of the skeletal muscle tissue (V8) were downloaded from the eQTL Catalogue (ftp.ebi.ac.uk/pub/databases/spot/eQTL/sumstats/QTS000015/QTD000121/QTD000121.all.tsv.gz). Ensembl gene annotation (version 113) was downloaded from BioMart (https://www.ensembl.org/). GWAS results of childhood BMI from MoBa were downloaded from the NIPH website (https://www.fhi.no/en/ch/studies/moba/for-forskere-artikler/gwas-data-from-moba/). Raw genotype data for child participants in JECS have not been uploaded to a publicly accessible repository due to parental consent, ethical restrictions, and legal framework of the Act on the Protection of Personal Information (Act No. 57 of 30 May 2003, amendment on 1 June 2025). Access to the raw data can be granted for scientific purposes only at the JECS facilities located in Japan. All inquiries about data access should be sent to. The person responsible for handling enquiries sent to this e-mail address is Dr Shoji F. Nakayama, JECS Programme Office, National Institute for Environmental Studies. The GWAS summary statistics for all 1,255 traits are available via the JECS PheWeb (https://jecs-pheweb.ncchd.go.jp/).

## Acknowledgement

We thank all the JECS participants. We would like to also thank all staff members of the JECS and the informatics group at National Center for Child Health and Development. We thank the members of the Kawasaki disease medical record transcription board (Drs. Sayaka Fukuda and Tohru Kobayashi). We thank Kazuyuki Nakazono, Naoko Miyagawa and Satoshi Nagashima for their contribution to the GWAS pipeline construction. This study was funded by the Ministry of the Environment, Japan. The findings and conclusions of this article are solely the responsibility of the authors and do not represent the official views of the above government.

## Methods

### Ethical compliance

JECS protocol was reviewed and approved by the Ministry of the Environment’s Institutional Review Board on Epidemiological Studies (No.100910001) and the Ethics Committees of all participating institutions. Written informed consent was obtained from all children’s legal guardians. Following informed consent, participants were given further information about SNP genotyping using microarray technology and the possibility to withdraw from the genetic study at any time.

### Data source

The present analyses were conducted using the jecs-ta-20190930 dataset, which encompasses follow-up data until 3 years of age for the children, maternal/cord blood laboratory test results, maternal blood concentrations of five elements (lead, cadmium, mercury, selenium, and manganese), and maternal urinary cotinine concentrations. Additionally, the jecs-qa-20210401 dataset (including 103,057 pregnancies and 100,300 live births) was used, extending follow-up to 4 years of age and including data on maternal plasma concentrations of PFAS.

### Collection of cord blood samples

JECS collected a single cord blood sample at delivery, even in cases of multiple births. For monozygotic twins or triplets, genotypes obtained from SNP arrays are expected to be identical for common variants, whereas dizygotic twins are genetically equivalent to siblings. When multiple-birth records were linked to a single cord blood genotype, we assigned the same genotype data of a twin to the co-twin(s), unless the genotype and clinical sex were inconsistent, and we excluded the child whose reported sex was inconsistent with the genotype-inferred sex. This procedure allowed partial resolution of dizygotic twin pairs when the co-twins were of opposite sex. However, same-sex dizygotic twins could not be distinguished from monozygotic twins using genotype data alone. Based on the reported ratio of monozygotic to dizygotic twins in Japan^103^, we estimate that approximately 100–200 non-genotyped dizygotic co-twins may remain represented in the analysis.

### DNA extraction from cord blood samples

DNA from whole cord blood was extracted using the QIAsymphony SP system and QIAsymphony DSP DNA Midi Kit (QIAGEN). Genomic DNA was quantitated on the SpectraMax M2e Microplate Reader (MOLECULAR DEVICE) and the Quant-iT PicoGreen dsDNA Assay Kit (Thermo Fisher Scientific).

### SNP genotyping

We genotyped 80,639 cord blood samples using the Illumina Asian Screening Array (ASA-24v1-0_E2) BeadChip SNP array (https://www.illumina.com/products/by-type/microarray-kits/infinium-asian-screening.html). We conducted genotype calling on Illumina GenomeStudio software to generate PED files. We excluded samples with sample call rate <0.98. These PED files were first converted into the VCF format using PLINK2 (**Code availability**). We used the fixref plugin of bcftools to align the strand orientation of each SNP (**Code availability**). For palindromic SNPs (A-T or C-G SNPs), BLAST software (ver 2.5.0) was used to identify the SNP probe orientations a priori (**Code availability**). After fixing the reference alleles, variants with a variant call rate less than 0.99, a HWE *P*-value less than 10^-6^ and an allele frequency that deviated from four times the standard error (SE) of the allele frequency estimate for the BioBank Japan (BBJ) or the Tohoku Medical Megabank (ToMMo) samples (**Data availability**) were excluded. We estimate that the BBJ and ToMMo allele frequencies have a SE of with sample sizes of 13,406 and 54,000, respectively. The genotype data was then pre-phased using the Eagle software (**Code availability**) without reference haplotype data because the sample size was sufficient for self-inference of population-specific haplotypes. Finally, the phased genotype data (with 617,557 variants) was harmonized with the 1000 Genomes Project VCF (**Data availability**) and then missing genotypes were imputed using the Minimac4 software (**Code availability**).

### Detection of chromosomal aberrations

Chromosomal aberrations were detected from the signal intensity data for each DNA sample. Nominal XY signal intensities were extracted from Illumina idat files and then converted into B allele frequency (BAF) using the polar coordinate transformation. Here, A and B denote the generic allele labels used by Illumina microarrays, and the BAF intuitively represents the estimated proportion of the B allele at each locus. After hybridization and scanning, the fluorescence intensities for these two alleles are measured. Note that, the A/B alleles are defined in the manifest file of Illumina SNP genotyping arrays (**Data availability**), where each SNP is designed with two probes corresponding to the reference (“A”) and the alternative (“B”) allele. However, this definition is irrelevant for the “Ref” and “Alt” alleles based on the human reference genome sequence. A mixture of Gaussian mixtures was fitted to detect the number of copies of each autosomal or sex chromosome. The means and variances for the Gaussian mixture distributions were fixed and posterior probabilities of aberrations were obtained.

### MoChA analysis for detecting mosaic chromosomal alterations

Illumina idat files were processed to generate VCF files including BAF and log R ratio (LRR) values for each variant. To detect somatic mosaic chromosomal alterations (mCAs), we further analyzed the BAF and LRR data using MoChA (**Code availability**), with GRCh38 as the reference genome. Haplotype phasing was performed with SHAPEIT5 (**Code availability**), following the established protocol of MoChA. To minimize the inclusion of germline aberrations, events with the cell fraction greater than 0.5 were excluded. However, we did not introduce a lower cutoff value for the cell fraction. Additionally, we filtered out events with relative coverage (rel_cov) ≥ 2.5, as these may represent germline duplications or technical artifacts.

To compare with adult samples, we additionally analyzed participants from the first (BBJ1) and second (BBJ2) cohorts of the BioBank Japan Project^104,105^. The mean age of BBJ1 participants was 62.7 (N = 179,254), and the mean age of BBJ2 participants was 65.2 (N = 71,469).

### Population stratification analysis using principal component analysis and ADMIXTURE

Population stratification analysis was performed by principal component analysis (PCA) of genotype dosages with reference populations obtained from the 1000 Genomes Project (1KGP) (**Data availability**) and Korean Personal Genome Project (KPGP) (**Data availability**). Korean genotype data (referred to as KOR) based on whole genome sequencing was harmonized with the 1000 Genomes Project VCF and then missing genotypes were imputed using BEAGLE software (**Code availability**). SNP loci were chosen based on our sample statistics with MAF ≥ 1% and HWE *P*-value > 1.0 × 10^−6^. Then, linkage disequilibrium pruning was performed using “--indep-pairwise 50 10 0.1” in PLINK2, yielding 165,756 biallelic SNPs. PCA was performed using the PLINK2 “--pca” command with the “allele-wts” modifier to obtain the first 20 principal components (PCs) as well as SNP loadings. The first two components were shown in **Fig. 1e**.

We then clustered JECS participants in principal component space by anchoring the known worldwide populations AFR, EUR, AMR, SAS, and EAS (including KOR) using the first 7 genotype principal components. A Gaussian mixture model was fitted to identify individuals with admixed ancestry, including those with approximately half or one-quarter ancestry from a given reference population (for example, Japanese–European mixed ancestry), and only samples assigned to the East Asian cluster were retained. We next performed principal component analysis in the East Asian subset together with JPT, CHB, and KOR reference samples from the 1KGP and the KPGP; the first two components are shown in **Fig. 1f**. Finally, we selected the first 7 PCs as covariates, as their SNP loadings were uniformly distributed across the genome.

The ADMIXTURE analysis (**Code availability**) was also applied to the same data set including the worldwide populations used in the PCA (**Fig. 1e**). We ran the ADMIXTURE software with K=5 and the default parameter settings.

### Phenotypes

Phenotype data were derived from the Japan Environment and Children’s Study (JECS), a large-scale birth cohort with longitudinal measurements collected from pregnancy through early childhood. We extracted 4,460 variables from 23 data sheets encompassing questionnaire responses, clinical measurements, and environmental exposures in children up to 4 years of age and their parents (all analyses were conducted using datasets named jecs-ta-20190930 and jecs-qa-20210401). Variables were first screened for usability based on predefined criteria, excluding metadata, duplicate entries, variables outside the scope of the study, and those with ambiguous definitions. The remaining variables were classified into candidate traits, composite trait source variables, and covariate source variables according to their role in downstream analyses.

All retained variables underwent standardized quality control and transformation procedures. For continuous variables, outliers were excluded using a four-standard-deviation (4SD) criterion unless clear physiological or measurement-based bounds were defined a priori (e.g., laboratory measurements or questionnaire-derived scores with fixed ranges), in which case lower and upper bounds were applied instead. The choice between 4SD filtering and bounded exclusion was determined based on clinical plausibility and measurement characteristics, as reviewed by domain experts. Binary and categorical variables were harmonized to ensure consistent encoding, and additional processing included conditional correction of nested questionnaire responses and exclusion of non-maternal respondents for respondent-dependent traits. The specific QC criteria applied to each variable are provided in **Supplementary Table 6**.

We then constructed traits from these variables, including both directly analyzable variables and composite traits derived from multiple source variables (e.g., BMI, allergy-related traits, and developmental measures). Variables that were not suitable for independent analysis (e.g., due to low interpretability, copyright restrictions, or distributional properties) were incorporated into composite traits where appropriate. Covariates were selected from a subset of variables representing participant metadata or potential confounders (e.g., gestational age), with redundancy removed where necessary.

After trait construction, we considered several traits unsuitable because they had too many missing responses (low call rate) or were too low in complexity (*i.e.* too many identical values/answers in the trait), leading to high false positives. We calculated the call rate of a trait, such as

`(call rate) = (number of valid data) / (number of questionnaires collected at the given age of the children)`,

and excluded traits with more than 40% missing responses from the analysis. The complexity score, defined as

`(complexity) = 1.0 - (frequency of the most frequent value in the phenotype data) / (number of observations without missing values)`,

was also calculated for each trait (note that this score should be the case ratio for a case-control trait). We removed binary traits with a score of less than 0.01 and quantitative traits with a score of less than 0.05 to reduce potential false positives in GWAS. A total of 1,255 parental and child traits were analyzed in the GWAS.

To ensure transparency and reproducibility, we systematically documented the full mapping from raw variables to derived traits, including excluded variables, covariates, and composite trait definitions, in the **Supplementary Notes 1** and **Supplementary Tables 6** and **7**.

### Genome-wide association study (GWAS)

GWAS of all 1,255 traits were performed using REGENIE (**Code availability**). In the first step, 546,576 SNPs directly genotyped on the Illumina ASA chip were used to obtain the leave-one-chromosome-out (LOCO) predictions. In the second step, 17,454,650 imputed variants (MAF greater than 0.1% and imputation score R2 greater than 0.3) were used for GWAS. Quantitative and ordered categorical traits were transformed using the rank-based inverse normal transformation (RINT) for genome-wide quantitative trait locus (QTL) mapping. Non-ordered categorical traits were transformed into a set of binary traits using the one-hot encoding, and the Firth logistic regression was used to perform GWAS in conjunction with other binary phenotypes (such as disease onset).

Covariates used for each trait were determined based on the child’s age in months at the time the questionnaire was filled out. For the traits at birth, we used (1) mother’s age at birth, (2) gestational duration (in days), (3) infant’s sex, (4) the first 7 genotype PCs. For the traits from 6 months to 4 years old of children, we also included the child’s age in months as a covariate. For the caregivers’ traits, age of the caregivers and the first 7 genotype PCs were used.

Gene annotation was applied using ANNOVAR (**Code availability**) with the Ensembl 113 gene annotation (**Data availability**). The rsID of each variant was assigned using ANNOVAR with the dbSNP version 151 as the default (**Code availability**).

A GWAS locus was defined by a variant with a *P*-value below the standard GWAS threshold of 5. 0 × 10 with 500 kb of upstream and downstream sequence (1 Mb window). If there are multiple variants with *P*-values below the standard GWAS threshold within the 1 Mb window, we group them as the same locus, and extend the window by 500 kb upstream and downstream for the set of variants until no variants exceed the standard GWAS threshold. The index variant in the locus was then defined as the variant with the minimum *P*-value within the window. The secondary independent variants within the same region were discovered by the REGENIE’s conditional analysis given the primary index variant(s) as the covariate(s). We iteratively performed the conditional analysis until no variants exceed the standard GWAS threshold within the window.

We used our imputed genotype to compute the linkage disequilibrium (LD) *r*^2^ index between all index variants in our study and variants in the GWAS Catalog Release 2024-11-03 (**Data availability**). We computed the LD index only for pairs of variants within 1 Mb distance between our index variant and the GWAS catalog variant. We omitted any GWAS catalog variant that was not imputed in our genotype data. A total of 231,405 out of 296,928 GWAS Catalog variants were covered by the analysis, excluding genotype-by-genotype interactions.

### HLA imputation and association analysis

We performed HLA imputation using a Japanese population–specific reference panel (n = 1,118) constructed in a previous study^106^. We applied DEEP*HLA (**Code availability**), a deep learning-based HLA imputation method^33^, to the genotyped SNPs in the MHC region (25 to 35 Mb on chromosome 6) and imputed the classical HLA alleles (one-, two-, and three-field [two-, four-, and six-digit]) and amino acid variants of the classical and non-classical HLA genes. HLA variants imputed with an imputation quality score (R2 in 10-fold cross-validation) > 0.7 and MAF > 0.005 were used for further analyses.

Associations between HLA variants and traits were evaluated using REGENIE (**Code availability**), following the same framework as the GWAS. To identify independently associated HLA loci, we performed step-wise conditional analyses by additionally including all classical HLA alleles of the HLA gene to which the lead variant belonged as covariates^106^. When the lead signals were *HLA-DRB2–9* variants and were in strong LD with *HLA-DRB1* variants (R2 > 0.9), we jointly included *HLA-DRB1* classical alleles in the model, considering that signals in the *HLA-DRB2-9* region are often driven by strong LD with *HLA-DRB1*, which is the primary functional locus in this region.

### GWAS with unrelated participants

To assess the robustness of our GWAS results to relatedness, we constructed a subset of unrelated individuals by excluding samples with kinship coefficients greater than 0.0442, corresponding to third-degree relatives or closer, as estimated using KING^21^. From each related pair, one individual was preferentially retained based on higher genotype call rate. The resulting subset (*N*=70,751) was used to perform GWAS using the same analytical framework as in the full dataset.

In the subsample analysis, association test statistics were interpreted under the assumption that χ² statistics for non-null variants follow a non-central chi-square distribution with one degree of freedom. Under this model, the non-centrality parameter is proportional to the sample size, such that the expected test statistics in the subsample are scaled by the ratio of sample sizes relative to the full dataset. A full derivation of the underlying statistical framework, including all relevant equations, is provided in **Supplementary Notes 2**, Appendix 2.1 and 2.2.

### GWAS for ancestry axes

To assess the influence of population structure, we conducted GWAS using genotype principal components (PCs) as quantitative traits (ancestry axes). The first two PCs, which capture the major axes of genetic variation within the cohort, were used as the basis for defining ancestry-related variation. To facilitate biological interpretation, we applied a linear rotation of these PCs using a rotation matrix to derive orthogonal axes corresponding to variation between (i) Japanese and other East Asian populations, and (ii) mainland Japanese (Hondo) and Ryukyu ancestries.

GWAS was then performed on these rotated axes using REGENIE, treating each axis as a quantitative phenotype. The analyses were conducted using the same pipeline as in the primary GWAS; however, genotype principal components were not included as covariates in these models, as they constitute the outcome variables. This approach enables identification of genetic variants associated with ancestry-related variation and allows evaluation of potential confounding due to population structure.

This analysis was used as a SNP-level negative-control diagnostic for residual population structure, rather than as a biological GWAS of genetic PCs. Because genotype PCs are linear combinations of allele dosages, GWAS summary statistics for these ancestry axes capture SNP-level allele-frequency gradients aligned with the major axes of genetic variation in the cohort. We used these ancestry-axis GWAS summary statistics in cross-trait LDSC analyses to evaluate whether the residual SNP-level association patterns of child traits, after standard GWAS adjustment, were aligned with ancestry-related allele-frequency patterns. This differs from directly regressing phenotypes on genetic PCs, which tests whether phenotype means vary along ancestry axes but does not assess whether the genome-wide SNP-level association signals themselves are driven by genetic drift or residual population structure. Accordingly, any correlation with ancestry-axis GWAS summary statistics was interpreted only as a diagnostic signal of potential population-structure confounding, not as evidence of biological genetic correlation with the PCs.

### LD score regression

The Python package for LD Score regression (LDSC) was downloaded from GitHub (**Code availability**). The pre-computed LD scores in an East Asian population (v2.2) were downloaded from Zenodo (**Data availability**). The additive SNP heritability (*h*^2^) was estimated for each of 1,255 traits using the baseline LD scores (the heritability estimates are provided in **Supplementary Table 1**). For case-control traits, we computed the effective sample size (*N*_eff_) following the standard LD score regression (LDSC) framework, using *N_eff_* = 4/((1/*N_case_*) + (1/*N_control_*)), and applied these values consistently in all LDSC analyses. We now provide the additive SNP heritability, its standard error, *Z*-score and *N*_eff_ value for each trait in the **Supplementary Table 1**.

For the cross-trait LDSC analysis between pairs of child and parent traits to compute genetic correlations (*r_g_*), those traits with *h*^2^ greater than 0.0 and one or more associations in GWAS were used. The environmental correlation (*r_e_*), which is the residual correlation after subtracting genetic correlation from the phenotypic correlation, was computed as follows

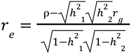

\where ρ indicates the total covariance, h^2^_1_ and h^2^_2_ indicate the additive SNP heritability estimates for the first and the second traits, respectively. Note that the genetic covariance is computed as 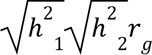. We provide the full detail of the derivation in the Appendix section 2.4 of the **Supplementary Notes 2**. The computed genetic correlations and environmental correlations for all the analyzed traits are available in **Supplementary Table 4**.

### scDRS

scDRS^55^ analysis was performed to assess cell type-specific enrichment of genetic risk across PBMC cell types from recent single-cell multi-omics data^56^. Gene-level association statistics were first obtained from GWAS summary statistics using MAGMA^107^ with the 1000 Genomes Phase 3 East Asian reference panel and were then used to construct scDRS input gene sets, as described previously^55^. Cell type-level enrichment was evaluated by the group-level downstream analysis.

### GWAS of parental traits using proxy genotypes (GWAS-by-proxy)

We first removed any parental trait that was observed from a mixture of mother or father or other family members (*e.g.* the question: *An estimate of cigarettes smoked inside the house per day*). For each parental trait, we regressed the maternal (or paternal) phenotype on the child’s imputed genotype dosages using REGENIE’s mixed-model framework (**Code availability**), analogous to the child-trait GWAS (see above). We used the same quality-filtered variants and covariates except for the child sex and the gestational duration, and we restricted to parent–child pairs with available phenotypes. Here, the offspring genotype acts as a proxy for the parental genotype, with an expected correlation of 0.5. The effect estimates were then attenuated by half, relative to a genome-wide association study (GWAS) based on parental genotypes under the assumption of no additional indirect genetic effects or assortative mating. In the **Supplementary Table 2**, we reported unadjusted effect sizes from the proxy analyses (except for Fig. 6a for visualization purposes), focusing on the presence and direction of associations. Therefore, caution is required when interpreting the effect sizes.

### Locus-specific sensitivity analysis of rs671 GWAS-by-proxy associations using maternal genotype data

The maternal genotyping using blood samples for the first and the second batches (*N*=80,964) was performed under conditions identical to those applied to the children. Genotype QC included exclusion of six samples with a call rate < 0.98. We then removed 11 samples with sex inconsistency, as all maternal samples were expected to be female. Next, we used KING^21^ to verify parent relationships and identify duplicate samples, particularly because multiple siblings from the same family participated in the study, which resulted in some mothers appearing more than once in the dataset. We retained mothers genetically confirmed to be linked to children included in the GWAS, with duplicate maternal samples consolidated by preferentially keeping the sample with the higher call rate, leaving 63,257 final maternal samples for the rs671 association analysis. The analysis was performed using REGENIE in the same manner as described above for the children, except that the second step was performed only for rs671. Covariates included maternal age at the child’s birth and the first eight PCs of maternal genotypes.

As this analysis was designed as a locus-specific validation of rs671, we did not perform whole-genome imputation or genome-wide analyses in the maternal dataset. Extending the analysis genome-wide would require comprehensive parent–offspring genotype harmonization and quality control, including detection of sample swaps, and imputation-level variant QC, which is currently ongoing and outside the scope of this locus-specific sensitivity analysis.

### Effect size comparison with MoBa’s childhood BMI GWAS

We downloaded the childhood BMI GWAS results from the Norwegian Mother, Father and Child Cohort (MoBa) at eight different ages (at birth, 6 weeks, 6 months, 8 months, 1 year, 1.5 years, 2 years and 3 years old, sample size ranging from 16,902 to 28,681 children) from the NIPH website (**data availability**) and linked them with our GWAS age strata (0 month, 1 month, 6 months, 1 year (early), 1 year (late), 1.5 years, 2 years and 3 years old). We used the UCSC Liftover tool to map GRCh37 coordinates to GRCh38 coordinates of GWAS results from MoBa. Then, we extracted the effect size and its standard error for each index variant discovered in our GWAS of the specific age stratum. Of the 454 index variants in our GWAS, 208 matched the coordinates, reference, and alternative alleles. After checking the *P*-values, we found that only 18 variants passed the standard GWAS threshold of *P* = 5 × 10^−8^.

### Longitudinal analysis using Gaussian processes

A Gaussian process (GP) regression model was used to map dynamic genetic associations along a child’s age in months. We first clarify that this GP regression model builds on the same GP-based genetic-association framework as GASPACHO^58^, but with important differences in the data structure and implementation. GASPACHO was developed for single-cell eQTL mapping, where latent cellular states are inferred from high-dimensional gene-expression data. In the present study, this latent-space inference step is unnecessary because the relevant longitudinal coordinate, child age, is directly observed. In addition, we improved the computational implementation so that the model can be applied to biobank-scale longitudinal cohort data with tens of thousands of participants and repeated measurements, while explicitly accounting for familial relatedness using KING-derived relatedness information. In contrast, the original GASPACHO study was applied to a much smaller set of unrelated donors (N=68).

We used inducing points within a sparse GP framework^108^ to handle the large number of observations (N = 684,228) from 79,743 children who had at least one BMI record from ages 0 to 4 (approximately 8 time points per participant). We included all the covariates used in our standard GWAS, as well as the location where the child’s height and weight were measured (community health center, nursery, hospital or home). Model parameters were estimated to maximize the Titsias lower bound^109^ of the model using the quasi-Newton method. To ensure stable parameter optimization, the length-scale parameter of the quadratic exponential kernel used in the Gaussian process regression model and a subset of the variance parameters were optimized sequentially across the three-step procedure (**Supplementary Note 2**).

During the parameter tuning, we found that the length-scale parameter was suboptimal. Because this parameter controls the smoothness and correlation structure of the longitudinal trajectory, its inappropriate specification led to overly optimistic association statistics and, consequently, an excess of significant findings. To address this issue, we fit the model with improved calibration of the kernel length-scale parameter with an inverse-Gamma prior distribution whose shape and scale parameters were set to 0.01*N_d_* and 5*N_d_*, respectively, where *N_d_* denotes the number of donors/individuals used in the dataset (not the number of data points). For sufficiently large *N_d_*, this prior has an approximate mean of 500.

Here, unmodeled familial relatedness could affect the calibration of longitudinal association tests, particularly because repeated BMI measurements from related children may be correlated beyond the within-individual longitudinal correlation. To address this concern, we incorporated pairwise kinship coefficients estimated by KING^21^ to construct an additive relationship matrix *A* and replaced the donor-level covariance *ZZ*^T^ with *ZAZ*^T^ where *Z* denotes the design matrix of individuals. Critically, relatedness is incorporated both as a time-invariant familial covariance component and as an age-dependent familial covariance component through *K*_age_⦿*ZAZ*^T^, while retaining the population-average age kernel *K*_age_ as a latent random function. This allows the model to account for correlated longitudinal trajectories among related children, including siblings and cousins up to the third degree, in addition to repeated measurements within the same child.

The dynamic QTL effect was defined as any effect that exhibits a deviation from zero over the course of time. The null hypothesis is the genetic effect being equivalent to zero for the duration of the modeling period (in this case, from 0 to 54 months of age). For each variant, we used a score test that evaluates whether introducing a genetic-effect function improves the model fit, while estimating the variance components only under the null model without fully refitting the alternative model for every variant. This score test should be interpreted as an omnibus longitudinal association test, because the alternative hypothesis includes any smooth non-zero genetic effect across age, including both approximately constant and age-varying effects. The dynamic effect and its standard error were estimated as the posterior distribution of the Gaussian process modeling the QTL effect. The score statistics to capture the dynamic QTL effects were computed without estimating the variance parameters under the alternative hypotheses, which dramatically reduces the computational time. The CompQuadForm package implemented in R was used to compute the *P*-value of a weighted sum of independent chi-square statistics (see the **Supplementary Notes 2** and ref^59^ for further details). We identified, in total, 158 dynamic QTL loci that passed the genome-wide significance threshold (of which 41 were not discovered by the cross-sectional GWAS as mentioned in the main text). The FTO locus is a special case where the effect reverses sign within the modeling period; many other dynamic loci show an effect that either emerges or attenuates over time but does not change sign (effect size plots for all the associated variants are shown in the figures in **Supplementary Notes 2**).

The LDSC regression was performed using the score test *P*-values, from which *Z*-scores were computed. Here, the QTL effect beta was set to a constant value of 1.0 for the analysis, because it was essentially not a single value in the model; rather, it varied across each data point in the model as a Gaussian process. Here, the standard LDSC regression does not account for the sign or magnitude of beta in the analysis, and the approximation of the Z-score is valid because the sample size is large.

To enable comparison with the longitudinal BMI GWAS, we also performed GWAS of raw BMI without applying RINT in each of the 11 age strata. The raw-BMI effect-size estimates were used for comparison, whereas statistical significance was evaluated using the corresponding GWAS of RINT-transformed BMI.

### Simulation and permutation for generating null genotypes

For null simulations, we generated genotype data for biallelic SNP loci under Hardy-Weinberg equilibrium while preserving the observed family-relatedness structure. For each simulated locus, the minor allele frequency *p* was sampled independently from a uniform distribution U(0.001,0.5). Genotype dosages for 79,743 individuals were then generated using a Gaussian copula based on a family-wise Cholesky representation of the additive relationship matrix constructed from KING relatedness estimates. Specifically, correlated standard normal variables were generated within each family block using the Cholesky factors, and were then thresholded according to the Hardy-Weinberg genotype probabilities (1−*p*)^2^, 2*p*(1−*p*), and *p*^2^ to obtain dosages 0, 1, and 2. In total, we generated genotype data for 17,454,650 SNP loci under the null hypothesis, matching the number of tests performed in the actual BMI GWAS.

For the permutation test using the real genotype data, we generated family-size-preserving permutation indices using the KING relatedness results after grouping related individuals up to the third degree. Individuals were first grouped by family identifier, and families were stratified according to their size. Within each family-size stratum, family blocks were randomly permuted, and all individuals within a block were moved together while preserving their within-family ordering. This procedure preserves the distribution of family sizes and the block structure of the relatedness design matrix, while breaking the correspondence between genotype and phenotype across family blocks. In this analysis, the same family-aware sample-label permutation was applied within each block of 5,000 variants to preserve the LD structure among variants in that block; a different permutation was then used for the next block. This function was also implemented in the DynamicPGS package (**Code availability**).

### Age-stratified meta-analysis for BMI GWAS

We used METAL (**Code availability**) to perform a fixed-effect meta-analysis across 11 age-specific BMI GWAS. At each age, BMI was RINT-transformed, and QTL effect estimates were combined using inverse-variance weighting. For the permutation analysis, BMI values across all 11 age strata were permuted once using the same individual-level permutation index across ages. Covariates were permuted using the same index to preserve the phenotype–covariate relationships. This procedure preserves each child’s longitudinal BMI trajectory, measurement schedule, and covariate structure while disrupting the genotype–phenotype relationship. We used the same covariates for both the original and permuted datasets, and GWAS was performed using REGENIE as described above.

### Quadratic polynomial mixed-effects model (QPMM)

To benchmark the Gaussian process framework against a conventional longitudinal model, we fitted a quadratic genotype-by-age interaction linear mixed-effects model (QPMM) for each variant using the lme4 package implemented in R. Child age in months was centered and scaled (`age`), and its squared term was included as `age2` to capture the nonlinear BMI trajectory. The null and alternative models were specified as follows:

`fit0 <- lme4::lmer(y ∼ age + age2 + covs + (1 + age + age2 | iid))`

`fit1 <- lme4::lmer(y ∼ g * age + g * age2 + covs + (1 + age + age2 | iid))`

Here, `y` denotes BMI, `g` denotes genotype dosage, `iid` denotes individual ID, and `covs` denotes a matrix of covariates. We included all the covariates used in our standard GWAS, as well as the location where the child’s height and weight were measured (community health center, nursery, hospital or home). For the mock data analysis, we simulated only two principal components and child sex as covariates to simplify the analysis and facilitate rapid optimization. The random-effects structure `(1 + age + age2 | iid)` allowed each child to have an individual-specific intercept, linear age slope, and quadratic age slope. Evidence for longitudinal genetic association was assessed by a likelihood-ratio test comparing the null and alternative models using `anova(fit0, fit1)`. This corresponds to a 3-degree-of-freedom test of the genotype main effect and its interactions with age and age squared. The model was used as a parametric comparator to evaluate whether the GP-based approach identified associations beyond those captured by a quadratic polynomial representation of age-dependent genetic effects. Note that we also set `REML = FALSE` in `lmer()` during the model fitting.

Based on pilot runtime estimates, applying this model to all genome-wide variants was expected to require approximately two months even on our parallel computing environment. We therefore restricted the comparison to 63,640 variants that showed suggestive evidence in the GP-based longitudinal association test (*P* < 1×10⁻⁴). The QPMM was then fitted to this subset of variants to evaluate whether the GP-identified signals were also supported by QPMM as a conventional parametric longitudinal model.

The QPMM can be viewed as a special case of the GP framework in which the sparse GP cross-kernel matrix *K_NM_* is replaced by the quadratic polynomial basis (1, age, age^2^), while the covariance matrix *K_MM_* for the latent GP at the inducing points is specified as a scaled identity matrix (σ^2^*I*). In this formulation, there is no separately estimated kernel length-scale parameter. Instead, the scale over which the genetic effect varies with age is implicitly fixed by the a priori standardization of the age and age-squared terms using prespecified, and therefore partly arbitrary, scaling constants. Consequently, the smoothness and relative contribution of the polynomial components are determined by these scaling choices rather than learned from the data.

### Estimation of QPMM associations missed by GP screening

To assess whether restricting the polynomial analysis to variants with a GP *P* < 10⁻⁴ could have excluded variants reaching genome-wide significance in QPMM, we estimated the probability of such an event for every screened-out variant. Among variants evaluated by both methods, the GP and QPMM *P*-values were converted to one-degree-of-freedom chi-square statistics. We fitted a linear regression of the polynomial statistic on the GP statistic and calculated their Pearson correlation. The regression slope was used to describe the relative strength of the QPMM statistic.

We represented the two chi-square statistics as squares of correlated latent normal variables. Their latent correlation was approximated as the square root of the observed correlation between the chi-square statistics. For each screened-out variant, the noncentrality parameter was estimated by the observed GP chi-square statistic. The corresponding noncentrality for QPMM was approximated by multiplying this estimate by the fitted regression slope.

Using the resulting variant-specific noncentrality parameters and the estimated latent correlation, we evaluated the joint distribution of the GP and QPMM statistics. The GP screening threshold was *P* = 10⁻⁴, and genome-wide significance under the QPMM was defined as P < 5 × 10⁻⁸. For each screened-out variant *i*, we calculated the conditional probability that the polynomial statistic would exceed its genome-wide significance threshold given that the GP statistic did not exceed its screening threshold: *qᵢ* = Pr(*T_QP_*,*ᵢ* > *c_QP_* | *T_GP_*,*ᵢ* ≤ *c_GP_*). The marginal and joint probabilities required for this calculation were obtained from the corresponding univariate and bivariate normal cumulative distribution functions. The expected number of genome-wide significant variants missed by GP screening was calculated by summing the variant-specific conditional probabilities: E(*N* ᵢ11) = Σ*ᵢ qᵢ*. This expectation follows from linearity of expectation and therefore does not require independence among variants.

Applying this approximation to all variants excluded by the GP screening threshold yielded an expected number of missed genome-wide significant QPMM associations of 3.8 × 10⁻¹¹, indicating that the expected number of such missed associations was effectively zero. The detailed mathematical derivation is provided in Appendix section 2.3 of **Supplementary Notes 2**.

### Simulating longitudinal BMI data from the estimated parameters of the GP model

We generated a synthetic longitudinal dataset that preserves the main structural features and parameter settings of an existing fitted dataset. It first simulated a pedigree and relatedness structure for *N_d_* individuals using mock kinship coefficients. Longitudinal observation times were then sampled from the empirical distribution of observation schedules in the original dataset, thereby preserving realistic variation in the number and timing of measurements across individuals. Two normally distributed principal components and a binary sex variable were generated as mock covariates.

The GP kernel parameters, including the inducing-point locations and kernel scale parameter, were inherited from the real estimates. The cross-kernel matrix between the simulated ages and inducing points was whitened using the Cholesky decomposition of the inducing-point kernel matrix. Fixed-effect coefficients were then specified using the intercept and GP coefficients estimated from the original dataset.

Family-correlated longitudinal random effects are generated separately for each family. For every individual within a family, the function simulates a random GP component and a random intercept using the variance parameters estimated from the original model. These individual-specific random effects are combined through the pedigree loading matrix derived from the simulated KING relationships. This construction induces correlations among repeated measurements from the same individual and among genetically related individuals.

Using the effect-size coefficients and allele frequencies estimated from the real data, genotype dosages were simulated for the corresponding alternative variants. For each variant, an age-dependent genetic effect is calculated from its coefficient vector and the whitened GP basis, and this effect is multiplied by the individual’s centred dosage. Contributions from all alternative variants are summed to produce a dynamic polygenic component.

The simulated phenotype was generated as the sum of the fixed effects, family-correlated longitudinal random effects, independent residual error, and the simulated age-dependent genetic effects. The residual variance and random-effect variance parameters were inherited from the fitted model for the real data. The complete analysis workflow and accompanying code are available on the DynamicPGS GitHub repository (see **Code Availability**).

### LD pruning and dynamic polygenic score calculation

We computed the LD index between the 590 index variants discovered through the cross-sectional and longitudinal GWAS (11 age strata and the dynamic QTL mapping) using our imputed genotype data. We then converted the 590-dimensional LD matrix *R* into the squared distance matrix *I* − *R*^2^ to perform the hierarchical clustering. Setting the cutoff at 0.9 (*i.e.* LD pruning at *r*^2^ < 0. 1) allowed us to merge loci, resulting in 238 clusters. We selected the index variant with the minimum *P*-value to represent each cluster of loci. Finally, we estimated the dynamic QTL effects at the 238 selected index variants. Here, the posterior distribution of the dynamic QTL effect at each month of child age is given by the GP regression model. Then the individual’s genotype was weighted by the effect size to obtain the polygenic score (PGS) across the child’s age in months. The prediction interval of the PGS was computed using the standard error of the QTL effects across all loci and the estimated residual variance. See the **Supplementary Notes 2** for further details. We have also developed an R package called `DynamicPGS` to predict the dynamic PGS given any genotype data, uploaded to our GitHub page (**Code availability**).

### PGS replication in the Norwegian Mother, Father & Child Cohort study (MoBa)

The Norwegian Mother, Father and Child Cohort Study (MoBa) is a pregnancy-based ongoing cohort study consisting of approximately 114,500 children, 95,200 mothers and 75,000 fathers, enrolled from 50 hospitals across Norway from 1999 to 2008^110^. Children’s height and weight values used to compute BMI in this study were measured by trained nurses during routine checkups at 6 weeks, at 3, 6 and 8 months, and at 1, 1.5, 2, 3, and 5 years of age and reported by parents in questionnaires. Genotyping was conducted from samples donated by participants^111^ by multiple research groups on various genotyping platforms. Genotyping data are managed by the Norwegian Institute of Public Health (NIPH). Information on the different genotyping batches is available from the NIPH (github.com/folkehelseinstituttet/mobagen). This study was conducted on the quality controlled, imputed, and harmonized set of genotypes available from the NIPH version 2025.09.25 (github.com/fhi-beta/mobaGenetics-qc). Only participants clustering with the CEU samples from the HapMap project in a principal component analysis (PCA) were considered (n = 65,455 children). Only variants with a minor allele frequency >0.1% were included.

Out of 238 index variants identified by our data, 200 could be matched to the genotype set in MoBa. Out of the 200, 135 were the same index variant in JECS and 65 were proxy variants in high LD (*r*^2^ > 0. 8) based on our imputed genotype data (**Supplementary Table 8**). Of the 38 non-imputed variants, 12 had no variation in a European population (MAF=0% in 1KGP EUR), 2 had low minor allele frequencies in a European population (MAF < 0.3% in 1KGP EUR), and 24 were either multi-allelic variants, INDELs, or long CNVs that were not included in the Haplotype Reference Consortium (HRC) panel used in MoBa (**Supplementary Table 8**).

Using these 200 variants, the dynamic PGS was used to predict the BMI of children in MoBa at the age strata corresponding to measurements. The variance explained in measured BMI by the PGS was evaluated using a linear model without any covariates. Note that the PGS was computed solely from genotype-weighted age-specific genetic effects and did not include non-genetic components of the GP model.

For comparison with conventional age-specific PGSs, we also constructed cross-sectional PGSs using the JECS GWAS results at each corresponding age stratum. At each age, variants reaching genome-wide significance in the age-stratified JECS BMI GWAS were selected (**Supplementary Table 2**), and the corresponding age-specific effect estimates were used as weights. The number of variants included in these cross-sectional PGSs ranged from 15 to 71 across age strata. The same allele harmonization and variant-availability criteria were applied in MoBa. MoBa data were used only for external evaluation and were not used for variant selection, effect-size estimation, or model tuning. The variance explained by each cross-sectional PGS was evaluated using the same framework as for the dynamic PGS.

The ethics, acknowledgements, and data availability for MoBa are available as **Supplementary Information**.

## Data availability

The manifest file of the Illumina Asian Screening Array (ASA-24v1-0_E2.csv) was available from the product web page (https://support.illumina.com/downloads/infinium-asian-screening-array-v1-0-product-files.html).

The allele frequency data of Tohoku Medical Megabank (ToMMo) samples was obtained from jMorp (https://jmorp.megabank.tohoku.ac.jp/). The latest GWAS results were downloaded from the GWAS Catalog (https://www.ebi.ac.uk/gwas/api/search/downloads/gwas_catalog_v1.0-associations_e113_r2024-11-03.tsv). Two imputation reference panels were obtained from the 1KGP (ALL.chr*.shapeit2_integrated_snvindels_v2a_27022019.GRCh38.phased.vcf.gz and 1kGP_high_coverage_Illumina.chr*.filtered.SNV_INDEL_SV_phased_panel.vcf.gz). The variant VCF of a South Korean population was obtained from the Korean Personal Genome Project (KPGP) (http://kpgp.kr/). The pre-computed LD scores using the 1000 Genomes Project (1KGP) East Asian Population (1000G_Phase3_EAS_baselineLD_v2.2_ldscores.tgz) was downloaded from ZENODO (https://zenodo.org/records/10515792). GTEx eQTL summary statistics of the skeletal muscle tissue (V8) were downloaded from the eQTL Catalogue (ftp.ebi.ac.uk/pub/databases/spot/eQTL/sumstats/QTS000015/QTD000121/QTD000121.all.tsv.gz).

Ensembl gene annotation (version 113) was downloaded from BioMart (https://www.ensembl.org/). GWAS results of childhood BMI from MoBa were downloaded from the NIPH website (https://www.fhi.no/en/ch/studies/moba/for-forskere-artikler/gwas-data-from-moba/). Raw genotype data for child participants in JECS have not been uploaded to a publicly accessible repository due to parental consent, ethical restrictions, and legal framework of the Act on the Protection of Personal Information (Act No. 57 of 30 May 2003, amendment on 1 June 2025). Access to the raw data can be granted for scientific purposes only at the JECS facilities located in Japan. All inquiries about data access should be sent to. The person responsible for handling enquiries sent to this e-mail address is Dr Shoji F. Nakayama, JECS Programme Office, National Institute for Environmental Studies. The GWAS summary statistics for all 1,255 traits are available via the JECS PheWeb (https://jecs-pheweb.ncchd.go.jp/).

## Code availability

The fixref plugin of bcftools to align strand orientations of SNP probes is available online (https://samtools.github.io/bcftools/howtos/plugin.fixref.html). BLAST+ was downloaded from the NCBI FTP site (https://ftp.ncbi.nlm.nih.gov/blast/executables/blast+/). KING (version 2.3.2) was used to estimate kinship between the genotyped samples in JECS (https://www.kingrelatedness.com/). Beagle (beagle.22Jul22.46e.jar) was used for imputation of the South Korean samples and Minimac4 (v4.1.6) was used for imputation of the genotyped samples in JECS (https://faculty.washington.edu/browning/beagle/beagle.html). ADMIXTURE (v1.3.0) was used to estimate mixed ancestries in our participants (https://dalexander.github.io/admixture/index.html). EAGLE software (v2.4.1) was used to pre-phase the genotype data prior to the imputation (https://alkesgroup.broadinstitute.org/Eagle/). SHAPEIT5 was used to perform haplotype phasing prior to call chromosomal mosaicisms. (https://github.com/odelaneau/shapeit5). MoChA (version 2024-09-27) was used to detect mosaic chromosomal alterations (https://github.com/freeseek/mocha/). PLINK2 (v2.00a5.10LM 64-bit Intel) was used to perform PCA and genotype data preparations. REGENIE (ver 3.4.1) was used to perform GWAS (https://rgcgithub.github.io/regenie/). ANNOVAR (2020-06-07) was obtained online (https://annovar.openbioinformatics.org/en/latest/). LDSC (v1.0.1) was used to estimate the trait (additive and SNP-based) heritability and genetic correlations between traits (https://github.com/bulik/ldsc). The DynamicPGS R package provides executable code for dynamic genetic association mapping and PGS computation available on the GitHub page (https://github.com/natsuhiko/DynamicPGS). DEEP*HLA was used to perform HLA imputation (https://github.com/tatsuhikonaito/DEEP-HLA). METAL (version released on 2011-03-25) was used to perform simple meta-analysis of BMI GWAS across 11 distinct age strata (https://csg.sph.umich.edu/abecasis/metal/download/).

---

**Extended Data Figure 1.**
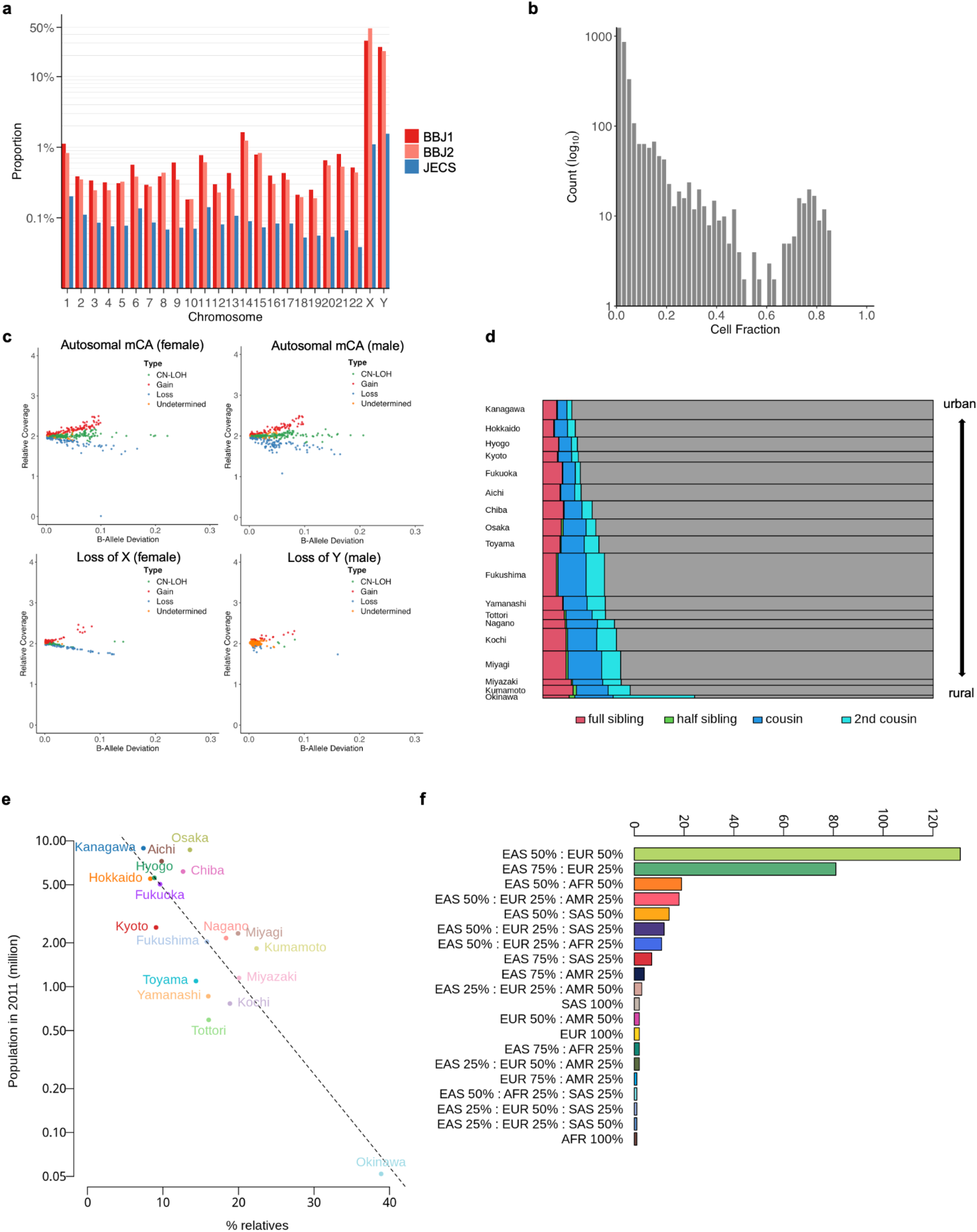
**a**. Barplot shows the proportion of mosaic chromosomal alterations (mCAs) detected in each chromosome across the BBJ1 (red), BBJ2 (orange) cohorts, and JECS (blue). The y-axis is shown on a logarithmic scale. **b.** Histogram shows the distribution of cell fractions among all detected mCA events. The y-axis is plotted on a logarithmic scale. **c.** Scatter plots show the distribution of mosaic chromosomal alterations by B-allele deviation (x-axis) and relative coverage (y-axis) in autosomes and sex chromosomes. Each dot represents a single event, color-coded by event type: CN-LOH (green), Gain (red), Loss (blue), and Undetermined (orange). (Top left) Autosomal mCAs in females. (Top right) Autosomal mCAs in males. (Bottom left) Loss of X in females. (Bottom right) Loss of Y in males. **d.** Mosaicplot shows the percentage of relatives in our data, stratified by the prefectures of Japan where participants were enrolled. **e.** Scatterplot shows the relationship between the percentage of relatives (full/half siblings, cousins and second cousins) and the population (million people) of each prefecture in 2011 obtained from e-Stat in Japan (https://www.e-stat.go.jp/). Note that the population of Okinawa (bottom right point) reflects the number of people in Miyakojima City (but not the total population of Okinawa Prefecture), where all pregnant mothers in the Okinawa area were enrolled. The dotted line shows a linear regression line with an adjusted R-square value of 0.72 and a P-value of 4. 7 × 10^−6^ . Even if we remove the data from Okinawa, the R-squared value is 0.43 with a P-value of 0.0026. **f**. Barplot shows the number of non-East Asian participants with different ancestral backgrounds inferred from the principal component analysis.

**Extended Data Figure 2.**
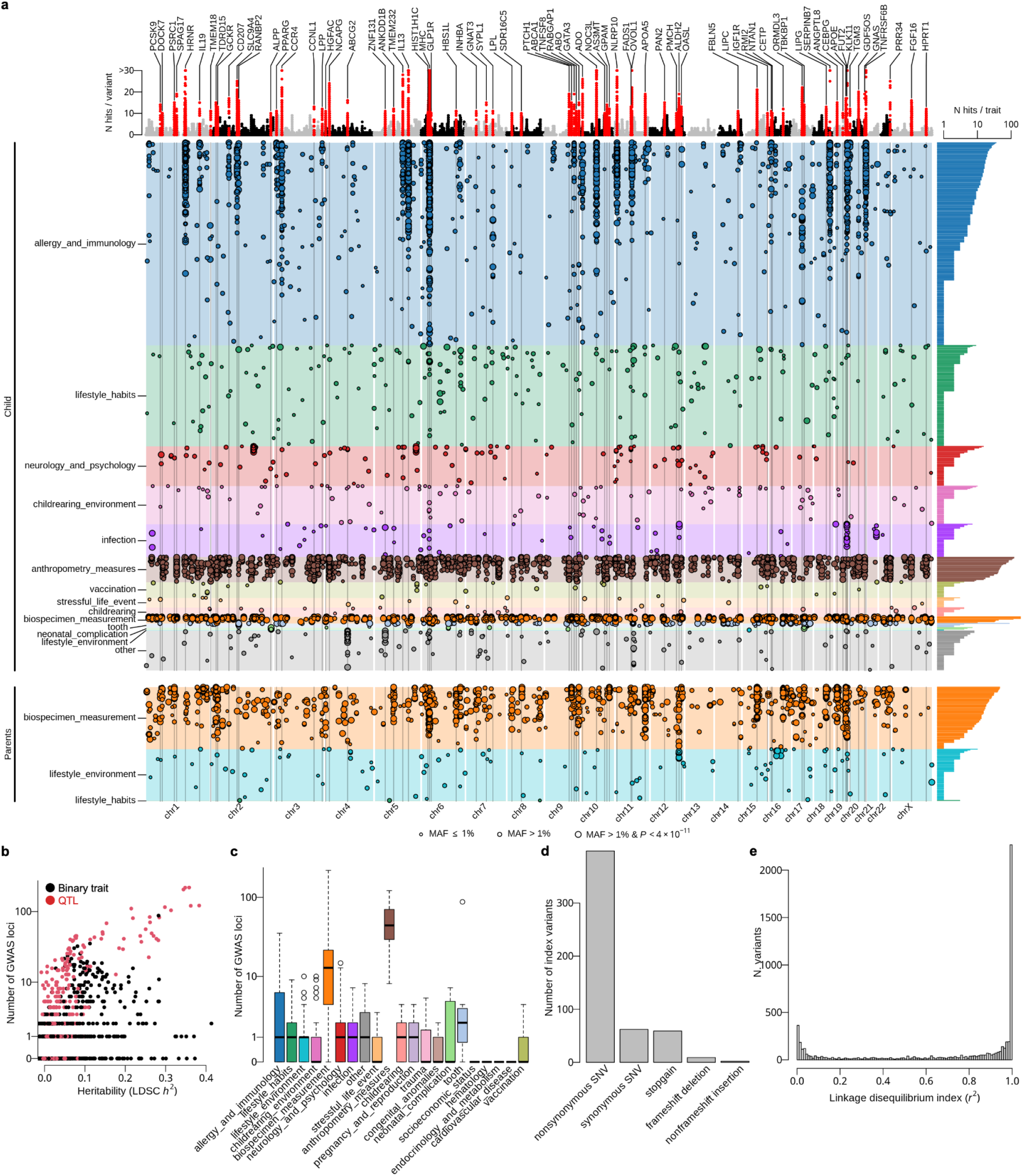
**a**. Genetic associations discovered through GWAS of 1,255 traits in JECS. The top panel is a Manhattan-like plot showing the number of GWAS hits (*P* < 5. 0 × 10^−8^) at each variant with the nearest gene annotation. Peaks are defined by variants shared across 10 or more traits. The main panel shows locations of index variants for each trait segregated by the trait category. The size of each dot indicates the magnitude of the association as well as the minor allele frequency (MAF) of the respective index variant. The largest dot indicates an index variant that passed the phenome-wide threshold of *P* < 4. 0 × 10^−11^, regardless of its MAF. If an index variant does not pass the phenome-wide threshold, it is classified by its MAF greater than 1% (mid-size dot) or less than 1% (smallest dot). The right panel shows a barplot of the number of GWAS hits for each trait. **b.** Scatterplot shows the number of GWAS loci for each trait as a function of the additive SNP heritability estimation obtained by the LD Score regression (LDSC). **c.** Boxplot shows the number of significant GWAS loci (*P* < 5. 0 × 10^−8^) for each trait stratified by the trait categories. **d.** The number of index variants detected within the coding regions, stratified by the putative functions of variants annotated by the Ensembl gene (version 113; GRCh38). **e.** The histogram shows the distribution of linkage disequilibrium index (*r*^2^) between the GWAS index variants in our study and the entire variants reported in the GWAS catalog. We took the maximum *r*^2^ value for each of our index variants.

**Extended Data Figure 3.**
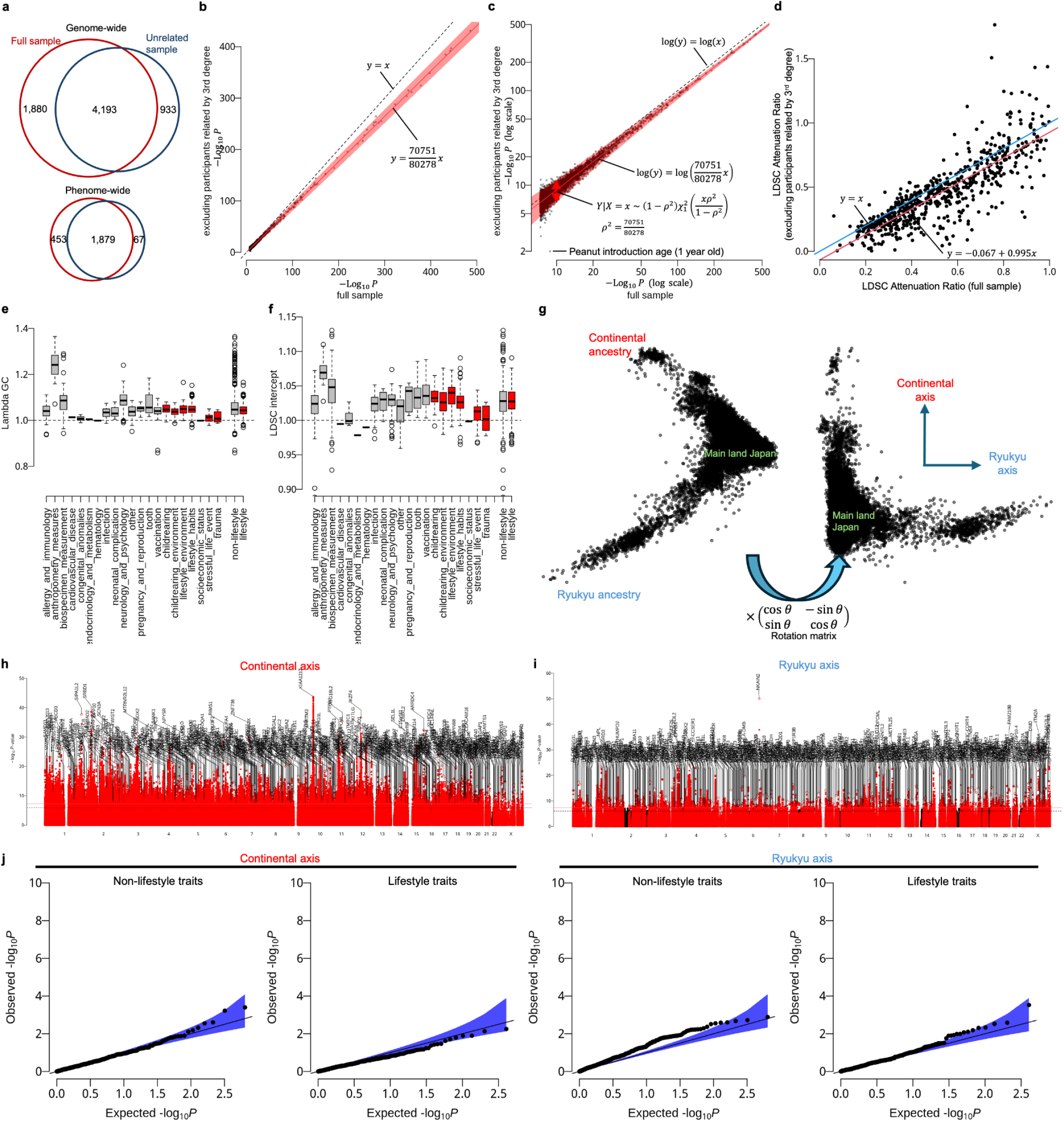
**a**. The number of GWAS loci shared by the full sample (*N*=80,278) and the unrelated sample (*N*=70,751), excluding third degree relatives. The top Venn diagram shows loci passed the standard GWAS threshold (*P* < 5. 0 × 10^−8^) and the bottom Venn diagram shows loci passed the phenome-wide threshold (*P* < 4. 0 × 10^−11^). **b.** Scatter plot of P-values for the 6,073 associations identified in the full-sample GWAS compared with those from the GWAS of unrelated individuals. Association signals were evaluated at the index variants identified in the full-sample GWAS. The dashed line indicates the diagonal (*y* = *x*), and the red line represents the expected reduction in statistical power when subsampling 70,751 individuals from the full sample of 80,278. The expected slope is given by 70,751/80,278, corresponding to the conditional mean of a non-central chi-square distribution. The red shaded area denotes the 95% confidence interval, also derived from the non-central chi-square distribution. See Methods for details. **c.** Same as in panel b, but with both axes shown on a logarithmic scale. The dashed line, red line, and shaded area are identical to those in panel b. **d.** Scatter plot of LDSC attenuation ratios for traits with heritability Z-score > 0 and at least one GWAS hit. The x-axis shows attenuation ratios from the full-sample GWAS, and the y-axis shows those from the GWAS of unrelated individuals. The blue line indicates the diagonal (*y* = *x*), and the red line represents the least-squares fit (*y* = *a* + *bx*). The negative intercept (*a* = −0.067) reflects a modest reduction in attenuation ratio after excluding individuals related up to the third degree. **e.** Boxplot shows the distribution of genomic control lambda (λ*_GC_*) for the 1,255 traits analyzed, stratified by trait categories (red boxes indicate the lifestyle trait categories). The last two boxes compare the distribution of aggregated non-lifestyle and lifestyle traits, in which the lifestyle traits combine *childrearing*, *childrearing environment*, *lifestyle environment*, *lifestyle habits*, *socioeconomic status*, *stressful life events*, and *trauma*. **f.** Boxplot shows the distribution of the LDSC intercept for the 1,255 traits analyzed, stratified by trait categories. **g.** The 2-dimensional rotation approach to extract the continental and Ryukyu axes. **h.** Manhattan plot shows GWAS of the continental axis extracted in the panel g. The continental axis is analyzed as a QTL trait. **i.** Manhattan plot shows GWAS of the Ryukyu axis extracted in the panel g. The Ryukyu axis is analyzed as a QTL trait. **j.** QQ plots of cross-trait LD score regression (LDSC) *P*-values between ancestry axes and all analyzed traits, stratified into lifestyle and non-lifestyle categories. No traits passed the trait-wide significance threshold (*P* < 0. 05/1255).

**Extended Data Figure 4.**
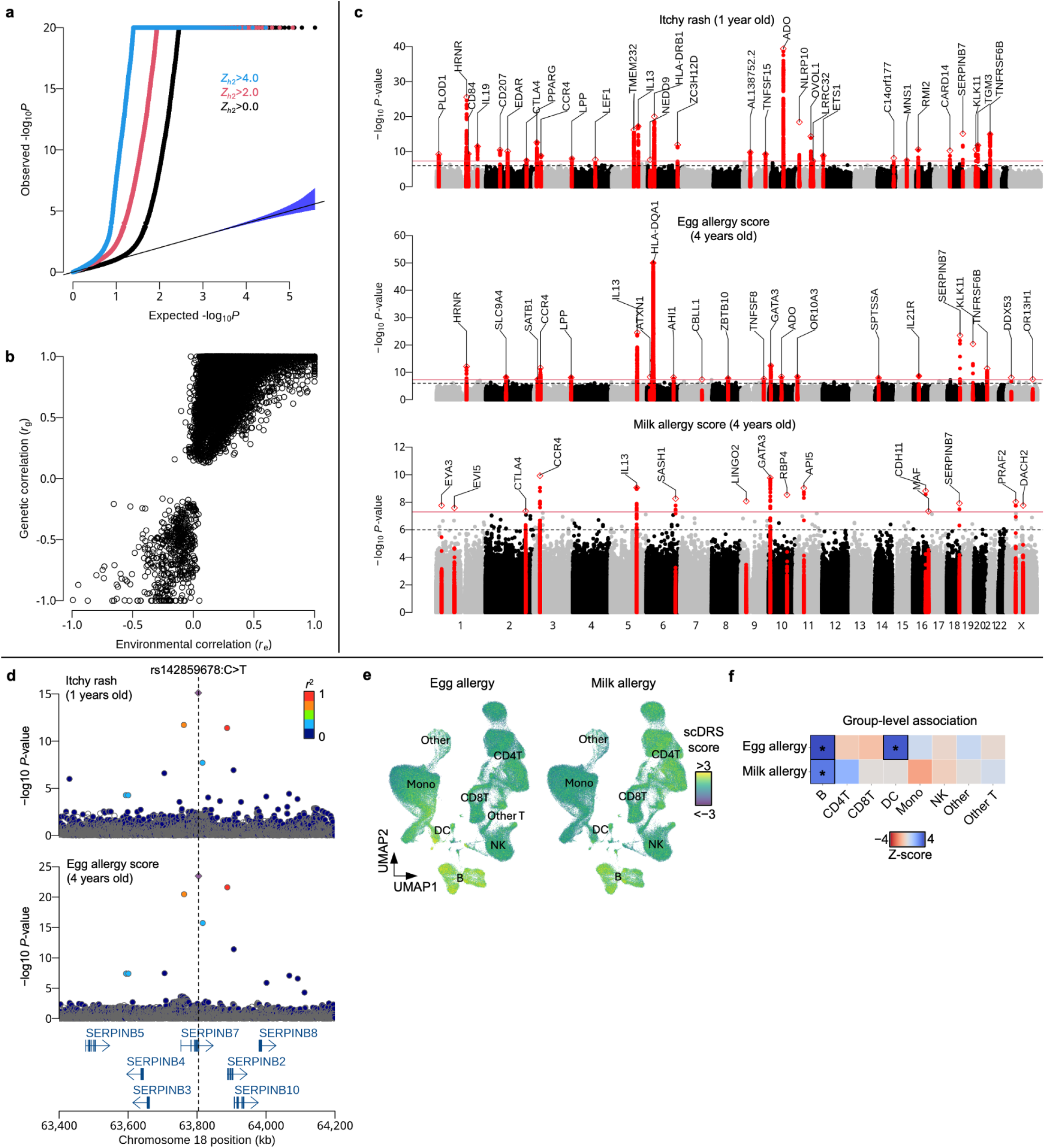
**a**. QQ-plot shows *P*-values of genetic correlations with different cutoffs for heritability Z-score (black: *Z*_ℎ_2 > 0; red *Z*_ℎ_2 > 2. 0; blue: *Z*_ℎ_2 > 4. 0) **b.** Scatterplot shows the relationship between the environmental correlation (x-axis) and the genetic correlation (y-axis) (**Methods**). **c.** Manhattan plots show the GWAS results of the itchy rash status at 1 year old (top), the egg allergy cumulative status until 4 years old (middle) and the milk allergy cumulative status until 4 years old (bottom). **d.** The Locuszoom plots show the GWAS associations around the SERPINB7 gene. The top panel shows the result for itchy rash at 1 year old and the bottom panel shows the result for egg allergy (both data extracted from the panel c of this figure). The location of the index variant (rs142859678:C>T), which is known to be the pathogenic variant of NPPK (Nagashima-Type Palmoplantar Keratosis), is indicated by a dashed line. **e.** UMAP plots showing scDRS^55^ score distributions for cumulative egg and milk allergy scores up to 4 years of age in the reference PBMC data.^56^ **f**. Heatmap showing cell type-level scDRS association Z-scores for cumulative egg and milk allergy scores up to 4 years of age. Asterisks indicate significant enrichment after correction for multiple testing across the tested cell types (FDR < 0.05).

**Extended Data Figure 5.**
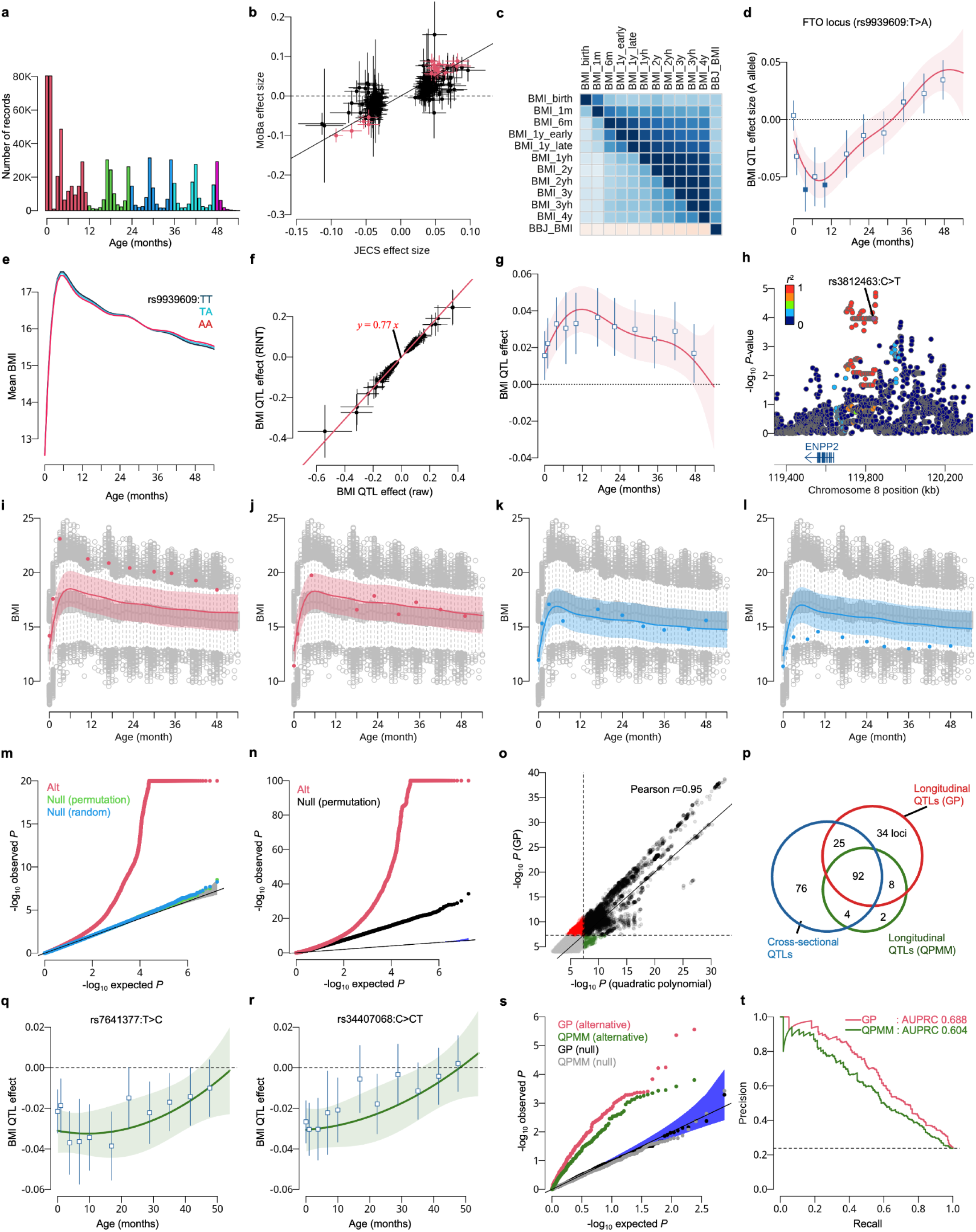
**a**. The histogram shows the number of samples along the child’s age in months (top) and the box plot shows the distribution of BMI along the child’s age in months (bottom). The color indicates the child’s age in years (red: 0 years old; green: 1 year old; blue: 2 years old; light blue: 3 years old; purple: 4 years old). **b.** The scatter plot compares QTL effect sizes between JECS and MoBa at variants discovered by the JECS GWAS of 11 different strata. The crossed lines indicate the 95% confidence intervals, which were estimated using standard error estimates. The red color indicates the variant was significant (*P* < 5. 0 × 10^−8^) in both JECS and MoBa. **c.** Heatmap shows genetic correlations (upper triangular matrix) and environmental correlations (lower triangular matrix) of BMI GWAS at the 11 time points along with BMI in adult (obtained from BioBank Japan^32^) estimated by the cross-trait LD score regression. The color coding is the same as in Fig. 4 in the main text. **d.** The panel shows the estimated dynamic genetic association at the FTO locus, where the mean effect size of T allele relative to A allele at rs9939609 is shown by the red line with the 95% credible interval. Cross-sectional QTL effect estimates (using the raw BMI) and their 95% confidence intervals across 11 age strata are overlaid as blue squares and error bars (filled squares denote the *P*-values < 5. 0 × 10^−8^). **e.** The panel shows the mean BMI along the child’s age in months, stratified by the three genotype groups (TT, TA and AA) at rs9939609. **f.** Scatterplot shows the cross-sectional GWAS effect sizes and their 95% confidence intervals for raw BMI (x-axis) and the RINT-transformed BMI (y-axis). The linear regression line in red shows the scale of the raw and RINT-transformed BMI to be 0.77. **g.** Age-dependent genetic effect of the T allele relative to the C allele at rs3812463, estimated by Gaussian process regression. The red line and shaded area indicate the posterior mean and 95% credible interval, respectively. Cross-sectional QTL effect estimates (using the RINT BMI; **Supplementary Table 2**) times 1.3 (=1/0.77) and their 95% confidence intervals across 11 age strata are overlaid as blue open squares and error bars. **h.** The locus zoom plot shows the ENPP2 eQTL in skeletal muscle tissue. The summary statistics were obtained from the eQTL Catalogue which is based on GTEx V8 (**Data availability**). The color of each point indicates the LD index (*r*^2^) to the TAF2 index variant (rs3812463:C>T). The LD index between the TAF2 index variant (rs3812463:C>T) and the eQTL lead variant (rs1433953:T>C) was 0.97. **i-l**. The lines show the estimated dynamic polygenic scores for four different participants, with dots indicating the actual observations for each participant (including the age-dependent non-genetic population-average trajectory for visualization purposes). Light color bands around the estimated polygenic scores show the prediction intervals. The color indicates whether the participant is underweight (blue) or overweight (red) compared to the population average, as shown by the boxplot. **m.** QQ-plot of P-values under the null (green and blue) and the alternative (red) hypotheses. The genotype under the two null hypotheses was obtained either (1) by permuting the real genotype data (green) or (2) by using a Gaussian copula based on a family-wise Cholesky representation of the additive relationship matrix constructed from KING relatedness estimates (blue) (**Methods**). We obtained the *P*-values from the alternative hypothesis (red) through the dynamic genetic mapping (shown in Fig. 5b of the main text). **n.** QQ-plot of P-values for the age-stratified meta-analysis (Methods). The red dots show the inverse-variance weighted P-values under alternative hypothesis and the black dots show those under the null hypothesis with permuted phenotypes (black). The genomic control lambda was 2.7 for the null hypothesis. **o.** Scatterplot shows the P-values of the GP regression model (shown in Fig. 5b) against the quadratic polynomial mixed-effect model (QPMM) for the 63,640 variants identified by GP-based screening at P < 10⁻⁴. The dashed lines show the genome-wide significance threshold ( *P* = 5. 0 × 10^−8^). Green dots show the variants only discovered through QPMM. Red dots show the variants only discovered through the GP approach. Black dots show the variants discovered through both approaches. **p.** Venn diagram shows the genetic loci (lead variants whose LD *r*^2^ < 0.1) discovered through cross-sectional GWAS (blue), GP-based longitudinal GWAS (red) and the QPMM-based longitudinal GWAS (green). **q-r**. Age-dependent genetic effect of the T allele relative to the C allele at rs7641377 (panel q) and the C allele relative to the CT allele at rs34407068 (panel r), estimated by QPMM. The green line and shaded area indicate the fitted quadratic function of child age as the posterior mean and 95% credible interval, respectively. Cross-sectional QTL effect estimates (using the raw BMI) and their 95% confidence intervals across 11 age strata are overlaid as blue open squares and error bars. **s.** QQ-plot of association *P* values obtained from the mock-data analysis. Red and green points represent the 238 variants simulated under the alternative hypothesis analyzed using the GP and QPMM approaches, respectively, whereas black and gray points represent the 762 variants simulated under the null hypothesis analyzed using the GP and QPMM approaches, respectively. **t.** Precision–recall curves for the GP and QPMM approaches based on association *P* values from the mock-data analysis. The area under the precision–recall curve (AUPRC) was 0.688 for GP and 0.604 for QPMM. The dashed horizontal line indicates the expected precision for random classification based on the proportion of variants simulated under the alternative hypothesis (238/1,000 = 0.238).

**Extended Data Figure 6.**
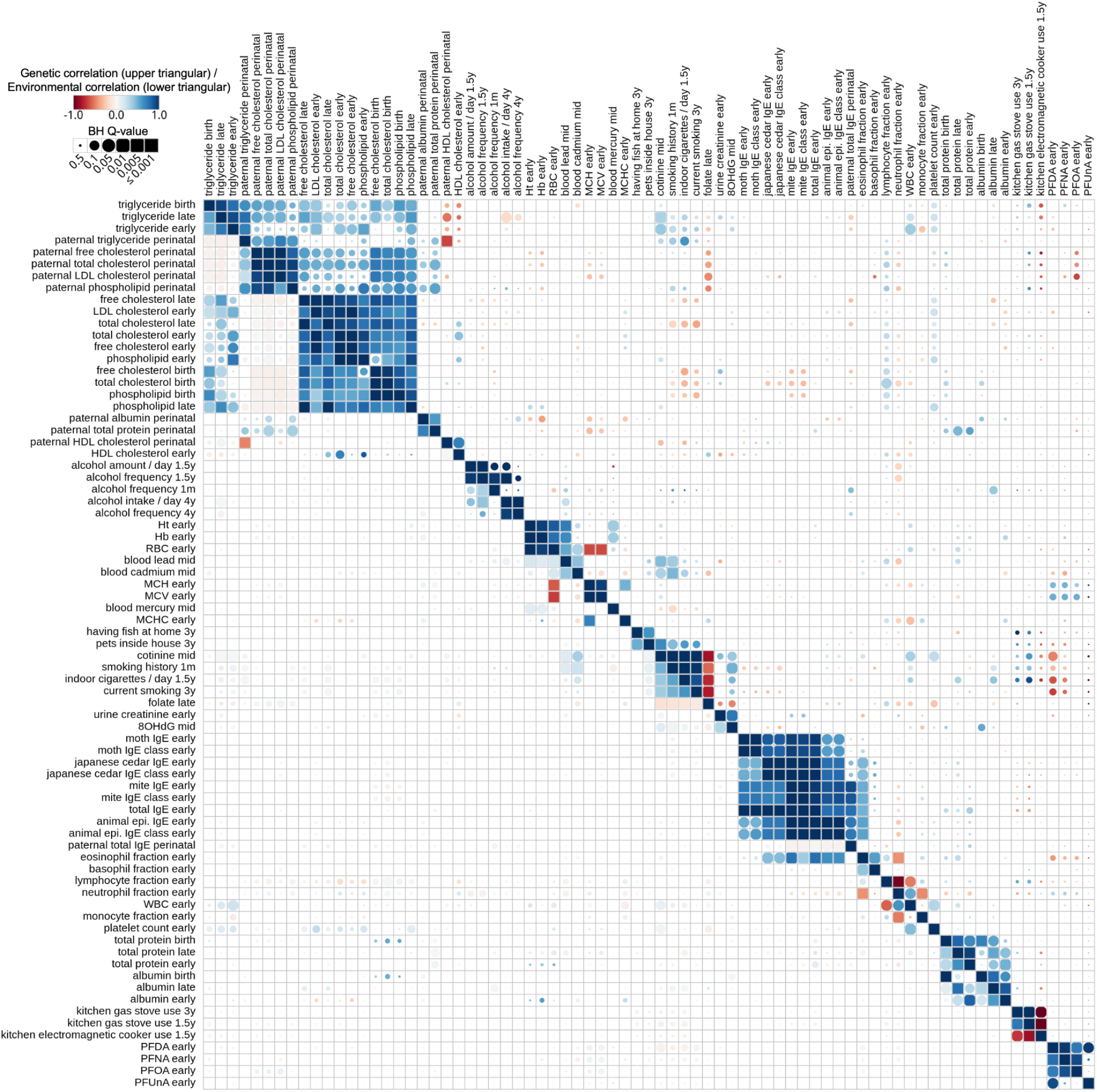
Cross-trait LDSC results for parent exposure traits. The upper triangular matrix shows the genetic correlations (*r_g_*) and the lower triangular matrix shows the environmental correlations (*r_e_*). Blue colors indicate positive correlations and red colors indicate negative correlations.

**JECS Genetics Research Group**

**JECS Genetics Research Working Group**

Natsuhiko Kumasaka, Tatsuhiko Naito, Hidetoshi Mezawa, Kei Sakamoto, Kenji Matsumoto, Kazuhiko Nakabayashi, Masayo Kagami, Emiko Noguchi, Akira Hata, Kenichiro Hata, Yukinori Okada, Atsushi Tajima, Yukihiro Ohya

**JECS Group**

Reiko Kishi, Chiharu Ota, Koichi Hashimoto, Chisato Mori, Shuichi Ito, Ryoji Shinohara, Hidekuni Inadera, Takeo Nakayama, Ryo Kawasaki, Yasuhiro Takeshima, Seiji Kageyama, Akemi Morita, Narufumi Suganuma, Shouichi Ohga, Takahiko Katoh, Michihiro Kamijima, Maki Fukami, Shin Yamazaki

**JECS Programme Office**

Tomohiko Isobe, Makiko Sekiyama, Miyuki Iwai-Shimada, Yayoi Kobayashi, Mai Takagi, Nozomi Tatsuta, Yu Taniguchi, Shoji F. Nakayama, Shin Yamazaki

**JECS Medical Support Center**

Natsuhiko Kumasaka, Tatsuhiko Naito, Hidetoshi Mezawa, Kei Sakamoto, Kazuhisa Akiba, Atsushi Hattori, Miori Sato, Mayako Saito-Abe, Aya Yoshikawa, Limin Yang, Daisuke Harama, Yumiko Miyaji, Minaho Nishizato, Kiwako Yamamoto-Hanada, Akihiro Umezawa, Maki Fukami

**BioBank Japan**

Natsuhiko Kumasaka, Shi Mingyang, Masaru Koido, Go Sato, Tatsuhiko Naito, Yukinori Okada, Yoichiro Kamatani, Koichi Matsuda

**National Center Biobank Network**

Yosuke Kawai, Katsushi Tokunaga

**Norwegian Mother, Father, and Child Cohort Study (MoBa)** Mikko Baur, Nicolas Fragoso-Bargas, Marc Vaudel, Stefan Johansson

## Contributions

All authors reviewed and revised the manuscript. **Study design:** R.Kishi, C.O., K.Hashimoto, C.M., S.I., R.S., H.I., T.Nakayama, R.Kawasaki, Y.Takeshima, S.K., A.M., N.S., S.H., T.K., M.Kamijima, T.I., M.Sekiyama, M.I-S., Y.K., M.T., N.T., Y.Taniguchi, S.F.N., S.Y., N.K., T.N., H.M., M.Sato, M.S-A, A.Y., L.Y., D.H., Y.M., M.N., K.Y-H., M.F. and Y.Ohya. **Sample collection:** R.Kishi, C.O., K.Hashimoto, C.M., S.I., R.S., H.I., T.Nakayama, R.Kawasaki, Y.Takeshima, S.K., A.M., N.S., S.H., T.K. and M.Kamijima. **Phenotype data curation:** N.K., T.N., H.M., K.S., T.I., M.Sekiyama, M.I-S., Y.K., M.T., N.T., Y.Taniguchi, S.F.N. and S.Y. **Sample handling and genotyping:** T.I. and S.F.N. **Genotype QC:** N.K. and T.N. **Conceptualization:** N.K., T.N., H.M., K.S., K.Matsumoto, K.N., M.Kagami, E.N., A.Hata, K.Hata, Y.O., A.T., S.J., S.F.N. and Y.Ohya. **External data provision:** S.M., M.Koido, Y.Kamatani., T.Naito, G.S., Y.O., K.Matsuda, BioBank Japan, Y.Kawai, K.T., National Center Biobank Network, M.B., N.F-B, M.V. and S.J. **External data validation and interpretation in MoBa cohort:** M.B., N.K., N.F-B, M.V. and S.J. **Methodology:** N.K. and T.N. **Analysis:** N.K., T.N., M.B., K.S. and K.A. **Writing original draft:** N.K., T.N., H.M., M.B., K.S., K.A., A.Hattori, M.V., S.J. and Y.O. **Ethics and Governance:** H.M., S.F.N., R.Kishi, C.O., K.Hashimoto, C.M., S.I., R.S., H.I., T.Nakayama, R.Kawasaki, Y.Takeshima, S.K., A.M., N.S., S.H., T.K. and M.Kamijima. **Supervision:** K.Y-H., A.U., M.F., S.Y., Y.O., S.F.N. and Y.Ohya.

## Notes

### Competing Interest Statement

The authors have declared no competing interest.

### Author Declarations

The Japan Environment and Children's Study (JECS) protocol was reviewed and approved by the Ministry of the Environment's Institutional Review Board on Epidemiological Studies (No.100910001) and the Ethics Committees of all participating institutions. Written informed consent was obtained from all children's legal guardians. Following informed consent, participants were given further information about SNP genotyping using microarray technology and the possibility to withdraw from the genetic study at any time.

### Summary of Updates

Updated longitudinal GWAS of BMI.

