## Supplementary Notes 1 for "Genome-wide association study on longitudinal and cross-sectional traits of child health and development in a Japanese population"

### JECS Phenotype Data Management for Children up to 4 Years of Age and Parental Exposures in the Flagship Paper

Version 5.0.2

Date: 2026/04/09

**1. Introduction** **3**
**2. Variable selection workflow** **3**
**3. Quality controls and transformation** **6**
3.1. Conditioning on other variable(s) 7
3.2 Outlier detection 7
3.3 Replacement 8
3.4 Offset of binary questionnaires 8
3.5 Masking responses other than those from the mother 8
**4. Composite traits** **9**
4.1 Categorical variables with multiple choices 9
4.2 Binarization of numerical variables 9
4.3 Complex composite traits from multiple questionnaire variables 9
BMI traits 10
Early pregnancy HbA1c (NGSP) 10
Perinatal-related traits 10
Breastfeeding-related traits 11
First vaccination-related traits 11
Food allergy-related traits 12
Feeding status-related traits 16
Rhinitis-related traits 17
Medical history-related traits 18
Eczema-related trait 20
Sleep-related traits 20
Childcare facility use-related traits 21
Toilet training-related trait 22
ASQ-3 questionnaire-related traits 22
**5. Trait selection for GWAS** **22**

### 1. Introduction

The Japan Environment and Children's Study (JECS) is a large-scale birth cohort study funded by Japan's Ministry of the Environment to evaluate the effects of environmental chemicals on children's health and development. More than 100,000 pregnant women were enrolled at 15 regional centers across Japan, representing the genetic diversity of the Japanese population. Since the participant mothers were pregnant, detailed data from questionnaires, biological and physical measurements have been collected from both parents and their children, with additional surveys conducted on average every six months for 80% of the child participants.

This document provides information on phenotype data management for participating children up to four years old and parental exposures obtained during that period. This document is intended to:

- provide an overview of the variable selection workflow (Section 2);
- describe the quality control criteria and appropriate transformations (Section 3);
- provide a detailed explanation of creating composite traits (Section 4);
- provide a detailed explanation of trait selection (Section 5).

Before we dive into the technical material, we will clearly distinguish the terms "variable" and "trait" to explicitly state the process of phenotype data creation. The term "variable" refers to any column in a data sheet or data matrix originally obtained by the JECS programme office. This includes answers to biannual survey questions and physical/biological measurements obtained by doctors or from biological samples. The term "trait," on the other hand, is more abstract and refers to any observable feature of a human being that is potentially heritable from parents to offspring. Therefore, some variables can be analyzed as traits (e.g., an episode of disease during a specific time period), while others are not analyzable until they are combined and transformed into an appropriate trait (e.g., sleep duration based on bedtime and wake-up time). In the next section, we start by selecting the variables that will be used to create the traits for the subsequent genome-wide association studies (GWAS) in the main text.

### 2. Variable selection workflow

All analyses were conducted using the frozen datasets **jecs-ta-20190930** and **jecs-qa-20210401**, obtained from the JECS Programme Office. We selected 23 data sheets containing information on child outcomes and parental exposures (**Table 1**). To construct the phenotype dataset for GWAS, we extracted a total of 4,460 variables from questionnaire, biological, and physical measurements collected from participating children up to 4 years of age and their parents (**Figure 1**). Note that, the actual questions and their corresponding options for the 4,460 variables are listed in **Supplementary Table 6** of the flagship paper.

Each variable was then subjectively classified as either *usable* or *non-usable* according to the meaning of the questionnaire (**Figure 1**). A total of 2,190 invalid variables were excluded and given the appropriate *out-of-use* flags (**Table 2**). Note that the data sheets contained not only child outcomes, but also parental outcomes (e.g., mental health conditions after birth, as measured by the PSI questionnaire). These outcomes were outside the scope of the current manuscript, because the complete parental genotype information was unavailable at the submission stage. On the other hand, we have decided to analyze parental exposure to various environmental factors (including heavy metals and PFAS), as these factors are of interest because they may affect child outcomes. These exposure traits were analyzed by the GWAX-by-proxy approach, where each trait was regressed on child genotypes. We understand the approach is limited in power and interpretation as noted in the flagship paper.

**Table 1**: The 23 data sheets obtained from the JECS programme office

| ID | Sheet_type | Sheet_file | Description |
|---|---|---|---|
| 1 | Questionnaires about children | ageof03_datadr0m_ver002.csv | Medical record transcriptions at birth |
| 2 | Questionnaires about children | ageof03_datadr1m_ver001.csv | Medical record transcriptions at age 1 month |
| 3 | Questionnaires about children | ageof03_datam1m_ver001 | Parent-reported questionnaires at age 1 month |
| 4 | Questionnaires about children | ageof03_datac6m_ver001.csv | Parent-reported questionnaires at age 6 months |
| 5 | Questionnaires about children | ageof03_datac1y_ver001.csv | Parent-reported questionnaires at age 12 months |
| 6 | Questionnaires about children | ageof03_datac1hy_ver001.csv | Parent-reported questionnaires at age 18 months |
| 7 | Questionnaires about children | ageof03_datac2y_ver002.csv | Parent-reported questionnaires at age 24 months |
| 8 | Questionnaires about children | ageof03_datac2hy_ver001.csv | Parent-reported questionnaires at age 30 months |
| 9 | Questionnaires about children | ageof03_datac3y_ver001.csv | Parent-reported questionnaires at age 36 months |
| 10 | Questionnaires about children | ageof04_datac3hy_ver007.csv | Parent-reported questionnaires at age 42 months |
| 11 | Questionnaires about children | ageof04_datac4y_ver008.csv | Parent-reported questionnaires at age 48 months |
| 12 | Questionnaires about children | ageof03_datatsh_ver001.csv | Newborn TSH measurements (dried blood spot) |
| 13 | Questionnaires about children | ageof03_asq_ver001.csv | Ages and Stages Questionnaire scores through age 48 months |
| 14 | Exposure measurements for parents | 03-data-mtlmt2_ver001.csv | Maternal blood heavy metal measurements in mid-pregnancy |
| 15 | Exposure measurements for parents | 03-data-ctnmt2_ver003.csv | Maternal urine cotinine measurements in mid-pregnancy |
| 16 | Exposure measurements for parents | 03-data-pfas_ver002.csv | Maternal blood PFAS measurements during pregnancy |
| 17 | Exposure measurements for parents | ageof03_bioall_ver001.csv | Blood and urine test results during the pregnancy and around delivery in mothers, fathers, and children |
| 18 | Exposure measurements for parents | ageof04_psi_ver006.csv | Parenting Stress Index scores at age 18, 30, 42 months |
| 19 | Additional information | ageof03_add001_ver001.csv | Program office-validated anthropometric measurements through age 36 months |
| 20 | Additional information | ageof03_datacal_ver001.csv | Program office-generated status indicators and composite variables through age 36 months |
| 21 | Additional information | ageof04_datacal_ver005.csv | Additional program office-generated status indicators and composite variables at age 42 and 48 months |
| 22 | Additional information | ageof03_dataagree_ver001.csv | Consent status and withdrawal status through age 36 months |
| 23 | Additional information | ageof04_dataagree_ver002.csv | Consent status and withdrawal status at age 42 and 48 |

After selecting the usable variables, they underwent quality control (**Figure 1**) to remove outliers and transformed into a proper format (e.g., disease cases are assigned to 1 and healthy controls are assigned to 0 for binary traits). For some variables, other variables were used only for QC. For example, maternal heavy metal exposure variables were left-censored by their QC variables. **Section 3** will describe the details of QC filters and transformations.

We then selected 24 variables from 2,183 analysis variables, which served as metadata for our participants and were potentially covariates rather than phenotypes (**Supplementary Table 6**). Note that the variables are not mutually exclusive in the workflow (**Figure 1**). For example, gestational week is used as a covariate to adjust for child development in various traits and is also analyzed as a trait in the GWAS of premature birth. Therefore, the total number of *trait candidate variables* and *covariate source variables* is not equal to the number of *analysis variables*. Additionally, not all covariate source variables were used as covariates in the analysis because some were duplicates (e.g., age of parents in different questionnaires) or were substituted by other covariates (e.g., the birthplace was substituted by genotype principal components).

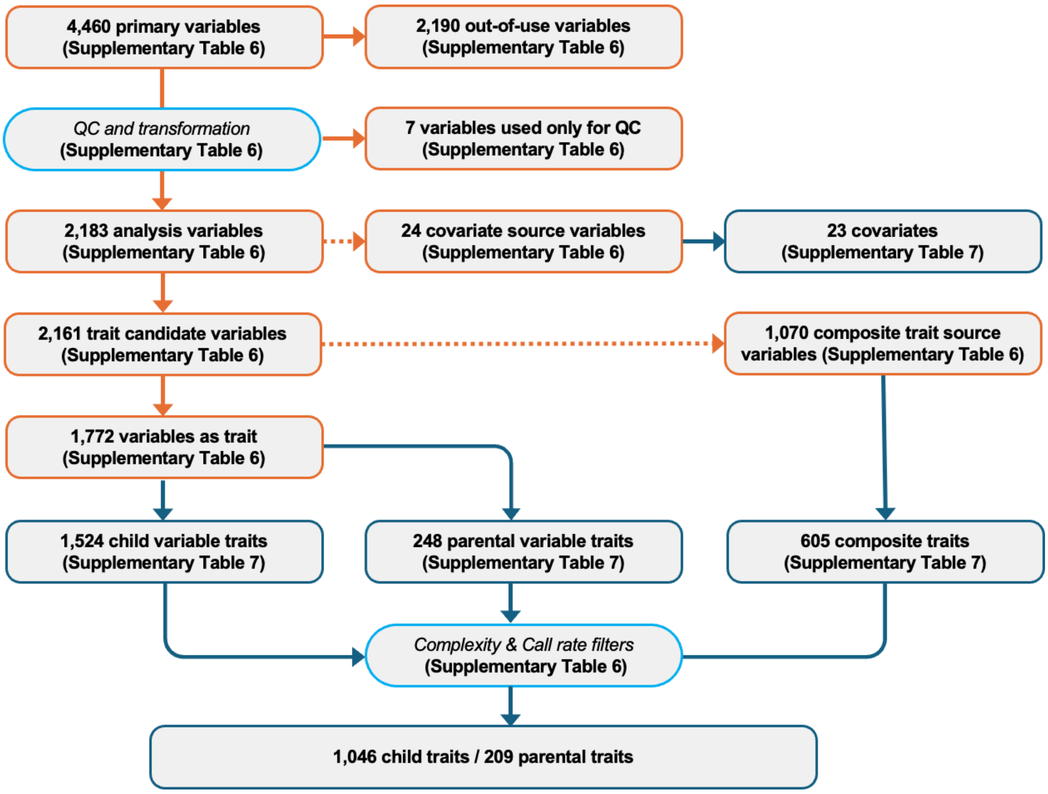

**Figure 1**: Variable selection workflow to create GWAS traits. Variables are indicated by orange boxes, and traits are indicated by navy boxes. Light blue boxes indicate pre- and post hoc QC procedures during trait creation. The dashed lines indicate the conditional branches are not mutually exclusive.

**Table 2**: Out-of-use flag information

| Flag | Category | Description |
|---|---|---|
| data_collection_flag | Metadata | Flags indicating whether data were collected |
| date | Metadata | Date variables |
| facility_flags | Metadata | Information on the external facilities where measurements or tests were performed |
| parental_metadata | Metadata | Parental metadata variables |
| pregnancy_information_flags | Metadata | Maternal pregnancy status at the time of questionnaire response |
| received_medical_checkup | Metadata | Indicator of whether a medical checkup was received |
| sex | Metadata | Sex information |
| unit_information | Metadata | Information on the unit centers where data were collected |
| agreement_and_withdrawn_flags | Nont-trait | Consent and withdrawal status flags |
| descriptive_answer | Nont-trait | Free-text response variables |
| duplicate | Nont-trait | Duplicate variables |
| inadequate_calcualtion_result | Nont-trait | Source variables used only to derive program-office–generated variables |
| job_code | Nont-trait | Occupation code variables |
| no_data_released | Nont-trait | Variables listed in the codebook but not released in the dataset |
| non_traits_age | Nont-trait | Age-related variables not treated as traits |
| phenotype_calculation_method | Nont-trait | Indicators of the calculation method used for the corresponding variable |
| uninformative_biospecimen | Nont-trait | Biospecimen collection information, such as collection time |
| uninformative_score_status | Nont-trait | Score-status variables not used in downstream analyses |
| ambiguous_target_definition | Out-of-scope | Variables with ambiguous target definitions (e.g., partner-related variables) |
| other_option | Out-of-scope | "Other" response options that are difficult to interpret without additional context |
| parental_outcome | Out-of-scope | Parental outcome variables outside the scope of the current study |
| unavailable | Restricted | Variables excluded from analysis due to data-use restrictions |

We obtained 2,161 potentially heritable variables, of which we classified the traits available for GWAS as primary variables or not, and found that 1,772 variables could be analyzed independently as traits (**Figure 1; Supplementary Table 6**,7). We confirmed that 248 traits were relevant to caregivers but not to children. However, we decided to analyze them, because they were useful for quantifying environmental exposures during pregnancy or cultural aspects of children after birth.

In parallel, we used 1,070 source variables for creating additional 605 composite traits, as some variables could not be analyzed independently (e.g., due to copyright) or be calculated a priori (e.g., BMI) (**Figure 1; Supplementary Table 6**). **Section 4** describes the details of how we constructed composite traits from the source variables. **Supplementary Table** 7 of the flagship paper also describes which variables were used to create the corresponding trait. Note that the variables are not mutually exclusive in the workflow (**Figure 1**). For example, the measurement of height at particular age is analyzed and also used as a composite source variable for computing BMI. Therefore, the total of *variables as trait* and *source variables for creating composite traits* are not equal to the number of *trait candidate variables*.

Once we had created a set of traits, we applied the final filters to determine whether to conduct a GWAS based on the trait call rate and complexity (**Figure 1**, **Supplementary Table** 7). This may reduce the potential false positive rate of variant associations. **Section 5** describes the common filters used to select traits. At last, we obtained 1,255 traits for the subsequent GWAS.

### 3. Quality controls and transformation

During the variable selection stage, appropriate data cleaning and transformations were applied to each variable, because the original data freeze was incomplete or conditioned by other variables. In addition, outliers with a high probability of misdescription were systematically excluded.

#### 3.1. Conditioning on other variable(s)

We introduced a standardized syntax for automated correction of a target variable using a conditional variable (e.g., the cadmium exposure variable `cd` and its left-censoring variable `cdc`), specified as:

*(ConditionalVariable):(value of conditional variable):(replacement value)*

Using e.g., `R`, the syntax is represented by:

```
target_var[cond_var%in%"value_of_cond_var"]="replacement value"
```

This syntax was also applied only for missing values (blanks) of the target variable:

```
target_var[target_var%in%""&cond_var%in%"value_of_cond_var"]="repl
acement value"
```

This is particularly useful when questionnaires are nested and the answer to a higher-level question determines the value of a lower-level question. For example, once you give a particular answer to a higher-level question, you don't need to answer the rest of the questionnaire (meaning the value to be "" (blank) for the lower-level question). All applied procedures are described in **Supplementary Table 6**.

#### 3.2 Outlier detection

For 282 numerical variables with continuous values, we excluded data points outside four standard deviations (SD), unless there were clear physiological or measurement limits (e.g. ASQ3 scores and certain laboratory measurements, including blood TSH levels). These criteria for valid data were discussed and agreed upon by a team comprising paediatricians and experts in epidemiological and statistical analysis. Additionally, for 66 variables with discrete values (e.g. count data), for which asymptotic normality does not apply, we did not introduce any upper bound because the distribution tends to be skewed and heavily weighted towards the right end. The specific cut-off values for each numerical variable are described in **Supplementary Table 6** of the flagship paper. For binary and multiple-choice categorical variables (including ordered categorical variables), all data with undefined values in the questionnaires were excluded.

#### 3.3 Replacement

Due to the questionnaire design, some binary variables were coded as "1" for controls and "0" for cases (of some disease, symptom, and so on). For consistency, we reversed these encodings so that "0" corresponds to controls and "1" to cases. Similarly, some ordered categorical variables were coded with levels in the reverse of their intuitive order (e.g., 1 = always, 2 = sometimes, 3 = never). In such cases, we reversed the order of the levels so that higher values consistently correspond to greater intensity or frequency of the trait.

We used the standardized syntax for automated replacement, specified as

*(Variable):(original level):(replacement level)*

which means that, in `R`, for example:

```
variable[variable%in%"original level"]="replacement level"
```

All procedures are also described in **Supplementary Table 6**.

In reality, multiple replacement procedures were applied simultaneously to the same variable (e.g. in the case-control data). In this case, the former replacement affects the latter. To avoid confusion, we internally create a temporary variable to which multiple replacements are applied. In `R`, for example, the following is applied for multiple *L* replacements:

```
tmp_var <- var
tmp_var[var%in%"original level 1"]="replacement level 1"
...
tmp_var[var%in%"original level L"]="replacement level L"
var <- tmp_var
```

#### 3.4 Offset of binary questionnaires

Due to the questionnaire definition, some binary variables are encoded as "1" for controls and "2" for cases. In this case, we subtract one (-1) from the variable to encode "0" for controls and "1" for cases.

##### 3.5 Masking responses other than those from the mother

Some questionnaires are completed by the mother, father, grandparents, or other caregivers. Therefore, the corresponding outcomes should be stratified according to the respondent. In JECS, more than 95% of questionnaires were completed by mothers, so all questionnaires targeting respondents were treated as mother-reported traits, and responses from non-mother respondents were set to NA.

### 4. Composite traits

This section provides a comprehensive description of composite traits constructed for the study. The purpose of this section is to ensure full transparency in trait construction and to facilitate reproducibility for future research.

##### 4.1 Categorical variables with multiple choices

The current GWAS pipeline, which is based on REGENIE, only allows us to analyse binary or quantitative traits. Therefore, in principle, a categorical variable with multiple choices is converted into a set of binary traits using one-hot encoding. Some categorical variables allow you to select multiple answers. In this case, the created binary traits are not mutually exclusive (see the next subsection on the vaccination questionnaire).

##### 4.2 Binarization of numerical variables

Some of the numerical variables are not suitable for analysis as quantitative traits because too many identical values or answers were observed for the variable. These repeated values skew the overall distribution, making it non-Gaussian. This violates the normality assumption in the linear regression model, causing a high false positive rate of variant association, especially for variants with a low minor allele frequency. For the numerical variables (including count variables or ordered categorical variables), we set the threshold at the mode of the empirical distribution. Let $p(x)$ denote the empirical probability mass function and let $m = \arg\max_x\ p(x)$ denote its mode. We then created binary variables using the indicator function $I(\cdot)$, which takes the value 1 if the condition is true and 0 otherwise. If $m$ was the minimum or maximum observed value, we created a single binary variable, $I(x > m)$ or $I(x < m)$, respectively. Otherwise, we created two binary variables, $I(x \leq m)$ or $I(x \geq m)$. These derived variables were then analysed as binary traits.

#### 4.3 Complex composite traits from multiple questionnaire variables

In this subsection, we introduce the following complex composite trait groups, typically involving the combination of multiple variables, in thematic order: 11 BMI traits, early pregnancy HbA1c (NGSP), 4 perinatal-related traits, 8 breastfeeding-related traits, 7 first vaccination-related traits, 25 food allergy-related traits, 31 feeding status-related traits, 18 rhinitis-related traits, 38 medical history-related traits, 1 eczema-related trait, 35 sleep-related traits, 3 childcare facility use-related traits, 1 toilet training-related trait, and 8 ASQ-3 questionnaire-related traits. For each trait group, we first provide a brief overview and then present a table listing the individual traits, their descriptions, source variables, and definitions in R-style pseudocode.

##### BMI traits

These traits represent body mass index (BMI) calculated at multiple ages from birth to 4 years. BMI traits enable consistent evaluation of physical growth trajectories and facilitate analyses relating early-life growth patterns to later health outcomes. Here we introduce 11 traits derived from weight and height measurements recorded at specific developmental time points.

| Trait_ID | Description | Source variables | Definition |
|---|---|---|---|
| dr0m_bmi | BMI at birth | Weight: Dr0m_0020801; Height: Dr0m_0020802 | (Dr0m_0020801 / 1000) / (Dr0m_0020802 / 100)^2 |
| dr1m_bmi | BMI at 1 month | Weight: Dr1m_0040401; Height: Dr1m_0040403 | (Dr1m_0040401 / 1000) / (Dr1m_0040403 / 100)^2 |
| c6m_bmi | BMI at 6 months | Weight: C6m_0070102_checked; Height: C6m_0070103_checked | (C6m_0070102_checked / 1000) / (C6m_0070103_checked / 100)^2 |
| c1y_1_bmi | BMI at 1 year (earlier measurement) | Weight: C1Y_0080103_checked; Height: C1Y_0080104_checked | (C1Y_0080103_checked / 1000) / (C1Y_0080104_checked / 100)^2 |
| c1y_2_bmi | BMI at 1 year (later measurement) | Weight: C1Y_0080203_checked; Height: C1Y_0080204_checked | (C1Y_0080203_checked / 1000) / (C1Y_0080204_checked / 100)^2 |
| c1hy_bmi | BMI at 1.5 years | Weight: C1hY_0060003_checked; Height: C1hY_0060002_checked | C1hY_0060003_checked / (C1hY_0060002_checked / 100)^2 |
| c2y_bmi | BMI at 2 years | Weight: C2Y_0030003_checked; Height: C2Y_0030002_checked | C2Y_0030003_checked / (C2Y_0030002_checked / 100)^2 |
| c2hy_bmi | BMI at 2.5 years | Weight: C2hY_0030003_checked; Height: C2hY_0030002_checked | C2hY_0030003_checked / (C2hY_0030002_checked / 100)^2 |
| c3y_bmi | BMI at 3 years | Weight: C3Y_0030003_checked; Height: C3Y_0030002_checked | C3Y_0030003_checked / (C3Y_0030002_checked / 100)^2 |
| c3hy_bmi | BMI at 3.5 years | Weight: C3hY_0030003; Height: C3hY_0030002 | C3hY_0030003 / (C3hY_0030002 / 100)^2 |
| c4y_bmi | BMI at 4 years | Weight: C4Y_0030003; Height: C4Y_0030002 | C4Y_0030003 / (C4Y_0030002 / 100)^2 |

##### Early pregnancy HbA1c (NGSP)

HbA1c measured in early pregnancy was harmonized to the National Glycohemoglobin Standardization Program (NGSP) scale to ensure consistency across records. Because the recorded HbA1c value could be reported either in the Japan Diabetes Society (JDS) scale or already in the NGSP scale, the measurement unit flag was used to determine whether conversion was required. This derived variable provides a unified early-pregnancy HbA1c measure for downstream epidemiological and genetic analyses.

| Trait_ID | Description | Source variables | Definition |
|---|---|---|---|
| hba1c_ngsp_early | Early pregnancy HbA1c (NGSP) | HbA1cflg, MT1b_0010001 | 1.02 * MT1b_0010001 + 0.25 if HbA1cflg == 1; MT1b_0010001 if HbA1cflg == 2 |

##### Perinatal-related traits

Perinatal-related traits capture key characteristics of pregnancy, delivery, and neonatal status. These factors are closely linked to early brain development, vulnerability to perinatal complications, and later neurodevelopmental outcomes. Here we introduce 4 traits derived from questionnaire variables related to perinatal events.

| Trait_ID | Description | Source variables | Definition |
|---|---|---|---|
| premature_birth | Premature birth | birth_w, birth_d | ifelse((birth_w * 7 + birth_d) <= 258, 1, 0) |
| postterm_birth | Postterm birth | birth_w, birth_d | ifelse((birth_w * 7 + birth_d) >= 294, 1, 0) |
| low_birth_weight | Low birth weight | Dr0m_0020801 | ifelse(Dr0m_0020801 < 2500, 1, 0) |
| length_of_labor | Length of labor | Dr0m_0020501, Dr0m_0020502 | Dr0m_0020501 * 60 + Dr0m_0020502 |

##### Breastfeeding-related traits

Breastfeeding-related traits capture breastfeeding discontinuation, breastfeeding duration, exclusive breastfeeding duration, and feeding frequency. These traits are useful for characterizing infant feeding patterns and their potential developmental or health impacts. Here we introduce 8 traits derived from questionnaire variables related to breastfeeding and formula feeding.

| Trait_ID | Description | Source variables | Definition |
|---|---|---|---|
| breast_feeding_cessation_1m | Discontinuation of breastfeeding by 1 month old | C6m_0010001–C6m_0010006, C1Y_0030007–C1Y_0030012 | ifelse(any(C6m_0010001 == 1) & sum(C6m_0010002:C6m_0010006, C1Y_0030007:C1Y_0030012) == 0, 1, 0) |
| breast_feeding_cessation_2m | Discontinuation of breastfeeding by 2 months old | C6m_0010001–C6m_0010006, C1Y_0030007–C1Y_0030012 | ifelse(any(C6m_0010001:C6m_0010002 == 1) & sum(C6m_0010003:C6m_0010006, C1Y_0030007:C1Y_0030012) == 0, 1, 0) |
| breast_feeding_cessation_3m | Discontinuation of breastfeeding by 3 months old | C6m_0010001–C6m_0010006, C1Y_0030007–C1Y_0030012 | ifelse(any(C6m_0010001:C6m_0010003 == 1) & sum(C6m_0010004:C6m_0010006, C1Y_0030007:C1Y_0030012) == 0, 1, 0) |
| breast_feeding_period_1y | Duration of breastfeeding up to 12 months old | C6m_0010001–C6m_0010006, C1Y_0030007–C1Y_0030012 | sum(all source variables) |
| exclusive_breastfeeding_1y | Duration of exclusive breastfeeding up to 12 months old | C6m_0010001–C6m_0010012, C1Y_0030007–C1Y_0030112 | sum(ifelse(C6m_0010001:C6m_0010006 == 1 & C6m_0010007:C6m_0010012 != 1, 1, 0), ifelse(C1Y_0030007:C1Y_0030012 == 1 & C1Y_0030107:C1Y_0030112 != 1, 1, 0)) |
| breast_feeding_cessation_age | Timing of breastfeeding cessation | C4Y_0490101, C4Y_0490102, C2Y_0080101, C2Y_0080102 | max(C4Y_0490101 * 12 + C4Y_0490102, C2Y_0080101 * 12 + C2Y_0080102), using the non-missing value when only one is available |
| feeding_type_yesterday_breast_feeding_count_1m | Number of breastfeeding episodes yesterday at 1 month old | M1m_0110101, M1m_0110201 | M1m_0110101 + M1m_0110201 |
| feeding_type_yesterday_formula_feeding_count_1m | Number of formula feeding episodes yesterday at 1 month old | M1m_0110203, M1m_0110301 | M1m_0110203 + M1m_0110301 |

##### First vaccination-related traits

These traits indicate whether specific vaccines were included in the child's initial vaccination series. Such traits allow assessment of early immunization patterns, adherence to recommended schedules, and potential associations between early vaccine exposure and later health outcomes. Here we introduce 7 traits derived from questionnaire items on first vaccination. In the questionnaire, respondents could select multiple responses from five tick boxes corresponding to eight vaccine types.

| Trait_ID | Description | Source variables | Definition |
|---|---|---|---|
| first_vaccination_dpt | DPT vaccine included in initial vaccination | C6m_0100101–C6m_0100105 | ifelse(any(source variables == 1), 1, 0) |
| first_vaccination_bcg | BCG vaccine included in initial vaccination | C6m_0100101–C6m_0100105 | ifelse(any(source variables == 2), 1, 0) |
| first_vaccination_polio | Polio vaccine included in initial vaccination | C6m_0100101–C6m_0100105 | ifelse(any(source variables == 3), 1, 0) |
| first_vaccination_rotavirus | Rotavirus vaccine included in initial vaccination | C6m_0100101–C6m_0100105 | ifelse(any(source variables == 5), 1, 0) |
| first_vaccination_haemophilus_b_influenzae | Haemophilus influenzae type b vaccine included in initial vaccination | C6m_0100101–C6m_0100105 | ifelse(any(source variables == 6), 1, 0) |
| first_vaccination_pneumococcus | Pneumococcus vaccine included in initial vaccination | C6m_0100101–C6m_0100105 | ifelse(any(source variables == 7), 1, 0) |
| first_vaccination_hbv | Hepatitis B virus (HBV) vaccine included in initial vaccination | C6m_0100101–C6m_0100105 | ifelse(any(source variables == 8), 1, 0) |

#### Food allergy-related traits

These traits summarize age-specific, cumulative, and food-specific patterns of food allergy or sensitization from 1.5 to 4 years of age. Categorizing these variables enables structured evaluation of allergy onset, persistence, and breadth across food groups, supporting analyses of early immune development and environmental influences. Here we introduce 25 additional traits composed of the questionnaires about food allergy.

| Trait_ID | Description | Source variables | Definition |
|---|---|---|---|
| egg_allergy_2y | Ongoing egg allergy at 2 years old | C2Y_0100102, C2Y_0100301–C2Y_0100305 | ifelse(C2Y_0100102 == 1 & any(C2Y_0100301:C2Y_0100305 == 1), 1, 0) |
| egg_allergy_3y | Ongoing egg allergy at 3 years old | C3Y_0110101, C3Y_0110301–C3Y_0110305 | ifelse(C3Y_0110101 == 1 & any(C3Y_0110301:C3Y_0110305 == 1), 1, 0) |
| egg_allergy_4y | Ongoing egg allergy at 4 years old | C4Y_0070101, C4Y_0070301–C4Y_0070305 | ifelse(C4Y_0070101 == 1 & any(C4Y_0070301:C4Y_0070305 == 1), 1, 0) |
| egg_allergy_1hy | Ongoing egg allergy at 1.5 years old | C1hY_0100102, C1hY_0100301–C1hY_0100305 | ifelse(C1hY_0100102 == 1 & any(C1hY_0100301:C1hY_0100305 == 1), 1, 0) |
| milk_allergy_1hy | Ongoing milk allergy at 1.5 years old | C1hY_0100602, C1hY_0100801–C1hY_0100805 | ifelse(C1hY_0100602 == 1 & any(C1hY_0100801:C1hY_0100805 == 1), 1, 0) |
| milk_allergy_2y | Ongoing milk allergy at 2 years old | C2Y_0100602, C2Y_0100801–C2Y_0100805 | ifelse(C2Y_0100602 == 1 & any(C2Y_0100801:C2Y_0100805 == 1), 1, 0) |
| milk_allergy_3y | Ongoing milk allergy at 3 years old | C3Y_0110601, C3Y_0110801–C3Y_0110805 | ifelse(C3Y_0110601 == 1 & any(C3Y_0110801:C3Y_0110805 == 1), 1, 0) |
| egg_allergy_cum_4y | History of egg allergy up to 4 years old | C1hY_0100102, C1hY_0100301–C1hY_0100305, C2Y_0100102, C2Y_0100301–C2Y_0100305, C3Y_0110101, C3Y_0110301–C3Y_0110305, C4Y_0070101, C4Y_0070301–C4Y_0070305 | ifelse((C1hY_0100102 == 1 & any(C1hY_0100301:C1hY_0100305 == 1)) \| (C2Y_0100102 == 1 & any(C2Y_0100301:C2Y_0100305 == 1)) \| (C3Y_0110101 == 1 & any(C3Y_0110301:C3Y_0110305 == 1)) \| (C4Y_0070101 == 1 & any(C4Y_0070301:C4Y_0070305 == 1)), 1, 0) |
| milk_allergy_cum_4y | History of milk allergy up to 4 years old | C1hY_0100602, C1hY_0100801–C1hY_0100805, C2Y_0100602, C2Y_0100801–C2Y_0100805, C3Y_0110601, C3Y_0110801–C3Y_0110805, C4Y_0070601, C4Y_0070801–C4Y_0070805 | ifelse((C1hY_0100602 == 1 & any(C1hY_0100801:C1hY_0100805 == 1)) \| (C2Y_0100602 == 1 & any(C2Y_0100801:C2Y_0100805 == 1)) \| (C3Y_0110601 == 1 & any(C3Y_0110801:C3Y_0110805 == |

| | | | |
|---|---|---|---|
| | | | 1)) \| (C4Y_0070601 == 1 & any(C4Y_0070801:C4Y_0070805 == 1)), 1, 0) |
| egg_allergy_score_until_4yo | Cumulative egg allergy score up to 4 years old | C4Y_0070101, C4Y_0070201, C4Y_0070301–C4Y_0070305, C3Y_0110101, C3Y_0110201, C3Y_0110301–C3Y_0110305, C2Y_0100102, C2Y_0100203, C2Y_0100301–C2Y_0100305, C1hY_0100102, C1hY_0100203, C1hY_0100301–C1hY_0100305 | sum(all source variables) |
| milk_allergy_score_until_4yo | Cumulative milk allergy score up to 4 years old | C4Y_0070601, C4Y_0070701, C4Y_0070801–C4Y_0070805, C3Y_0110601, C3Y_0110701, C3Y_0110801–C3Y_0110805, C2Y_0100602, C2Y_0100703, C2Y_0100801–C2Y_0100805, C1hY_0100602, C1hY_0100703, C1hY_0100801–C1hY_0100805 | sum(all source variables) |
| egg_allergy_score_until_4yo_positive | Egg allergy score positive up to 4 years old | C4Y_0070101, C4Y_0070201, C4Y_0070301–C4Y_0070305, C3Y_0110101, C3Y_0110201, C3Y_0110301–C3Y_0110305, C2Y_0100102, C2Y_0100203, C2Y_0100301–C2Y_0100305, C1hY_0100102, C1hY_0100203, C1hY_0100301–C1hY_0100305 | ifelse(any(source variables == 1), 1, 0) |
| milk_allergy_score_until_4yo_positive | Milk allergy score positive up to 4 years old | C4Y_0070601, C4Y_0070701, C4Y_0070801–C4Y_0070805, C3Y_0110601, C3Y_0110701, C3Y_0110801–C3Y_0110805, C2Y_0100602, C2Y_0100703, C2Y_0100801–C2Y_0100805, C1hY_0100602, C1hY_0100703, C1hY_0100801–C1hY_0100805 | ifelse(any(source variables == 1), 1, 0) |
| wheat_allergy_score_until_4yo_positive | Wheat allergy score positive up to 4 years old | C4Y_0071101, C4Y_0071201, C4Y_0071301–C4Y_0071305, C3Y_0111101, C3Y_0111201, C3Y_0111301–C3Y_0111305, C2Y_0101102, C2Y_0101203, C2Y_0101301–C2Y_0101305, C1hY_0101102, C1hY_0101203, C1hY_0101301–C1hY_0101305 | ifelse(any(source variables == 1), 1, 0) |
| soybean_allergy_score_until_4yo_positive | Soybean allergy score positive up to 4 years old | C4Y_0071601, C4Y_0071701, C4Y_0071801–C4Y_0071805, C3Y_0111601, C3Y_0111701, C3Y_0111801–C3Y_0111805, C2Y_0101602, C2Y_0101703, C2Y_0101801–C2Y_0101805, C1hY_0101602, C1hY_0101703, C1hY_0101801–C1hY_0101805 | ifelse(any(source variables == 1), 1, 0) |
| fish_allergy_score_until_4yo_positive | Fish allergy score positive up to 4 years old | C4Y_0072101, C4Y_0072201, C4Y_0072301–C4Y_0072305, C3Y_0112101, C3Y_0112201, C3Y_0112301–C3Y_0112305, C2Y_0102102, C2Y_0102203, C2Y_0102301–C2Y_0102305, C1hY_0102102, C1hY_0102203, C1hY_0102301–C1hY_0102305 | ifelse(any(source variables == 1), 1, 0) |
| fruit_allergy_score_until_4yo_positive | Fruit allergy score positive up to 4 years old | C4Y_0073101, C4Y_0073201, C4Y_0073301–C4Y_0073305, C3Y_0113101, C3Y_0113201, C3Y_0113301–C3Y_0113305, C2Y_0103102, C2Y_0103203, C2Y_0103301–C2Y_0103305, C1hY_0103102, C1hY_0103203, C1hY_0103301–C1hY_0103305 | ifelse(any(source variables == 1), 1, 0) |
| shellfish_allergy_score_until_4yo_positive | Shellfish allergy score positive up to 4 years old | C4Y_0073601, C4Y_0073701, C4Y_0073801–C4Y_0073805, C3Y_0113601, C3Y_0113701, C3Y_0113801–C3Y_0113805, C2Y_0103602, C2Y_0103703, C2Y_0103801–C2Y_0103805, C1hY_0103602, C1hY_0103703, C1hY_0103801–C1hY_0103805 | ifelse(any(source variables == 1), 1, 0) |
| soba_allergy_score_until_4yo_positive | Soba allergy score positive up to 4 years old | C4Y_0074101, C4Y_0074201, C4Y_0074301–C4Y_0074305, C3Y_0114101, C3Y_0114201, C3Y_0114301–C3Y_0114305, C2Y_0104102, C2Y_0104203, C2Y_0104301–C2Y_0104305, C1hY_0104102, C1hY_0104203, C1hY_0104301–C1hY_0104305 | ifelse(any(source variables == 1), 1, 0) |
| peanuts_allergy_score_4yo_positive | Peanut allergy score positive at 4 years old | C4Y_0075101, C4Y_0075201, C4Y_0075301–C4Y_0075305 | ifelse(any(source variables == 1), 1, 0) |
| nuts_allergy_score_until_4yo_positive | Nut allergy score positive up to 4 years old | C3Y_0115101, C3Y_0115201, C3Y_0115301–C3Y_0115305, C2Y_0105102, C2Y_0105203, C2Y_0105301–C2Y_0105305, C1hY_0105102, C1hY_0105203, C1hY_0105301–C1hY_0105305, C4Y_0075101, C4Y_0075601, C4Y_0075201, C4Y_0075701, C4Y_0075301, C4Y_0075801, C4Y_0075302, C4Y_0075802, C4Y_0075303, C4Y_0075803, C4Y_0075304, C4Y_0075804, C4Y_0075305, C4Y_0075805 | ifelse(any(source variables == 1), 1, 0) |
| participant_reported_food_allergy_cum_4y | Participant-reported food allergy up to 4 years old | C1hY_0100102, C1hY_0100301–C1hY_0100305, C2Y_0100102, C2Y_0100301–C2Y_0100305, C3Y_0110101, C3Y_0110301–C3Y_0110305, | ifelse(any(all source variables == 1), 1, 0) |

| | | | |
|---|---|---|---|
| | | C4Y_0070101, C4Y_0070301–C4Y_0070305, C1hY_0100602, C1hY_0100801–C1hY_0100805, C2Y_0100602, C2Y_0100801–C2Y_0100805, C3Y_0110601, C3Y_0110801–C3Y_0110805, C4Y_0070601, C4Y_0070801–C4Y_0070805, C1hY_0101102, C1hY_0101301–C1hY_0101305, C2Y_0101102, C2Y_0101301–C2Y_0101305, C3Y_0111101, C3Y_0111301–C3Y_0111305, C4Y_0071101, C4Y_0071301–C4Y_0071305, C1hY_0101602, C1hY_0101801–C1hY_0101805, C2Y_0101602, C2Y_0101801–C2Y_0101805, C3Y_0111601, C3Y_0111801–C3Y_0111805, C4Y_0071601, C4Y_0071801–C4Y_0071805, C1hY_0102102, C1hY_0102301–C1hY_0102305, C2Y_0102102, C2Y_0102301–C2Y_0102305, C3Y_0112101, C3Y_0112301–C3Y_0112305, C4Y_0072101, C4Y_0072301–C4Y_0072305, C1hY_0102602, C1hY_0102801–C1hY_0102805, C2Y_0102602, C2Y_0102801–C2Y_0102805, C3Y_0112601, C3Y_0112801–C3Y_0112805, C4Y_0072601, C4Y_0072801–C4Y_0072805, C1hY_0103102, C1hY_0103301–C1hY_0103305, C2Y_0103102, C2Y_0103301–C2Y_0103305, C3Y_0113101, C3Y_0113301–C3Y_0113305, C4Y_0073101, C4Y_0073301–C4Y_0073305, C1hY_0103602, C1hY_0103801–C1hY_0103805, C2Y_0103602, C2Y_0103801–C2Y_0103805, C3Y_0113601, C3Y_0113801–C3Y_0113805, C4Y_0073601, C4Y_0073801–C4Y_0073805, C1hY_0104102, C1hY_0104301–C1hY_0104305, C2Y_0104102, C2Y_0104301–C2Y_0104305, C3Y_0114101, C3Y_0114301–C3Y_0114305, C4Y_0074101, C4Y_0074301–C4Y_0074305, C1hY_0104602, C1hY_0104801–C1hY_0104805, C2Y_0104602, C2Y_0104801–C2Y_0104805, C3Y_0114601, C3Y_0114801–C3Y_0114805, C4Y_0074601, C4Y_0074801–C4Y_0074805, C1hY_0105102, C1hY_0105301–C1hY_0105305, C2Y_0105102, C2Y_0105301–C2Y_0105305, C3Y_0115101, C3Y_0115301–C3Y_0115305, C4Y_0075101, C4Y_0075301–C4Y_0075305, C4Y_0075601, C4Y_0075801–C4Y_0075805 | |
| food_allergy_score_cum_count_until_4yo | Cumulative number of food-specific allergic symptom types and sensitization | C4Y_0070101, C4Y_0070201, C4Y_0070301–C4Y_0070305, C3Y_0110101, C3Y_0110201, C3Y_0110301–C3Y_0110305, C2Y_0100102, C2Y_0100203, C2Y_0100301–C2Y_0100305, C1hY_0100102, C1hY_0100203, C1hY_0100301–C1hY_0100305, C4Y_0070601, C4Y_0070701, C4Y_0070801–C4Y_0070805, C3Y_0110601, C3Y_0110701, C3Y_0110801–C3Y_0110805, C2Y_0100602, C2Y_0100703, C2Y_0100801–C2Y_0100805, C1hY_0100602, C1hY_0100703, C1hY_0100801–C1hY_0100805, C4Y_0071101, C4Y_0071201, C4Y_0071301–C4Y_0071305, C3Y_0111101, C3Y_0111201, C3Y_0111301–C3Y_0111305, C2Y_0101102, C2Y_0101203, C2Y_0101301–C2Y_0101305, C1hY_0101102, C1hY_0101203, C1hY_0101301–C1hY_0101305, C4Y_0071601, C4Y_0071701, C4Y_0071801–C4Y_0071805, C3Y_0111601, C3Y_0111701, C3Y_0111801–C3Y_0111805, C2Y_0101602, C2Y_0101703, C2Y_0101801–C2Y_0101805, C1hY_0101602, C1hY_0101703, C1hY_0101801–C1hY_0101805, C4Y_0072101, C4Y_0072201, C4Y_0072301–C4Y_0072305, C3Y_0112101, C3Y_0112201, C3Y_0112301–C3Y_0112305, C2Y_0102102, C2Y_0102203, C2Y_0102301–C2Y_0102305, C1hY_0102102, C1hY_0102203, C1hY_0102301–C1hY_0102305, C4Y_0072601, C4Y_0072701, C4Y_0072801–C4Y_0072805, C3Y_0112601, C3Y_0112701, C3Y_0112801–C3Y_0112805, C2Y_0102602, C2Y_0102703, C2Y_0102801–C2Y_0102805, C1hY_0102602, C1hY_0102703, C1hY_0102801–C1hY_0102805, C4Y_0073101, C4Y_0073201, C4Y_0073301–C4Y_0073305, C3Y_0113101, C3Y_0113201, C3Y_0113301–C3Y_0113305, C2Y_0103102, C2Y_0103203, C2Y_0103301–C2Y_0103305, C1hY_0103102, C1hY_0103203, C1hY_0103301–C1hY_0103305, C4Y_0073601, C4Y_0073701, C4Y_0073801–C4Y_0073805, C3Y_0113601, | sum(all source variables) |

| | | | |
|---|---|---|---|
| | | C3Y_0113701, C3Y_0113801–C3Y_0113805, C2Y_0103602, C2Y_0103703, C2Y_0103801–C2Y_0103805, C1hY_0103602, C1hY_0103703, C1hY_0103801–C1hY_0103805, C4Y_0074101, C4Y_0074201, C4Y_0074301–C4Y_0074305, C3Y_0114101, C3Y_0114201, C3Y_0114301–C3Y_0114305, C2Y_0104102, C2Y_0104203, C2Y_0104301–C2Y_0104305, C1hY_0104102, C1hY_0104203, C1hY_0104301–C1hY_0104305, C4Y_0074601, C4Y_0074701, C4Y_0074801–C4Y_0074805, C3Y_0114601, C3Y_0114701, C3Y_0114801–C3Y_0114805, C2Y_0104602, C2Y_0104703, C2Y_0104801–C2Y_0104805, C1hY_0104602, C1hY_0104703, C1hY_0104801–C1hY_0104805, C3Y_0115101, C3Y_0115201, C3Y_0115301–C3Y_0115305, C2Y_0105102, C2Y_0105203, C2Y_0105301–C2Y_0105305, C1hY_0105102, C1hY_0105203, C1hY_0105301–C1hY_0105305, C4Y_0075101, C4Y_0075601, C4Y_0075201, C4Y_0075701, C4Y_0075301, C4Y_0075801, C4Y_0075302, C4Y_0075802, C4Y_0075303, C4Y_0075803, C4Y_0075304, C4Y_0075804, C4Y_0075305, C4Y_0075805 | |
| food_allergy_cum_count_until_4yo | Total number of causative food allergens | C1hY_0100102, C1hY_0100301–C1hY_0100305, C2Y_0100102, C2Y_0100301–C2Y_0100305, C3Y_0110101, C3Y_0110301–C3Y_0110305, C4Y_0070101, C4Y_0070301–C4Y_0070305, C1hY_0100602, C1hY_0100801–C1hY_0100805, C2Y_0100602, C2Y_0100801–C2Y_0100805, C3Y_0110601, C3Y_0110801–C3Y_0110805, C4Y_0070601, C4Y_0070801–C4Y_0070805, C1hY_0101102, C1hY_0101301–C1hY_0101305, C2Y_0101102, C2Y_0101301–C2Y_0101305, C3Y_0111101, C3Y_0111301–C3Y_0111305, C4Y_0071101, C4Y_0071301–C4Y_0071305, C1hY_0101602, C1hY_0101801–C1hY_0101805, C2Y_0101602, C2Y_0101801–C2Y_0101805, C3Y_0111601, C3Y_0111801–C3Y_0111805, C4Y_0071601, C4Y_0071801–C4Y_0071805, C1hY_0102102, C1hY_0102301–C1hY_0102305, C2Y_0102102, C2Y_0102301–C2Y_0102305, C3Y_0112101, C3Y_0112301–C3Y_0112305, C4Y_0072101, C4Y_0072301–C4Y_0072305, C1hY_0102602, C1hY_0102801–C1hY_0102805, C2Y_0102602, C2Y_0102801–C2Y_0102805, C3Y_0112601, C3Y_0112801–C3Y_0112805, C4Y_0072601, C4Y_0072801–C4Y_0072805, C1hY_0103102, C1hY_0103301–C1hY_0103305, C2Y_0103102, C2Y_0103301–C2Y_0103305, C3Y_0113101, C3Y_0113301–C3Y_0113305, C4Y_0073101, C4Y_0073301–C4Y_0073305, C1hY_0103602, C1hY_0103801–C1hY_0103805, C2Y_0103602, C2Y_0103801–C2Y_0103805, C3Y_0113601, C3Y_0113801–C3Y_0113805, C4Y_0073601, C4Y_0073801–C4Y_0073805, C1hY_0104102, C1hY_0104301–C1hY_0104305, C2Y_0104102, C2Y_0104301–C2Y_0104305, C3Y_0114101, C3Y_0114301–C3Y_0114305, C4Y_0074101, C4Y_0074301–C4Y_0074305, C1hY_0104602, C1hY_0104801–C1hY_0104805, C2Y_0104602, C2Y_0104801–C2Y_0104805, C3Y_0114601, C3Y_0114801–C3Y_0114805, C4Y_0074601, C4Y_0074801–C4Y_0074805, C1hY_0105102, C1hY_0105301–C1hY_0105305, C2Y_0105102, C2Y_0105301–C2Y_0105305, C3Y_0115101, C3Y_0115301–C3Y_0115305, C4Y_0075101, C4Y_0075301–C4Y_0075305, C4Y_0075601, C4Y_0075801–C4Y_0075805 | sum(all source variables) |
| any_food_allergy_score_positive_until_4yo | History of any food allergy or sensitization up to 4 years old | C4Y_0070101, C4Y_0070201, C4Y_0070301–C4Y_0070305, C3Y_0110101, C3Y_0110201, C3Y_0110301–C3Y_0110305, C2Y_0100102, C2Y_0100203, C2Y_0100301–C2Y_0100305, C1hY_0100102, C1hY_0100203, C1hY_0100301–C1hY_0100305, C4Y_0070601, C4Y_0070701, C4Y_0070801–C4Y_0070805, C3Y_0110601, C3Y_0110701, C3Y_0110801–C3Y_0110805, C2Y_0100602, C2Y_0100703, C2Y_0100801–C2Y_0100805, C1hY_0100602, C1hY_0100703, C1hY_0100801–C1hY_0100805, C4Y_0071101, C4Y_0071201, C4Y_0071301–C4Y_0071305, C3Y_0111101, | ifelse(any(source variables == 1), 1, 0) |

| | | |
|---|---|---|
| | | C3Y_0111201, C3Y_0111301–C3Y_0111305, C2Y_0101102, C2Y_0101203, C2Y_0101301–C2Y_0101305, C1hY_0101102, C1hY_0101203, C1hY_0101301–C1hY_0101305, C4Y_0071601, C4Y_0071701, C4Y_0071801–C4Y_0071805, C3Y_0111601, C3Y_0111701, C3Y_0111801–C3Y_0111805, C2Y_0101602, C2Y_0101703, C2Y_0101801–C2Y_0101805, C1hY_0101602, C1hY_0101703, C1hY_0101801–C1hY_0101805, C4Y_0072101, C4Y_0072201, C4Y_0072301–C4Y_0072305, C3Y_0112101, C3Y_0112201, C3Y_0112301–C3Y_0112305, C2Y_0102102, C2Y_0102203, C2Y_0102301–C2Y_0102305, C1hY_0102102, C1hY_0102203, C1hY_0102301–C1hY_0102305, C4Y_0072601, C4Y_0072701, C4Y_0072801–C4Y_0072805, C3Y_0112601, C3Y_0112701, C3Y_0112801–C3Y_0112805, C2Y_0102602, C2Y_0102703, C2Y_0102801–C2Y_0102805, C1hY_0102602, C1hY_0102703, C1hY_0102801–C1hY_0102805, C4Y_0073101, C4Y_0073201, C4Y_0073301–C4Y_0073305, C3Y_0113101, C3Y_0113201, C3Y_0113301–C3Y_0113305, C2Y_0103102, C2Y_0103203, C2Y_0103301–C2Y_0103305, C1hY_0103102, C1hY_0103203, C1hY_0103301–C1hY_0103305, C4Y_0073601, C4Y_0073701, C4Y_0073801–C4Y_0073805, C3Y_0113601, C3Y_0113701, C3Y_0113801–C3Y_0113805, C2Y_0103602, C2Y_0103703, C2Y_0103801–C2Y_0103805, C1hY_0103602, C1hY_0103703, C1hY_0103801–C1hY_0103805, C4Y_0074101, C4Y_0074201, C4Y_0074301–C4Y_0074305, C3Y_0114101, C3Y_0114201, C3Y_0114301–C3Y_0114305, C2Y_0104102, C2Y_0104203, C2Y_0104301–C2Y_0104305, C1hY_0104102, C1hY_0104203, C1hY_0104301–C1hY_0104305, C4Y_0074601, C4Y_0074701, C4Y_0074801–C4Y_0074805, C3Y_0114601, C3Y_0114701, C3Y_0114801–C3Y_0114805, C2Y_0104602, C2Y_0104703, C2Y_0104801–C2Y_0104805, C1hY_0104602, C1hY_0104703, C1hY_0104801–C1hY_0104805, C3Y_0115101, C3Y_0115201, C3Y_0115301–C3Y_0115305, C2Y_0105102, C2Y_0105203, C2Y_0105301–C2Y_0105305, C1hY_0105102, C1hY_0105203, C1hY_0105301–C1hY_0105305, C4Y_0075101, C4Y_0075601, C4Y_0075201, C4Y_0075701, C4Y_0075301, C4Y_0075801, C4Y_0075302, C4Y_0075802, C4Y_0075303, C4Y_0075803, C4Y_0075304, C4Y_0075804, C4Y_0075305, C4Y_0075805 |

#### Feeding status-related traits

Feeding status-related traits were designed to capture current non-consumption of specific foods in the context of possible allergic sensitization or food avoidance. In these questionnaire items, responses other than normal consumption may reflect clinically or epidemiologically meaningful patterns, such as foods that have never been introduced, are partially avoided, or are now completely avoided. These traits therefore provide a harmonized indicator of food non-consumption status across food items and ages.

| Trait_ID | Description | Source variables | Definition |
|---|---|---|---|
| food_allergy_egg_feeding_status_case_combined_1yh | Combined egg feeding status category at 1.5 years old | C1hY_0100001 | ifelse(source variable != 1, 1, 0) |
| food_allergy_milk_feeding_status_case_combined_1yh | Combined milk feeding status category at 1.5 years old | C1hY_0100501 | ifelse(source variable != 1, 1, 0) |
| food_allergy_wheat_feeding_status_case_combined_1yh | Combined wheat feeding status category at 1.5 years old | C1hY_0101001 | ifelse(source variable != 1, 1, 0) |
| food_allergy_fish_feeding_status_case_combined_1yh | Combined fish feeding status category at 1.5 years old | C1hY_0102001 | ifelse(source variable != 1, 1, 0) |
| food_allergy_fruit_feeding_status_case_combined_1yh | Combined fruit feeding status category at 1.5 years old | C1hY_0103001 | ifelse(source variable != 1, 1, 0) |

| | | | |
|---|---|---|---|
| food_allergy_shellfish_feeding_status_case_combined_1yh | Combined shellfish feeding status category at 1.5 years old | C1hY_0104001 | ifelse(source variable != 1, 1, 0) |
| food_allergy_soba_feeding_status_case_combined_1yh | Combined soba feeding status category at 1.5 years old | C1hY_0105001 | ifelse(source variable != 1, 1, 0) |
| food_allergy_sesame_feeding_status_case_combined_1yh | Combined sesame feeding status category at 1.5 years old | C1hY_0106001 | ifelse(source variable != 1, 1, 0) |
| food_allergy_nuts_feeding_status_case_combined_1yh | Combined nut feeding status category at 1.5 years old | C1hY_0107001 | ifelse(source variable != 1, 1, 0) |
| food_allergy_egg_feeding_status_case_combined_2y | Combined egg feeding status category at 2 years old | C2Y_0070501 | ifelse(source variable != 1, 1, 0) |
| food_allergy_milk_feeding_status_case_combined_2y | Combined milk feeding status category at 2 years old | C2Y_0071001 | ifelse(source variable != 1, 1, 0) |
| food_allergy_fish_feeding_status_case_combined_2y | Combined fish feeding status category at 2 years old | C2Y_0072001 | ifelse(source variable != 1, 1, 0) |
| food_allergy_fruit_feeding_status_case_combined_2y | Combined fruit feeding status category at 2 years old | C2Y_0073001 | ifelse(source variable != 1, 1, 0) |
| food_allergy_shellfish_feeding_status_case_combined_2y | Combined shellfish feeding status category at 2 years old | C2Y_0074001 | ifelse(source variable != 1, 1, 0) |
| food_allergy_soba_feeding_status_case_combined_2y | Combined soba feeding status category at 2 years old | C2Y_0075001 | ifelse(source variable != 1, 1, 0) |
| food_allergy_sesame_feeding_status_case_combined_2y | Combined sesame feeding status category at 2 years old | C2Y_0076001 | ifelse(source variable != 1, 1, 0) |
| food_allergy_nuts_feeding_status_case_combined_2y | Combined nut feeding status category at 2 years old | C2Y_0077001 | ifelse(source variable != 1, 1, 0) |
| food_allergy_egg_feeding_status_case_combined_3y | Combined egg feeding status category at 3 years old | C3Y_0100001 | ifelse(source variable != 1, 1, 0) |
| food_allergy_milk_feeding_status_case_combined_3y | Combined milk feeding status category at 3 years old | C3Y_0101001 | ifelse(source variable != 1, 1, 0) |
| food_allergy_fish_feeding_status_case_combined_3y | Combined fish feeding status category at 3 years old | C3Y_0103001 | ifelse(source variable != 1, 1, 0) |
| food_allergy_fruit_feeding_status_case_combined_3y | Combined fruit feeding status category at 3 years old | C3Y_0104001 | ifelse(source variable != 1, 1, 0) |
| food_allergy_shellfish_feeding_status_case_combined_3y | Combined shellfish feeding status category at 3 years old | C3Y_0105001 | ifelse(source variable != 1, 1, 0) |
| food_allergy_soba_feeding_status_case_combined_3y | Combined soba feeding status category at 3 years old | C3Y_0106001 | ifelse(source variable != 1, 1, 0) |
| food_allergy_sesame_feeding_status_case_combined_3y | Combined sesame feeding status category at 3 years old | C3Y_0107001 | ifelse(source variable != 1, 1, 0) |
| food_allergy_nuts_feeding_status_case_combined_3y | Combined nut feeding status category at 3 years old | C3Y_0108001 | ifelse(source variable != 1, 1, 0) |
| food_allergy_egg_feeding_status_case_combined_4y | Combined egg feeding status category at 4 years old | C4Y_0110001 | ifelse(source variable != 1, 1, 0) |
| food_allergy_milk_feeding_status_case_combined_4y | Combined milk feeding status category at 4 years old | C4Y_0111001 | ifelse(source variable != 1, 1, 0) |
| food_allergy_shellfish_feeding_status_case_combined_4y | Combined shellfish feeding status category at 4 years old | C4Y_0115001 | ifelse(source variable != 1, 1, 0) |
| food_allergy_soba_feeding_status_case_combined_4y | Combined soba feeding status category at 4 years old | C4Y_0116001 | ifelse(source variable != 1, 1, 0) |
| food_allergy_peanuts_feeding_status_case_combined_4y | Combined peanut feeding status category at 4 years old | C4Y_0118001 | ifelse(source variable != 1, 1, 0) |
| food_allergy_non_peanuts_nuts_feeding_status_case_combined_4y | Combined non-peanut nut feeding status category at 4 years old | C4Y_0119001 | ifelse(source variable != 1, 1, 0) |

#### Rhinitis-related traits

Rhinitis-related traits were constructed from modified ISAAC questionnaire items capturing the seasonal occurrence of runny nose symptoms. Because each individual item reflects symptom occurrence only within a limited seasonal window, separate items can be sparse and may not fully capture broader seasonal patterns. Composite traits were therefore defined by combining related questionnaire variables within each season or season group.

| Trait_ID | Description | Source variables | Definition |
|---|---|---|---|
| isaac_modified_current_runny_nose_spring_2y | Runny nose in spring at 2 years old based on modified ISAAC | C2Y_0210203–C2Y_0210205 | ifelse(any(source variables == 1), 1, 0) |
| isaac_modified_current_runny_nose_summer_2y | Runny nose in summer at 2 years old based on modified ISAAC | C2Y_0210206–C2Y_0210208 | ifelse(any(source variables == 1), 1, 0) |
| isaac_modified_current_runny_nose_autumn_2y | Runny nose in autumn at 2 years old based on modified ISAAC | C2Y_0210209–C2Y_0210211 | ifelse(any(source variables == 1), 1, 0) |
| isaac_modified_current_runny_nose_winter_2y | Runny nose in winter at 2 years old based on modified ISAAC | C2Y_0210201, C2Y_0210202, C2Y_0210212 | ifelse(any(source variables == 1), 1, 0) |
| isaac_modified_current_runny_nose_pollen_season_2y | Runny nose during pollen season at 2 years old based on modified ISAAC | C2Y_0210202–C2Y_0210205 | ifelse(any(source variables == 1), 1, 0) |
| isaac_modified_current_runny_nose_nonpollen_season_2y | Runny nose during non-pollen season at 2 years old based on modified ISAAC | C2Y_0210201, C2Y_0210206–C2Y_0210212 | ifelse(any(source variables == 1), 1, 0) |
| isaac_modified_current_runny_nose_spring_3y | Runny nose in spring at 3 years old based on modified ISAAC | C3Y_0220203–C3Y_0220205 | ifelse(any(source variables == 1), 1, 0) |
| isaac_modified_current_runny_nose_summer_3y | Runny nose in summer at 3 years old based on modified ISAAC | C3Y_0220206–C3Y_0220208 | ifelse(any(source variables == 1), 1, 0) |
| isaac_modified_current_runny_nose_autumn_3y | Runny nose in autumn at 3 years old based on modified ISAAC | C3Y_0220209–C3Y_0220211 | ifelse(any(source variables == 1), 1, 0) |
| isaac_modified_current_runny_nose_winter_3y | Runny nose in winter at 3 years old based on modified ISAAC | C3Y_0220201, C3Y_0220202, C3Y_0220212 | ifelse(any(source variables == 1), 1, 0) |
| isaac_modified_current_runny_nose_pollen_season_3y | Runny nose during pollen season at 3 years old based on modified ISAAC | C3Y_0220202–C3Y_0220205 | ifelse(any(source variables == 1), 1, 0) |
| isaac_modified_current_runny_nose_nonpollen_season_3y | Runny nose during non-pollen season at 3 years old based on modified ISAAC | C3Y_0220201, C3Y_0220206–C3Y_0220212 | ifelse(any(source variables == 1), 1, 0) |
| isaac_modified_current_runny_nose_spring_4y | Runny nose in spring at 4 years old based on modified ISAAC | C4Y_0250203–C4Y_0250205 | ifelse(any(source variables == 1), 1, 0) |
| isaac_modified_current_runny_nose_summer_4y | Runny nose in summer at 4 years old based on modified ISAAC | C4Y_0250206–C4Y_0250208 | ifelse(any(source variables == 1), 1, 0) |
| isaac_modified_current_runny_nose_autumn_4y | Runny nose in autumn at 4 years old based on modified ISAAC | C4Y_0250209–C4Y_0250211 | ifelse(any(source variables == 1), 1, 0) |
| isaac_modified_current_runny_nose_winter_4y | Runny nose in winter at 4 years old based on modified ISAAC | C4Y_0250201, C4Y_0250202, C4Y_0250212 | ifelse(any(source variables == 1), 1, 0) |
| isaac_modified_current_runny_nose_pollen_season_4y | Runny nose during pollen season at 4 years old based on modified ISAAC | C4Y_0250202–C4Y_0250205 | ifelse(any(source variables == 1), 1, 0) |
| isaac_modified_current_runny_nose_nonpollen_season_4y | Runny nose during non-pollen season at 4 years old based on modified ISAAC | C4Y_0250201, C4Y_0250206–C4Y_0250212 | ifelse(any(source variables == 1), 1, 0) |

##### Medical history-related traits

Medical history-related traits summarize clinically important health conditions and symptom burdens during early childhood. These traits enable integrative analyses of early-life disease burden and related symptom patterns. To improve readability, we grouped the 38 traits into four categories: (i) cumulative medical history traits indicating whether a condition had ever been reported during the observation period, (ii) cumulative fever count traits summarizing the total number of fever episodes reported up to a given age, (iii) constipation severity traits based on summed symptom items at a given age, and (iv) constipation status traits indicating ongoing or cumulative constipation based on predefined symptom thresholds. Here we introduce 36 traits derived from questionnaire variables related to medical history.

| Trait_ID | Description | Source variables | Definition |
|---|---|---|---|
| kawasaki_disease_cum_4y | Past medical history of Kawasaki disease | C6m_0112001, C1Y_0112401, C2Y_0052001, C3Y_0081001, C4Y_0041001 | ifelse(any(source variables == 1), 1, 0) |
| febrile_convulsion_cum_4y | Past medical history of febrile convulsion | C6m_0112101, C1Y_0118001, C2Y_0053101, C3Y_0082101, | ifelse(any(source variables == 1), 1, 0) |

| | | | |
|---|---|---|---|
| | | C4Y_0042101 | |
| asthma_cum_4y | Past medical history of asthma | C1Y_0112001, C1hY_0090001, C2Y_0060001, C3Y_0091001, C4Y_0051001 | ifelse(any(source variables == 1), 1, 0) |
| food_allergy_cum_4y | Past medical history of food allergy | C1Y_0112201, C1hY_0090201, C2Y_0060101, C3Y_0091101, C4Y_0051101 | ifelse(any(source variables == 1), 1, 0) |
| atopic_dermatitis_cum_4y | Past medical history of atopic dermatitis | C1Y_0112101, C1hY_0090101, C2Y_0060201, C3Y_0091201, C4Y_0051201 | ifelse(any(source variables == 1), 1, 0) |
| urticaria_cum_4y | Past medical history of urticaria | C3Y_0091401, C4Y_0051401 | ifelse(any(source variables == 1), 1, 0) |
| allergic_conjunctivitis_cum_4y | Past medical history of allergic conjunctivitis up to 4 years old | C2Y_0060301, C3Y_0091301, C4Y_0051301 | ifelse(any(source variables == 1), 1, 0) |
| allergic_rhinitis_cum_4y | Past medical history of allergic rhinitis up to 4 years old | C2Y_0060401, C3Y_0091501, C4Y_0051501 | ifelse(any(source variables == 1), 1, 0) |
| allergic_conjunctivitis_or_rhinitis_or_hay_fever_cum_4y | Past medical history of allergic conjunctivitis or rhinitis or hay fever | C1Y_0112501, C1hY_0090301, C2Y_0060301, C2Y_0060401, C3Y_0091301, C3Y_0091501, C4Y_0051301, C4Y_0051501 | ifelse(any(source variables == 1), 1, 0) |
| otitis_media_cum_2y | Past medical history of otitis media | C6m_0110201, C1Y_0115301, C1hY_0092001, C2Y_0061301 | ifelse(any(source variables == 1), 1, 0) |
| upper_respiratory_tract_infection_cum_2y | Past medical history of upper respiratory infection | C6m_0110301, C1Y_0115401, C1hY_0092101, C2Y_0061401 | ifelse(any(source variables == 1), 1, 0) |
| lower_respiratory_tract_infection_cum_2y | Past medical history of lower respiratory infection | C6m_0110401, C1Y_0115501, C1hY_0092201, C2Y_0061501 | ifelse(any(source variables == 1), 1, 0) |
| gastroenteritis_cum_2y | Past medical history of gastroenteritis | C6m_0111101, C1Y_0115601, C1hY_0092301, C2Y_0061601 | ifelse(any(source variables == 1), 1, 0) |
| roseola_infantum_cum_2y | Past medical history of roseola infantum | C6m_0110601, C1Y_0116001, C1hY_0092701, C2Y_0062001 | ifelse(any(source variables == 1), 1, 0) |
| herpangina_cum_2y | Past medical history of herpangina | C6m_0110801, C1Y_0116101, C1hY_0092801, C2Y_0062101 | ifelse(any(source variables == 1), 1, 0) |
| hand_foot_and_mouth_disease_cum_2y | Past medical history of hand foot and mouth disease | C6m_0110901, C1Y_0116201, C1hY_0092901, C2Y_0062201 | ifelse(any(source variables == 1), 1, 0) |
| adenovirus_infection_cum_2y | Past medical history of adenovirus infection | C1Y_0116301, C1hY_0093001, C2Y_0062301 | ifelse(any(source variables == 1), 1, 0) |
| rsv_infection_cum_2y | Past medical history of RSV infection | C6m_0110501, C1Y_0116401, C1hY_0093101, C2Y_0062401 | ifelse(any(source variables == 1), 1, 0) |
| hsv_infection_cum_2y | Past medical history of HSV infection | C6m_0111301, C1Y_0116501, C1hY_0093201, C2Y_0062501 | ifelse(any(source variables == 1), 1, 0) |
| mycotic_or_fungal_infection_cum_2y | Past medical history of mycotic or fungal infection | C6m_0111201, C1Y_0116601, C1hY_0093301, C2Y_0062601 | ifelse(any(source variables == 1), 1, 0) |
| urinary_tract_infections_cum_4y | Past medical history of urinary tract infection | C1Y_0115701, C1hY_0092401, C2Y_0061701, C3Y_0092701, C4Y_0053701 | ifelse(any(source variables == 1), 1, 0) |
| varicella_cum_4y | Past medical history of varicella | C1Y_0116701, C2Y_0062701, C3Y_0092801, C4Y_0053801 | ifelse(any(source variables == 1), 1, 0) |
| influenza_viral_infections_cum_4y | Past medical history of influenza viral infection | C6m_0111001, C1Y_0115901, C1hY_0092601, C2Y_0061901, C3Y_0092901, C4Y_0053901 | ifelse(any(source variables == 1), 1, 0) |
| streptococcus_pyogenes_cum_4y | Past medical history of Streptococcus pyogenes infection | C3Y_0093001, C4Y_0054001 | ifelse(any(source variables == 1), 1, 0) |
| mumps_cum_4y | Past medical history of mumps | C6m_0110701, C1Y_0117001, C2Y_0063001, C3Y_0093401, C4Y_0054401 | ifelse(any(source variables == 1), 1, 0) |
| intellectual_and_developmental_disability_including_language_delay_cum_4y | Past medical history of intellectual and developmental disability including language delay | C3Y_0095001, C4Y_0056001 | ifelse(any(source variables == 1), 1, 0) |
| psychomotor_developmental_delay_cum_4y | Past medical history of psychomotor developmental delay | C1Y_0113001, C1hY_0091001, C3Y_0094901, C3Y_0095001, C4Y_0055901, C4Y_0056001 | ifelse(any(source variables == 1), 1, 0) |
| autistic_spectrum_disorder_and_pddnos_cum_4y | Past medical history of autistic spectrum disorder and PDDNOS | C3Y_0095101, C4Y_0056101 | ifelse(any(source variables == 1), 1, 0) |
| bone_fracture_cum_4y | Past medical history of bone fracture | C3Y_0095501, C4Y_0056501 | ifelse(any(source variables == 1), 1, 0) |
| fever_count_cum_2hy | Total number of fever episodes over 38 degrees Celsius up to 2.5 years old | C1Y_0090001, C2Y_0070001, C2hY_0040101 | sum(all source variables) |
| fever_count_cum_2y | Total number of fever episodes over 38 degrees Celsius up to 2 years old | C1Y_0090001, C2Y_0070001 | sum(all source variables) |

| | | | |
|---|---|---|---|
| fever_over_39c_count_cum_2hy | Total number of fever episodes over 39 degrees Celsius up to 2.5 years old | C1Y_0090002, C2Y_0070002, C2hY_0040102 | sum(all source variables) |
| fever_over_39c_count_cum_2y | Total number of fever episodes over 39 degrees Celsius up to 2 years old | C1Y_0090002, C2Y_0070002 | sum(all source variables) |
| constipation_severity_3y | Severity of constipation at 3 years old | C3Y_0270101, C3Y_0270201, C3Y_0270301, C3Y_0270401, C3Y_0270501, C3Y_0270601 | sum(all source variables) |
| constipation_severity_4y | Severity of constipation at 4 years old | C4Y_0300101, C4Y_0300201, C4Y_0300301, C4Y_0300401, C4Y_0300501, C4Y_0300601 | sum(all source variables) |
| constipation_cum_4y | History of constipation up to 4 years old | C3Y_0270101, C3Y_0270201, C3Y_0270301, C3Y_0270401, C3Y_0270501, C3Y_0270601, C4Y_0300101, C4Y_0300201, C4Y_0300301, C4Y_0300401, C4Y_0300501, C4Y_0300601 | ifelse(sum(C3Y_0270101:C3Y_0270601) >= 2 \| sum(C4Y_0300101:C4Y_0300601) >= 2, 1, 0) |

##### Eczema-related trait

This trait captures a broader history of eczema at 1 year of age by combining questionnaire responses indicating either diagnosed eczema or eczema symptoms without a formal diagnosis. It was defined to represent overall eczema history rather than diagnostic status alone.

| Trait_ID | Description | Source variables | Definition |
|---|---|---|---|
| c1y_0170001_1_3 | History of eczema including undiagnosed cases at 1 year old | C1Y_0170001 | ifelse(source variable == 1 \| source variable == 3, 1, 0) |

##### Sleep-related traits

Sleep-related traits capture multiple dimensions of infant and early childhood sleep behavior, including total sleep duration, daytime and nighttime sleep distribution, nap frequency, and patterns of nighttime awakenings, bedtime, and wake-up timing. Because sleep is closely linked to brain maturation and behavioral regulation, analyzing these traits across ages allows us to characterize developmental changes in sleep–wake regulation and to identify atypical or disrupted sleep patterns that may be relevant for later neurodevelopmental outcomes. Here we introduce 35 traits composed of the questionnaires about sleeping.

| Trait_ID | Description | Source variables | Definition |
|---|---|---|---|
| m1m_total_sleep_time | Total sleep duration per day at 1 month old | M1m_0270001–M1m_0270024, M1m_0270101–M1m_0270124 | 30 * sum(all source variables) |
| c6m_total_sleep_time | Total sleep duration per day at 6 months old | C6m_0200001–C6m_0200024, C6m_0200101–C6m_0200124 | 30 * sum(all source variables) |
| c1y_total_sleep_time | Total sleep duration per day at 1 year old | C1Y_0240001–C1Y_0240024, C1Y_0240101–C1Y_0240124 | 30 * sum(all source variables) |
| c1hy_total_sleep_time | Total sleep duration per day at 1.5 years old | C1hY_0270001–C1hY_0270024, C1hY_0270101–C1hY_0270124 | 30 * sum(all source variables) |
| c3y_total_sleep_time | Total sleep duration per day at 3 years old | C3Y_0070001–C3Y_0070024, C3Y_0070101–C3Y_0070124 | 30 * sum(all source variables) |
| m1m_sleep_time_day | Total daytime sleep time (7:00 am–7:00 pm) at 1 month old | M1m_0270015–M1m_0270024, M1m_0270101–M1m_0270114 | 30 * sum(all source variables) |
| c6m_sleep_time_day | Total daytime sleep time (7:00 am–7:00 pm) at 6 months old | C6m_0200015–C6m_0200024, C6m_0200101–C6m_0200114 | 30 * sum(all source variables) |
| c1y_sleep_time_day | Total daytime sleep time (7:00 am–7:00 pm) at 1 year old | C1Y_0240015–C1Y_0240024, C1Y_0240101–C1Y_0240114 | 30 * sum(all source variables) |
| c1hy_sleep_time_day | Total daytime sleep time (7:00 am–7:00 pm) at 1.5 years old | C1hY_0270015–C1hY_0270024, C1hY_0270101–C1hY_0270114 | 30 * sum(all source variables) |
| c3y_sleep_time_day | Total daytime sleep time (7:00 am–7:00 pm) at 3 years old | C3Y_0070015–C3Y_0070024, C3Y_0070101–C3Y_0070114 | 30 * sum(all source variables) |
| m1m_sleep_time_night | Total nighttime sleep duration (7:00 pm–7:00 am) at 1 | M1m_0270001–M1m_0270014, | 30 * sum(all source variables) |

| | | | |
|---|---|---|---|
| | month old | M1m_0270115–M1m_0270124 | |
| c6m_sleep_time_night | Total nighttime sleep duration (7:00 pm–7:00 am) at 6 months old | C6m_0200001–C6m_0200014, C6m_0200115–C6m_0200124 | 30 * sum(all source variables) |
| c1y_sleep_time_night | Total nighttime sleep duration (7:00 pm–7:00 am) at 1 year old | C1Y_0240001–C1Y_0240014, C1Y_0240115–C1Y_0240124 | 30 * sum(all source variables) |
| c1hy_sleep_time_night | Total nighttime sleep duration (7:00 pm–7:00 am) at 1.5 years old | C1hY_0270001–C1hY_0270014, C1hY_0270115–C1hY_0270124 | 30 * sum(all source variables) |
| c3y_sleep_time_night | Total nighttime sleep duration (7:00 pm–7:00 am) at 3 years old | C3Y_0070001–C3Y_0070014, C3Y_0070115–C3Y_0070124 | 30 * sum(all source variables) |
| m1m_nap_count | Number of naps during the day (7:00 am–7:00 pm) at 1 month old | M1m_0270014–M1m_0270024, M1m_0270101–M1m_0270114 | sum(diff(x) == 1) |
| c6m_nap_count | Number of naps during the day (7:00 am–7:00 pm) at 6 months old | C6m_0200014–C6m_0200024, C6m_0200101–C6m_0200114 | sum(diff(x) == 1) |
| c1y_nap_count | Number of naps during the day (7:00 am–7:00 pm) at 1 year old | C1Y_0240014–C1Y_0240024, C1Y_0240101–C1Y_0240114 | sum(diff(x) == 1) |
| c1hy_nap_count | Number of naps during the day (7:00 am–7:00 pm) at 1.5 years old | C1hY_0270014–C1hY_0270024, C1hY_0270101–C1hY_0270114 | sum(diff(x) == 1) |
| c3y_nap_count | Number of naps during the day (7:00 am–7:00 pm) at 3 years old | C3Y_0070014–C3Y_0070024, C3Y_0070101–C3Y_0070114 | sum(diff(x) == 1) |
| m1m_night_wake_up_count | Number of nighttime awakenings (7:00 pm–7:00 am) at 1 month old | M1m_0270114–M1m_0270124, M1m_0270001–M1m_0270014 | sum(diff(source variables) == -1) |
| c6m_night_wake_up_count | Number of nighttime awakenings (7:00 pm–7:00 am) at 6 months old | C6m_0200114–C6m_0200124, C6m_0200001–C6m_0200014 | sum(diff(source variables) == -1) |
| c1y_night_wake_up_count | Number of nighttime awakenings (7:00 pm–7:00 am) at 1 year old | C1Y_0240114–C1Y_0240124, C1Y_0240001–C1Y_0240014 | sum(diff(source variables) == -1) |
| c1hy_night_wake_up_count | Number of nighttime awakenings (7:00 pm–7:00 am) at 1.5 years old | C1hY_0270114–C1hY_0270124, C1hY_0270001–C1hY_0270014 | sum(diff(source variables) == -1) |
| c3y_night_wake_up_count | Number of nighttime awakenings (7:00 pm–7:00 am) at 3 years old | C3Y_0070114–C3Y_0070124, C3Y_0070001–C3Y_0070014 | sum(diff(source variables) == -1) |
| m1m_bed_time | Bedtime at 1 month old | M1m_0270111–M1m_0270124, M1m_0270001–M1m_0270024, M1m_0270101–M1m_0270110 | min(i such that four consecutive source variables == 1) |
| c6m_bed_time | Bedtime at 6 months old | C6m_0200111–C6m_0200124, C6m_0200001–C6m_0200024, C6m_0200101–C6m_0200110 | min(i such that four consecutive source variables == 1) |
| c1y_bed_time | Bedtime at 1 year old | C1Y_0240111–C1Y_0240124, C1Y_0240001–C1Y_0240024, C1Y_0240101–C1Y_0240110 | min(i such that four consecutive source variables == 1) |
| c1hy_bed_time | Bedtime at 1.5 years old | C1hY_0270111–C1hY_0270124, C1hY_0270001–C1hY_0270024, C1hY_0270101–C1hY_0270110 | min(i such that four consecutive source variables == 1) |
| c3y_bed_time | Bedtime at 3 years old | C3Y_0070111–C3Y_0070124, C3Y_0070001–C3Y_0070024, C3Y_0070101–C3Y_0070110 | min(i such that four consecutive source variables == 1) |
| m1m_wake_up_time | Wake-up time at 1 month old | M1m_0270011–M1m_0270024, M1m_0270101–M1m_0270124, M1m_0270001–M1m_0270010 | min(i such that four consecutive source variables sum to 0) |
| c6m_wake_up_time | Wake-up time at 6 months old | C6m_0200011–C6m_0200024, C6m_0200101–C6m_0200124, C6m_0200001–C6m_0200010 | min(i such that four consecutive source variables sum to 0) |
| c1y_wake_up_time | Wake-up time at 1 year old | C1Y_0240011–C1Y_0240024, C1Y_0240101–C1Y_0240124, C1Y_0240001–C1Y_0240010 | min(i such that four consecutive source variables sum to 0) |
| c1hy_wake_up_time | Wake-up time at 1.5 years old | C1hY_0270011–C1hY_0270024, C1hY_0270101–C1hY_0270124, C1hY_0270001–C1hY_0270010 | min(i such that four consecutive source variables sum to 0) |
| c3y_wake_up_time | Wake-up time at 3 years old | C3Y_0070011–C3Y_0070024, C3Y_0070101–C3Y_0070124, C3Y_0070001–C3Y_0070010 | min(i such that four consecutive source variables sum to 0) |

##### Childcare facility use-related traits

These traits describe the child's exposure to group childcare environments, including the frequency of use and the age at which childcare was initiated. Such traits support analyses of early social environmental influences, exposure to group settings, and the potential developmental or behavioral implications associated with childcare use. Here we introduce 3 traits derived from questionnaire variables related to childcare facility use.

| Trait_ID | Description | Source variables | Definition |
|---|---|---|---|
| childcare_facility_daycare_time_3y | Frequency of group childcare use at 3 years old | C3Y_0350201, C3Y_0350301 | C3Y_0350301 * C3Y_0350201 |
| childcare_facility_daycare_time_4y | Frequency of group childcare use at 4 years old | C4Y_0380201, C4Y_0380301 | C4Y_0380301 * C4Y_0380201 |
| childcare_facility_starting_age | Age at starting group childcare | C3Y_0350101, C3Y_0350102, C2Y_0360101, C2Y_0360102 | min(C3Y_0350101 * 12 + C3Y_0350102, C2Y_0360101 * 12 + C2Y_0360102), using the non-missing value when only one is available |

##### Toilet training-related trait

This trait captures the age at which night diaper use was discontinued by 4 years of age. Because the original questionnaire records this information separately in years and months, the trait was standardized into a single continuous measure expressed in months. This harmonization simplifies downstream analyses while preserving the original timing information.

| Trait_ID | Description | Source variables | Definition |
|---|---|---|---|
| out_of_night_diapers_age_4y | Age at stopping use of night diapers by 4 years old | C4Y_0290101, C4Y_0290102 | C4Y_0290101 * 12 + C4Y_0290102 |

##### ASQ-3 questionnaire-related traits

Interpreting ASQ-3 scores in an integrated, multidomain manner provides a more comprehensive understanding of a child's developmental status. Because developmental domains are interrelated, a composite interpretation may help identify broader patterns and detect subtle delays that may not be evident within a single domain. Here we introduce 8 traits representing the total ASQ-3 score at different ages.

| Trait_ID | Description | Source variables | Definition |
|---|---|---|---|
| c6m_ASQ_total | Total score of ASQ-3 at 6 months old | C6M_ASQ_aComu, C6M_ASQ_bGM, C6M_ASQ_cFM, C6M_ASQ_dProblem, C6M_ASQ_ePersonal | sum(all source variables) |
| c1y_ASQ_total | Total score of ASQ-3 at 1 year old | C1Y_ASQ_aComu, C1Y_ASQ_bGM, C1Y_ASQ_cFM, C1Y_ASQ_dProblem, C1Y_ASQ_ePersonal | sum(all source variables) |
| c1hy_ASQ_total | Total score of ASQ-3 at 1.5 years old | C1hY_ASQ_aComu, C1hY_ASQ_bGM, C1hY_ASQ_cFM, C1hY_ASQ_dProblem, C1hY_ASQ_ePersonal | sum(all source variables) |
| c2y_ASQ_total | Total score of ASQ-3 at 2 years old | C2Y_ASQ_aComu, C2Y_ASQ_bGM, C2Y_ASQ_cFM, C2Y_ASQ_dProblem, C2Y_ASQ_ePersonal | sum(all source variables) |
| c2hy_ASQ_total | Total score of ASQ-3 at 2.5 years old | C2hY_ASQ_aComu, C2hY_ASQ_bGM, C2hY_ASQ_cFM, C2hY_ASQ_dProblem, C2hY_ASQ_ePersonal | sum(all source variables) |
| c3y_ASQ_total | Total score of ASQ-3 at 3 years old | C3Y_ASQ_aComu, C3Y_ASQ_bGM, C3Y_ASQ_cFM, C3Y_ASQ_dProblem, C3Y_ASQ_ePersonal | sum(all source variables) |
| c3hy_ASQ_total | Total score of ASQ-3 at 3.5 years old | C3hY_ASQ_aComu, C3hY_ASQ_bGM, C3hY_ASQ_cFM, C3hY_ASQ_dProblem, C3hY_ASQ_ePersonal | sum(all source variables) |
| c4y_ASQ_total | Total score of ASQ-3 at 4 years old | C4Y_ASQ_aComu, C4Y_ASQ_bGM, C4Y_ASQ_cFM, C4Y_ASQ_dProblem, C4Y_ASQ_ePersonal | sum(all source variables) |

### 5. Trait selection for GWAS

When defining the set of traits for the GWAS, we considered several traits unsuitable because they had too many missing responses (low call rate) or were too low in complexity (*i.e.* too many identical values/answers in the trait), leading to high false positives. We calculated the call rate of a trait, such as

```
(call rate) = 1.0 - (number of valid data points without missing values) / (number of questionnaires collected at the given age of the children),
```

and excluded traits with more than 40% missing responses from the analysis. The complexity score, defined as

```
(complexity) = 1.0 - (frequency of the most frequent value in the trait) / (number of valid data points),
```

was calculated for each trait (note that this score should be the case ratio for a case-control trait). We removed binary traits with a score of less than 0.01 and quantitative traits with a score of less than 0.05 to reduce potential false positives in GWAS. A total of 1,255 parental and child traits were analyzed in the GWAS.
