## Supplementary Notes 2 for "Genome-wide association study on longitudinal and cross-sectional traits of child health and development in a Japanese population"

### 1 Association mapping using Gaussian process regression model

At the beginning, we would clarify that the Gaussian-process (GP) regression model in this manuscript builds on the same GP-based genetic-association framework as GASPACHO (Kumasaka et al., 2023), but with important differences in the data structure and implementation. GASPACHO was developed for single-cell eQTL mapping, where latent cellular states are inferred from high-dimensional gene-expression data. In the present study, this latent-space inference step is unnecessary because the relevant longitudinal coordinate, child age, is directly observed. In addition, we improved the computational implementation so that the model can be applied to biobank-scale longitudinal cohort data with tens of thousands of participants and repeated measurements, while explicitly accounting for familial relatedness using KING-derived relatedness information. In contrast, the original GASPACHO study was applied to a much smaller set of unrelated donors (N=68).

We first consider the null model in which no genetic effect exists. We model a quantitative trait $y \in \mathbb{R}^N$ observed at the time point $x \in \mathbb{R}^N$ using a Gaussian process (GP) regression model as follows:

$$
\begin{aligned}
y|a,b,f,h_1,\ldots,h_{N_d} &\sim \mathcal{N}\left(Xa + Zb + f + \sum_{i=1}^{N_d} h_i, \sigma^2 I\right), \qquad (1)\\
a &\sim \mathcal{N}(0, \sigma^2\Delta)\\
b &\sim \mathcal{N}(0, \sigma^2\delta_d^2 I)\\
f|u &\sim \mathcal{N}(K_{NM}K_{MM}^{-1}u, \sigma^2\delta_c^2\tilde{K}_{NN}),\\
u &\sim \mathcal{N}(0, \sigma^2\delta_c^2 K_{MM}),\\
h_i|v_i &\sim \mathcal{N}(K_{NM}K_{MM}^{-1}v_i, \sigma^2\delta_{d\times c}^2\tilde{K}_{NN}),\\
v_i &\sim \mathcal{N}(0, \sigma^2\delta_{d\times c}^2 K_{MM}).
\end{aligned}
$$

where $X \in \mathbb{R}^{N\times P}$ is a matrix of the known covariates, $Z \in \mathbb{R}^{N\times N_d}$ is an indicator matrix of $N_d$ donors. We also denote the variance parameters $\delta_d^2$, $\delta_c^2$ and $\delta_{d\times c}^2$ for donor, context and donor-by-context, respectively. The residual noise is given by $\sigma^2$. Here we introduce $M$ inducing points $z$ to reduce the complexity of the model so that

$$
\begin{aligned}
K_{NN} &= \left(\exp\left\{-\frac{(x_i - x_j)^2}{\rho}\right\}; 1 \le i,j \le N\right),\\
K_{NM} &= \left(\exp\left\{-\frac{(x_i - z_j)^2}{\rho}\right\}; 1 \le i \le N, 1 \le j \le M\right),\\
K_{MM} &= \left(\exp\left\{-\frac{(z_i - z_j)^2}{\rho}\right\}; 1 \le i,j \le M\right).
\end{aligned}
$$

with $\tilde{K}_{NN} = K_{NN} - K_{NM}K_{MM}^{-1}K_{NM}^{\top}$.

We then have a lower bound by exchanging the integral and log as

$$
\begin{aligned}
\log p(y|u, v_1, \ldots, v_{N_d}) &= \log \int p(y|Xa + Zb + f + \sum_i h_i \odot z_i, \sigma^2 I) p(f|u) p(h_i|v_i) df \prod_i dh_i\\
&\ge \log p(y|Xa + Zb + \bar{f} + \sum_i \bar{h}_i \odot z_i, \sigma^2 I) - \frac{1}{2}\mathrm{tr}\{(\delta_c^2 + \delta_{d\times c}^2)\tilde{K}_{NN}\}\\
&\equiv \mathcal{L}_1
\end{aligned}
$$

where $\bar{f} = K_{NM}K_{MM}^{-1}u$ and $\bar{h}_i = K_{NM}K_{MM}^{-1}v_i$, $i = 1, \ldots, N_d$,which leads to the Titsias lower bound

Titsias (2009):

$$
\begin{aligned}
\log p(y) &\geq \log \int \exp\{\mathcal{L}_1\} p(u) \prod_i p(v_i) du \prod_i dv_i \\
&= \log p(y|0, \sigma^2 V) - \frac{1}{2}\mathrm{tr}\{(\delta_c^2 + \delta_{d\times c}^2)\tilde{K}_{NN}\} \\
&= const - \frac{N}{2}\log\sigma^2 - \frac{1}{2}\log|V| - \frac{1}{2\sigma^2} y^\top V^{-1} y - \frac{1}{2}(\delta_c^2 + \delta_{d\times c}^2)\mathrm{tr}\{\tilde{K}_{NN}\} \\
&\equiv \mathcal{L}_2
\end{aligned}
$$

where

$$
\begin{aligned}
V &= I + X\Delta X^\top + \delta_c^2 K_{NM}K_{MM}^{-1}K_{MN} + \delta_d^2 ZZ^\top + \delta_{d\times c}^2 (K_{NM}K_{MM}^{-1}K_{MN}) \odot (ZZ^\top) \\
&\equiv I + \tilde{Z}\tilde{\Delta}\tilde{Z}^\top
\end{aligned}
$$

and

$$
\begin{aligned}
\tilde{Z} &= (X, \tilde{K}_{NM}, \tilde{K}_{NM}^{(1)}, \ldots, \tilde{K}_{NM}^{(N_d)}), \\
\tilde{\Delta} &= \begin{pmatrix} \Delta & 0 & 0 & \cdots & 0 \\ 0 & \delta_c^2 I_M & 0 & \cdots & 0 \\ 0 & 0 & \Delta_d & \cdots & 0 \\ \vdots & \vdots & \vdots & \ddots & \vdots \\ 0 & 0 & 0 & \cdots & \Delta_d \end{pmatrix}
\end{aligned}
$$

with

$$
\begin{aligned}
\tilde{K}_{NM}^{(i)} &= (z_i 1_{M+1}^\top) \odot (\tilde{K}_{NM}, z_i), \\
\tilde{K}_{NM} &= K_{NM} R^{-1}, \\
K_{MM} &= R^\top R, \\
\Delta_d &= \begin{pmatrix} \delta_{d\times c}^2 I_M & 0 \\ 0 & \delta_d^2 \end{pmatrix}.
\end{aligned}
$$

To compute $V^{-1} = I - \tilde{Z}\Phi\tilde{Z}^\top$, we introduce

$$
\begin{aligned}
\Phi^{-1} &= \tilde{\Delta}^{-1} + \tilde{Z}^\top \tilde{Z} \\
&= \left(\begin{array}{cc|ccc} \Delta^{-1} + X^\top X & X^\top \tilde{K}_{NM} & X^\top \tilde{K}_{NM}^{(1)} & \cdots & X^\top \tilde{K}_{NM}^{(N_d)} \\ K_{MN} X & \delta_c^{-2} I_M + \tilde{K}_{MN}\tilde{K}_{NM} & \tilde{K}_{MN}\tilde{K}_{NM}^{(1)} & \cdots & \tilde{K}_{MN}\tilde{K}_{NM}^{(N_d)} \\ \hline \tilde{K}_{MN}^{(1)} X & \tilde{K}_{MN}^{(1)}\tilde{K}_{NM} & \Delta_d^{-1} + \tilde{K}_{MN}^{(1)}\tilde{K}_{NM}^{(1)} & \cdots & 0 \\ \vdots & \vdots & \vdots & \ddots & \vdots \\ \tilde{K}_{MN}^{(N_d)} X & \tilde{K}_{MN}^{(N_d)}\tilde{K}_{NM} & 0 & \cdots & \Delta_d^{-1} + \tilde{K}_{MN}^{(N_d)}\tilde{K}_{NM}^{(N_d)} \end{array}\right) \\
&\equiv \begin{pmatrix} A & B^\top \\ B & C \end{pmatrix}.
\end{aligned}
$$

This gives

$$
\Phi = \begin{pmatrix} A & B^\top \\ B & C \end{pmatrix}^{-1} = \begin{pmatrix} D^{-1} & -D^{-1}B^\top C^{-1} \\ -C^{-1}BD^{-1} & C^{-1} + C^{-1}BD^{-1}B^\top C^{-1} \end{pmatrix},
$$

where $D = A - B^\top C^{-1} B$. To further compute

$$\begin{aligned} |V| &= |I + \tilde{Z}^\top \tilde{Z} \tilde{\Delta}| \\ &= |\tilde{\Delta}||\Phi^{-1}|, \end{aligned}$$

we can use

$$\left| \begin{array}{cc} A & B^\top \\ B & C \end{array} \right| = |C||A - B^\top C^{-1} B| = |C||D|.$$

It is also noting that

$$\begin{aligned} V^{-1}\tilde{Z} &= \tilde{Z}\Phi\tilde{\Delta}^{-1}, \\ \tilde{Z}^\top V^{-1}\tilde{Z} &= \tilde{\Delta}^{-1}(\tilde{\Delta} - \Phi)\tilde{\Delta}^{-1}. \end{aligned}$$

#### 1.1 Estimating $\rho$

To estimate the length parameter $\rho$ in the exponential kernel, we use the following reduced model (Step 1):

$$\begin{aligned} y|a, f &\sim \mathcal{N}(Xa + f, \sigma^2 I), \\ a &\sim \mathcal{N}(0, \sigma^2 \Delta), \\ f|u &\sim \mathcal{N}(K_{NM}K_{MM}^{-1}u, \sigma^2\delta_c^2\tilde{K}_{NN}), \\ u &\sim \mathcal{N}(0, \sigma^2\delta_c^2 K_{MM}). \end{aligned} \tag{2}$$

We obtain the Titsias lower bound:

$$\begin{aligned} \log p(y|a,u) &= \log \int p(y|Xa + f, \sigma^2 I) p(f|u) \\ &\geq \log p(y|Xa + \overline{f}, \sigma^2 I) - \frac{1}{2}\mathrm{tr}\{\delta_c^2 \tilde{K}_{NN}\} \\ &\equiv \mathcal{L}_1, \\ \log p(y) &\geq \log \int \exp\{\mathcal{L}_1\} p(u) \\ &= \log p(y|0, \sigma^2 V) - \frac{1}{2}\mathrm{tr}\{\delta_c^2 \tilde{K}_{NN}\} \\ &= const - \frac{N}{2}\log\sigma^2 - \frac{1}{2}\log|V| - \frac{1}{2\sigma^2} y^\top V^{-1} y - \frac{1}{2}\delta_c^2 \mathrm{tr}\{\tilde{K}_{NN}\}, \\ &\equiv \mathcal{L}_2, \end{aligned}$$

where

$$\begin{aligned} V &= \sigma^2(I + \tilde{Z}\tilde{\Delta}\tilde{Z}^\top), \\ \tilde{Z} &= (X, K_{NM}), \\ \tilde{\Delta} &= \left( \begin{array}{cc} \Delta^{-1} & 0 \\ 0 & \delta_c^2 K_{MM}^{-1} \end{array} \right). \end{aligned}$$

We then obtain

$$\begin{aligned}
\mathrm{tr}\{V^{-1}\partial_\rho V\} &= 2\mathrm{tr}\{\tilde{\Delta}\tilde{Z}^\top V^{-1}(0, \partial K_{NM})\} + \mathrm{tr}\{V^{-1}\tilde{Z}\tilde{\Delta}\mathrm{diag}(0, -\delta_c^{-2}\partial K_{MM})\tilde{\Delta}\tilde{Z}^\top\} \\
&= 2\mathrm{tr}\{\Phi\tilde{Z}^\top(0, \partial K_{NM})\} - \mathrm{tr}\{(\tilde{\Delta} - \Phi)\mathrm{diag}(0, \delta_c^{-2}\partial K_{MM})\}, \\
y^\top V^{-1}(\partial_\rho V)V^{-1}y &= 2\mathrm{tr}\{\tilde{\Delta}\tilde{Z}^\top V^{-1}yy^\top V^{-1}(0, \partial K_{NM})\} + \mathrm{tr}\{\tilde{\Delta}\tilde{Z}^\top V^{-1}yy^\top V^{-1}\tilde{Z}\tilde{\Delta}\mathrm{diag}(0, -\delta_c^{-2}\partial K_{MM})\}, \\
&= 2\mathrm{tr}\{\Phi\tilde{Z}^\top yy^\top(I - \tilde{Z}\Phi\tilde{Z}^\top)(0, \partial K_{NM})\} - \mathrm{tr}\{\Phi\tilde{Z}^\top yy^\top\tilde{Z}\Phi\mathrm{diag}(0, \delta_c^{-2}\partial K_{MM})\} \\
\partial_\rho\mathrm{tr}\{\tilde{K}_{NN}\} &= -2\mathrm{tr}\{K_{MM}^{-1}K_{MN}(\partial K_{NM})\} + \mathrm{tr}\{K_{MM}^{-1}K_{MN}K_{NM}K_{MM}^{-1}(\partial K_{MM})\},
\end{aligned}$$

which gives

$$\begin{aligned}
\frac{\partial\mathcal{L}_2}{\partial\rho} &= -\mathrm{tr}\{\Phi\tilde{Z}^\top(0, \partial K_{NM})\} + \frac{1}{2}\mathrm{tr}\{(\tilde{\Delta} - \Phi)\mathrm{diag}(0, \delta_c^{-2}\partial K_{MM})\} \\
&\quad + \frac{1}{\sigma^2}\mathrm{tr}\{\Phi\tilde{Z}^\top yy^\top(I - \tilde{Z}\Phi\tilde{Z}^\top)(0, \partial K_{NM})\} - \frac{1}{2\sigma^2}\mathrm{tr}\{\Phi\tilde{Z}^\top yy^\top\tilde{Z}\Phi\mathrm{diag}(0, \delta_c^{-2}\partial K_{MM})\} \\
&\quad + \delta_c^2\mathrm{tr}\{K_{MM}^{-1}K_{MN}(\partial K_{NM})\} - \delta_c^2\mathrm{tr}\{K_{MM}^{-1}K_{MN}K_{NM}K_{MM}^{-1}(\partial K_{MM})\}/2.
\end{aligned}$$

We can use the quasi Newton method to obtain the maximum a posteriori (MAP) estimate of $\rho$.

In practice, the estimate can suffer from overfitting when the data contain multiple nonlinear patterns, such as a broad trend with small oscillations. To regularize this behavior, we place an Inverse-Gamma prior on $\rho$. Let $p(\rho)$ denote the probability density of $\rho$ under an Inverse-Gamma distribution with shape and scale parameters $\alpha$ and $\beta$, then,

$$\frac{\partial \log p(\rho)}{\partial\rho} = -\frac{\alpha + 1}{\rho} + \frac{\beta}{\rho^2},$$

which is added to $\partial\mathcal{L}_2/\partial\rho$ to obtain the MAP estimate. The shape and scale parameters were set to be $\alpha = 0.01N_d$ and $\beta = 5N_d$, suggesting the prior mean approaches 500 for sufficiently large $N_d$.

#### 1.2 Estimating $\tilde{\Delta}$

The variance parameter $\tilde{\Delta} = \mathrm{diag}(\Delta, \delta_c^2 I_M)$ is first estimated in conjunction with the length parameter $\rho$ with the reduced model (Eq. 2). Here we have the first derivative of $\mathcal{L}_2$ with respect to $\tilde{\Delta}$:

$$\begin{aligned}
\partial_{\tilde{\Delta}}\mathcal{L}_2 &= \frac{1}{2\sigma^2}y^\top V^{-1}(\partial V)V^{-1}y - \frac{1}{2}\mathrm{tr}\{V^{-1}(\partial V)\} \\
&= \frac{1}{2\sigma^2}y^\top V^{-1}\tilde{Z}(\partial\tilde{\Delta})\tilde{Z}^\top V^{-1}y - \frac{1}{2}\mathrm{tr}\{\tilde{Z}^\top V^{-1}\tilde{Z}(\partial\tilde{\Delta})\} \\
&= \frac{1}{2\sigma^2}y^\top\tilde{Z}\Phi\tilde{\Delta}^{-1}(\partial\tilde{\Delta})\tilde{\Delta}^{-1}\Phi\tilde{Z}^\top y - \frac{1}{2}\mathrm{tr}\{\tilde{\Delta}^{-1}(\tilde{\Delta} - \Phi)\tilde{\Delta}^{-1}(\partial\tilde{\Delta})\}.
\end{aligned}$$

We again use the quasi Newton method to obtain the MAP estimate of $\tilde{\Delta}$.

Once we obtained the MAP estimate of $\Delta$, $\delta_c^2$ and $\rho$, we fix these parameters and estimate the rest of variance parameters $\delta_{d\times c}^2$ and $\delta_d^2$ under the null model demonstrated in Eq.1 (Step 2). The first derivative is obtained by the above equation without the loss of generality.

#### 1.3 Score statistics

After fitting the null models in Steps 1 and 2, we performed genetic association mapping using score statistics (Step 3). For the genotype dosage vector $g \in \{0, 1, 2\}^{N_d}$, we now consider the full model to account for both static and dynamic genetic effect, $\alpha$ and $\beta$, such that

$$
\begin{aligned}
y|a,b,f,h_1,\ldots,h_{N_d},\alpha,\beta &\sim \mathcal{N}\left(Xa+Zb+f+\sum_{i=1}^{N_d} h_i+\alpha g^*+\beta\odot g^*,\sigma^2 I\right),\\
a &\sim \mathcal{N}(0,\sigma^2\Delta)\\
b &\sim \mathcal{N}(0,\sigma^2\delta_d^2 I)\\
f|u &\sim \mathcal{N}(K_{NM}K_{MM}^{-1}u,\sigma^2\delta_c^2\tilde{K}_{NN}),\\
u &\sim \mathcal{N}(0,\sigma^2\delta_c^2 K_{MM}),\\
h_i|v_i &\sim \mathcal{N}(K_{NM}K_{MM}^{-1}v_i,\sigma^2\delta_{d\times c}^2\tilde{K}_{NN}),\\
v_i &\sim \mathcal{N}(0,\sigma^2\delta_{d\times c}^2 K_{MM}),\\
\alpha &\sim \mathcal{N}(0,\sigma^2\delta_g^2),\\
\beta|w &\sim \mathcal{N}(K_{NM}K_{MM}^{-1}w,\sigma^2\delta_{g\times c}^2\tilde{K}_{NN}),\\
w &\sim \mathcal{N}(0,\sigma^2\delta_{g\times c}^2 K_{MM}),
\end{aligned}
$$

where $g^*=Zg$ is the expanded genotype vector. The Tistias lower bound can be calculated as follows:

$$
\begin{aligned}
\log p(y) &\geq \log\int \exp\{\mathcal{L}_1\}p(u)\prod_i p(v_i)du\prod_i dv_i dw\\
&= \log p(y|0,\sigma^2 V)-\frac{1}{2}\mathrm{tr}\{(\delta_c^2+\delta_{d\times c}^2+\delta_{g\times c}^2)\tilde{K}_{NN}\}\\
&= const-\frac{N}{2}\log\sigma^2-\frac{1}{2}\log|V|-\frac{1}{2\sigma^2}y^\top V^{-1}y-\frac{1}{2}(\delta_c^2+\delta_{d\times c}^2+\delta_{g\times c}^2)\mathrm{tr}\{\tilde{K}_{NN}\}\\
&\equiv \mathcal{L}_2
\end{aligned}
$$

The score like statistics is given by

$$
S=\frac{1}{\sigma^2}y^\top V^{-1}GG^\top V^{-1}y,
$$

where

$$
G=(g^*1_{M+1}^\top)\odot(\tilde{K}_{NM},1).
$$

The distribution of $S$ is the generalized $\chi^2$ distribution, that is, the distribution of the weighted sum of $M$ independent $\chi^2$ statistics, such as $\sum_{m=1}^{M}\lambda_m\chi_m^2$ (Cuomo et al., 2022; Kumasaka et al., 2023). It is known that the weights $\lambda_m$ ($m=1,\ldots,M$) are given by the non-negative eigenvalues of

$$
\begin{aligned}
U &= G^\top V^{-1}G\\
&= G^\top(I-\tilde{Z}\Phi\tilde{Z}^\top)G,
\end{aligned}
$$

where $\tilde{Z}=(X,\tilde{K}_{NM},\tilde{K}_{NM}^{(1)},\ldots,\tilde{K}_{NM}^{(N_d)})$. To compute $U$, we need

$$
\Phi\tilde{Z}^\top G=\begin{pmatrix} D^{-1}\tilde{X}^\top G-D^{-1}B^\top C^{-1}\tilde{K}_d^\top G\\ -C^{-1}BD^{-1}\tilde{X}_d^\top G+(C^{-1}+C^{-1}BD^{-1}B^\top C^{-1})\tilde{K}_d^\top G\end{pmatrix},
$$

where

$$
\begin{aligned}
\tilde{X} &= (X,\tilde{K}_{NM}),\\
\tilde{K}_d &= (\tilde{K}_{NM}^{(1)},\ldots,\tilde{K}_{NM}^{(N_d)}).
\end{aligned}
$$

To compute the $p$-value from $S$, we can use the `momentchi2` package on `R`.

##### 1.4 Posterior distribution

To estimate the genetic effects, we compute the posterior distribution

$$\begin{pmatrix} \hat{a} \\ \hat{u} \\ \hat{v}_1 \\ \hat{b}_1 \\ \vdots \\ \hat{v}_{N_d} \\ \hat{b}_{N_d} \\ \hat{w} \\ \hat{\alpha} \end{pmatrix} \sim \mathcal{N}(\tilde{\Phi}\tilde{Z}_g^\top y, \sigma^2\tilde{\Phi}),$$

where $\tilde{Z}_g^\top = (X, \tilde{K}_{NM}, \tilde{K}_{NM}^{(1)}, \ldots, \tilde{K}_{NM}^{(N_d)}, G)$ and

$$\tilde{\Phi} = \begin{pmatrix} \Phi^{-1} & \tilde{Z}^\top G \\ G^\top \tilde{Z} & \Phi_g^{-1} \end{pmatrix}^{-1}$$

with

$$\Delta_g = \begin{pmatrix} \delta_{g\times c}^2 I_M & 0 \\ 0 & \delta_g^2 \end{pmatrix},$$

$$\Phi_g^{-1} = \Delta_g^{-1} + G^\top G.$$

This gives

$$\begin{aligned}
\tilde{\Phi}\tilde{Z}_g^\top y &= \begin{pmatrix} \Phi^{-1} & \tilde{Z}^\top G \\ G^\top \tilde{Z} & \Phi_g^{-1} \end{pmatrix}^{-1} \begin{pmatrix} \tilde{Z}^\top y \\ G^\top y \end{pmatrix} \\
&= \begin{pmatrix} \Phi + \Phi\tilde{Z}^\top G D_g^{-1} G^\top \tilde{Z}\Phi & -\Phi\tilde{Z}^\top G D_g^{-1} \\ -D_g^{-1} G^\top \tilde{Z}\Phi & D_g^{-1} \end{pmatrix} \begin{pmatrix} \tilde{Z}^\top y \\ G^\top y \end{pmatrix} \\
&= \begin{pmatrix} \Phi\tilde{Z}^\top y + \Phi\tilde{Z}^\top G D_g^{-1} G^\top \tilde{Z}\Phi\tilde{Z}^\top y - \Phi\tilde{Z}^\top G D_g^{-1} G^\top y \\ -D_g^{-1} G^\top \tilde{Z}\Phi\tilde{Z}^\top y + D_g^{-1} G^\top y \end{pmatrix} \\
&= \begin{pmatrix} \Phi\tilde{Z}^\top y - \Phi\tilde{Z}^\top G D_g^{-1} G^\top V^{-1} y \\ D_g^{-1} G^\top V^{-1} y \end{pmatrix},
\end{aligned}$$

where $D_g = \Phi_g^{-1} - G^\top \tilde{Z}\Phi\tilde{Z}G = \Delta_g^{-1} + U$. The maximum likelihood estimator of $\sigma^2$ is given by

$$\hat{\sigma}^2 = \frac{y^\top V^{-1} y}{N}.$$

To estimate the genetic effects on $x^{(0)}$, we introduce $\tilde{K}_{N_0M} = (K_{N_0M}R^{-1}, 1)$, where

$$K_{N_0M} = \left( \exp\left\{ -\frac{(x_i^{(0)} - z_j)^2}{\rho} \right\}; 1 \le i \le N_0, 1 \le j \le M \right),$$

with which, we can calculate the effect

$$\hat{\beta}_{x^{(0)}} \sim \mathcal{N}(\tilde{K}_{N_0M}(\hat{w}^\top, \hat{\alpha}^\top)^\top, \hat{\sigma}^2 \tilde{K}_{N_0M} D_g \tilde{K}_{N_0M}^\top).$$

For example, we set $x^{(0)} = (0, \ldots, 54)^\top$ for the BMI study. Figure 1 (below) shows the posterior estimate of dynamic effects at variants discovered through static GWAS of BMI at 11 time points. Figure 2 shows the posterior estimate at variants discovered by both dynamic and static GWAS. Lastly Figure 3 shows the posterior estimate at variants discovered only by dynamic GWAS. For comparison, we also depicted the QTL effect sizes of static GWAS at the 11 time points.

#### 1.5 Dynamic polygenic score

Using the genome-wide association results, we can compute the dynamic polygenic score (PGS) for an individual based on their genotype data. Assuming that we have $L$ independent GWAS loci, we can choose the lead variant for the locus $l$ and estimate the effect $\hat{\beta}^{(l)}_{x^{(0)}}$ on $x^{(0)}$. Using the genotype data $g_i = (g_i^{(1)}, \ldots, g_i^{(L)})$ for the individual $i$ at the $L$ lead variants, we have the dynamic PGS, such as

$$PGS^{(i)}_{x^{(0)}} = \bar{f}_{x^{(0)}} + \sum \hat{\beta}^{(l)}_{x^{(0)}} g_i^{(l)},$$

where

$$\bar{f}_{x^{(0)}} = \frac{1}{L} \sum_{l=1}^{L} \tilde{K}_{N_0 M} \begin{pmatrix} \hat{u}^{(l)} \\ \hat{a}_1^{(l)} \end{pmatrix}$$

is the average context effect, estimated using the output $\hat{u}^{(l)}$ for the $M$ inducing points and the random intercept $\hat{a}^{(l)}$ for the variant $l$. We take the average because these estimators are not identical across $l$.

The prediction interval is given by

$$\mathrm{Var}\left(PGS^{(i)}_{x^{(0)}}\right) = \hat{\sigma}^2 \tilde{K}_{N_0 M} \left( \sum_{l=1}^{L} D_g^{(l)} \right) \tilde{K}_{N_0 M}^\top + \hat{\sigma}^2 I,$$

where $D_g^{(l)} = \Delta_g^{-1} + U^{(l)}$. Here $U^{(l)}$ is computed when the genetic association for the variant $l$ is tested (see the computation of weights for the generalized $\chi^2$ distribution).

#### 1.6 Incorporating kinship information

If the genotype data include closely related individuals, the test statistics may be inflated unless this relatedness is properly accounted for. To address this issue, we can extend the variance matrix as

$$V = I + X\Delta X^\top + \delta_c^2 K_{NM} K_{MM}^{-1} K_{MN} + \delta_d^2 Z A Z^\top + \delta_{d\times c}^2 (K_{NM} K_{MM}^{-1} K_{MN}) \odot (Z A Z^\top)$$

where $A$ denotes the additive relationship matrix, defined as twice the kinship matrix. Directly incorporating a dense matrix $A$ is computationally infeasible, because the computational complexity of inverting $V$ scales as $\mathcal{O}(N N_d^2 M^2)$.

Here, we assume that the kinship coefficient is approximately zero for pairs of individuals who are more distantly related than third-degree relatives. Under this assumption, if individuals are ordered such that related individuals are clustered together, the matrix $A$ can be represented as a block-diagonal matrix,

$$A = \begin{pmatrix} A_1 & \cdots & 0 \\ \vdots & \ddots & \vdots \\ 0 & \cdots & A_{N_f} \end{pmatrix}$$

where $A_i$ is the additive relationship matrix for the $i$th family cluster, and $N_f$ denotes the number of family clusters defined by 3rd-degree-or-closer relationships.

The Cholesky decomposition $A = LL^\top$ gives $ZAZ = (ZL)(ZL)^\top$. Because $A$ is block diagonal with respect to the family clusters, we obtain

$$ZL = \begin{pmatrix} Z_1 L_1 & \cdots & 0 \\ \vdots & \ddots & \vdots \\ 0 & \cdots & Z_{N_f} L_{N_f} \end{pmatrix},$$

where $Z_i$ $(i = 1, \ldots, N_f)$ is an indicator matrix assigning individuals to the members of family cluster $i$, and $L_i$ is the lower triangular matrix (*i.e.*, $A_i = L_i L_i^\top$). Thus,

$$Z_i L_i = \begin{pmatrix} l_{11}^{(i)} 1 & \cdots & 0 \\ \vdots & \ddots & \vdots \\ l_{n_i 1}^{(i)} 1 & \cdots & l_{n_i n_i}^{(i)} 1 \end{pmatrix},$$

where $l_{jk}^{(i)}$ denotes the $j, k$ element of $L_i$, $n_i$ denotes the number of family members in cluster $i$, and 1 denotes a vector of ones corresponding to individuals assigned to a given family member. Therefore, as long as $n_i$ remains modest for all family clusters, the computation of $V^{-1}$ remains tractable. In fact, in JECS, the maximum number of family members per cluster remains 7 when we define family clusters using a kinship-coefficient cutoff corresponding to third-degree relatives or closer.

Using the above block representation, we redefine

$$\tilde{K}_{NM}^{(i)} = \begin{pmatrix} \vdots & & \vdots \\ 0 & \cdots & 0 \\ l_{11}^{(i)} \tilde{K}_{NM}^{((i,1))} & \cdots & 0 \\ \vdots & \ddots & \vdots \\ l_{n_i 1}^{(i)} \tilde{K}_{NM}^{((i,n_i))} & \cdots & l_{n_i n_i}^{(i)} \tilde{K}_{NM}^{((i,n_i))} \\ 0 & \cdots & 0 \\ \vdots & & \vdots \end{pmatrix} \in \mathbb{R}^{N \times n_i(M+1)}$$

where $\tilde{K}_{NM}^{((i,j))}$ denotes the corresponding row slices of $\tilde{K}_{NM}$ corresponding to individual $j$ in family cluster $i$. With this notation, we further redefine

$$B = \begin{pmatrix} B^{(1)} \\ \vdots \\ B^{(N_f)} \end{pmatrix}, \qquad C = \begin{pmatrix} C_1 & \cdots & 0 \\ \vdots & \ddots & \vdots \\ 0 & \cdots & C_{N_f} \end{pmatrix},$$

where

$$B^{(i)} = \tilde{K}_{MN}^{((i))} \begin{pmatrix} X^{(i,1)} & \tilde{K}_{NM}^{((i,1))} \\ \vdots & \vdots \\ X^{(i,n_i)} & \tilde{K}_{NM}^{((i,n_i))} \end{pmatrix}, \qquad C_i = \begin{pmatrix} \Delta_d^{-1} & \cdots & 0 \\ \vdots & \ddots & \vdots \\ 0 & \cdots & \Delta_d^{-1} \end{pmatrix} + \tilde{K}_{MN}^{(i)} \tilde{K}_{NM}^{(i)},$$

and $\tilde{K}_{MN}^{((i))}$ denotes non-zero row slices of $\tilde{K}_{MN}^{(i)}$.

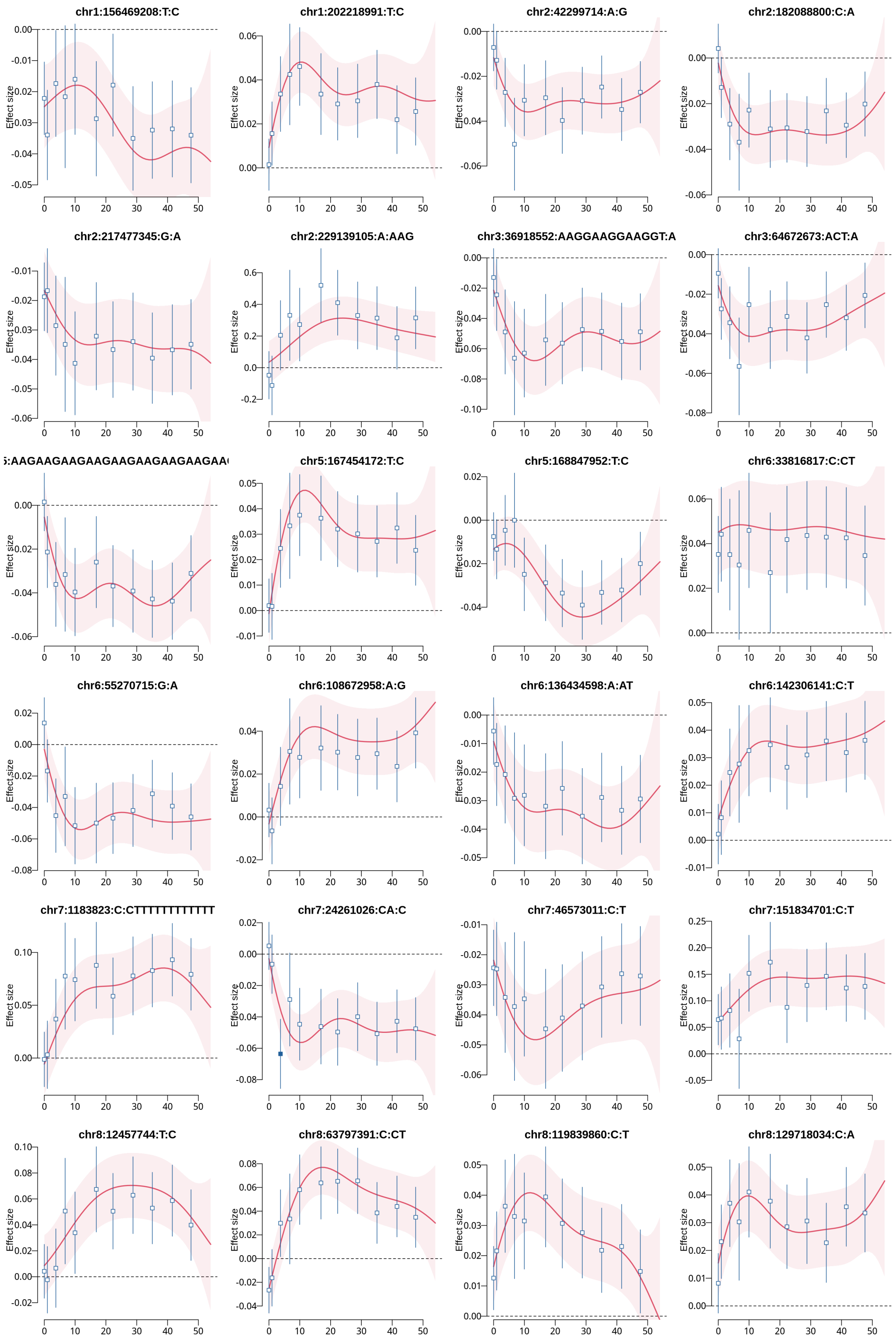


Figure 1. The longitudinal QTL effect and the cross-sectional QTL effects (detected only by longitudinal GWAS). The plot shows the dynamic genetic effect of the alternative allele relative to the reference allele, as estimated by Gaussian process regression over child age. The red line shows the posterior mean, and the shaded area shows the 95% credible interval of the longitudinal effect. The cross-sectional genetic effects, estimated by GWAS at 11 different age strata, are superimposed by blue square and segment as the 95% confidence interval (filled square: $P < 5 \times 10^{-8}$; open square $P \geq 5 \times 10^{-8}$).

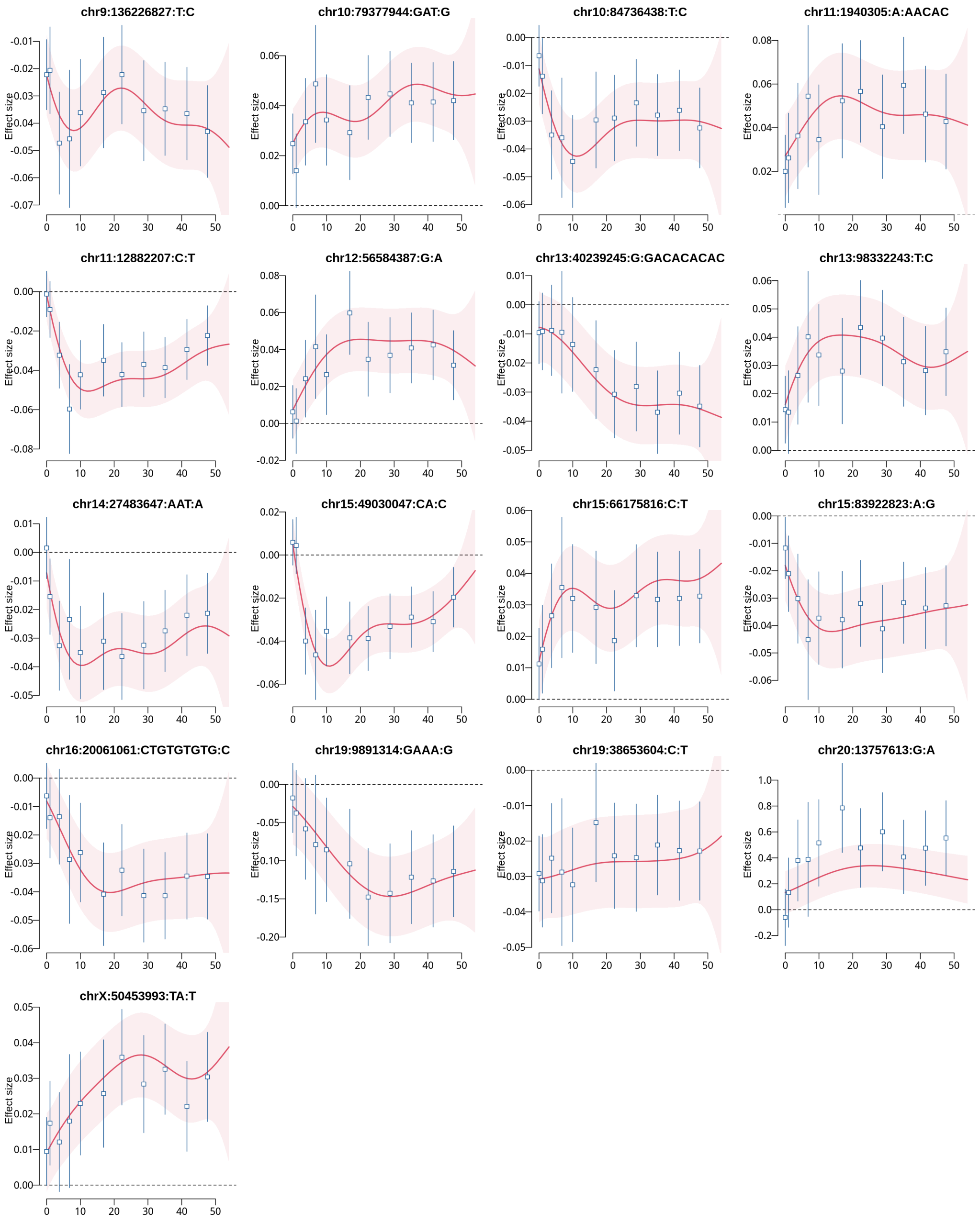


Figure 1 cont. The longitudinal QTL effect (red) and the cross-sectional QTL effects (blue) (detected only by longitudinal GWAS).

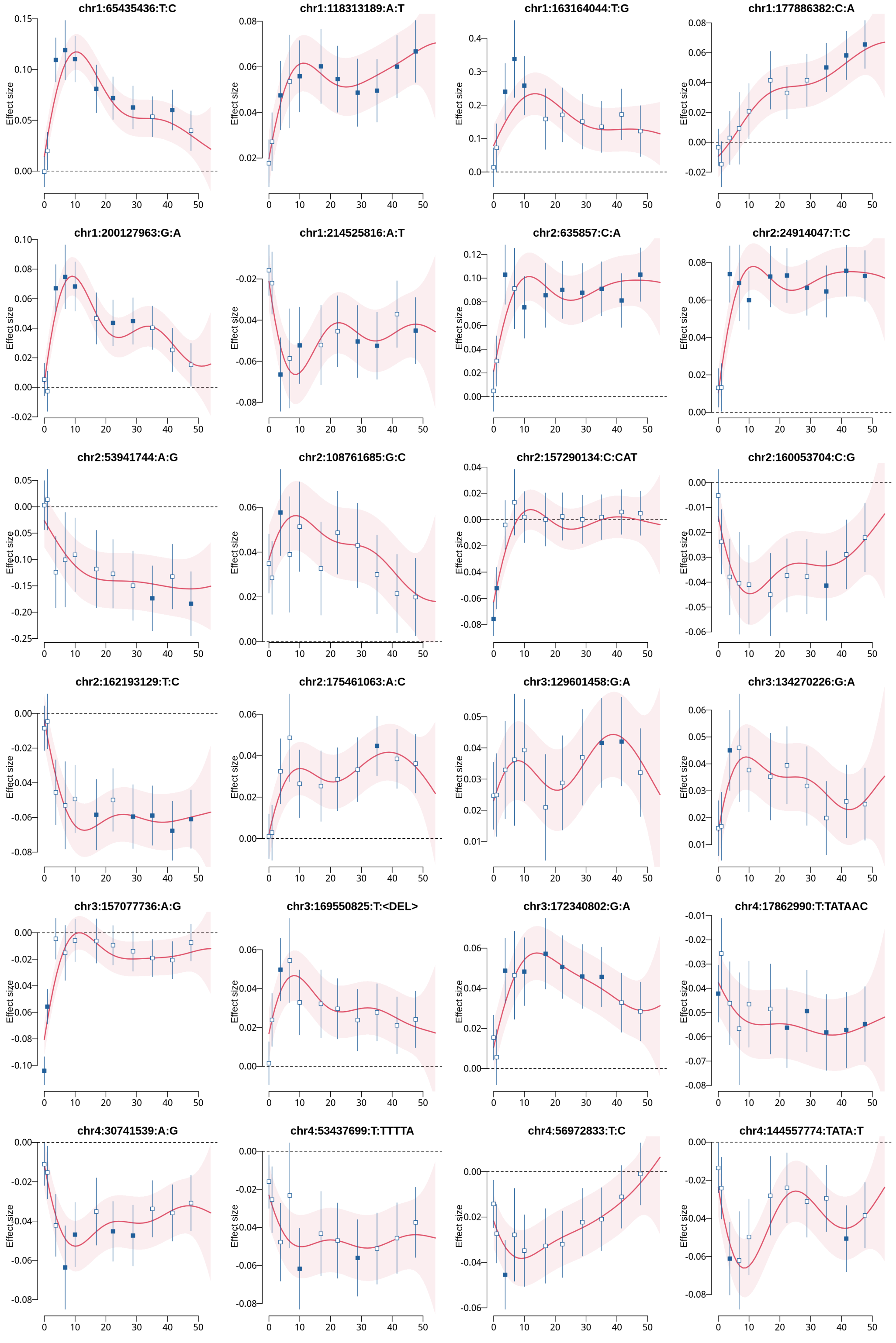


Figure 2. The longitudinal QTL effect (red) and the cross-sectional QTL effects (blue) (detected by both longitudinal and cross-sectional GWAS).

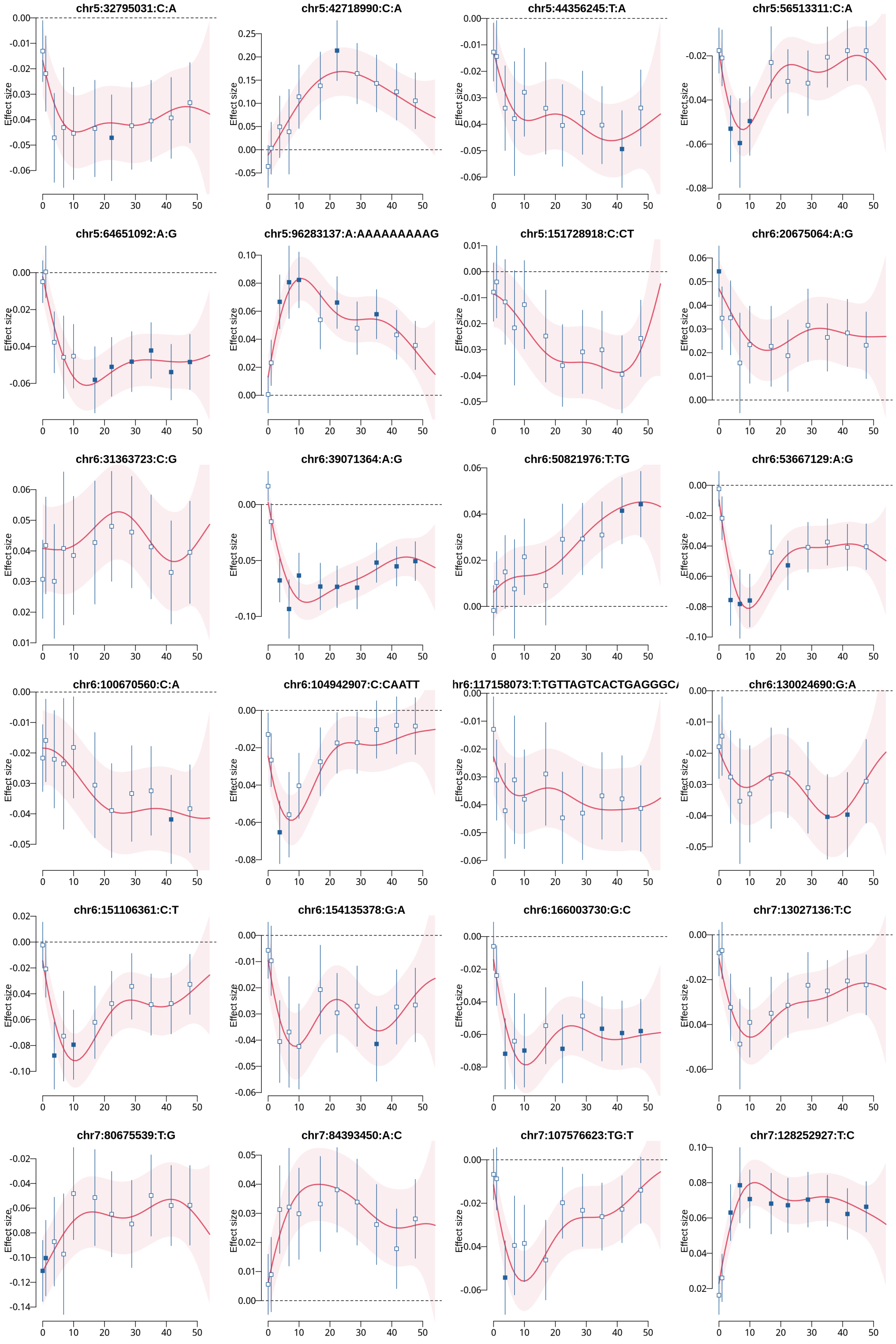


Figure 2 cont. The longitudinal QTL effect (red) and the cross-sectional QTL effects (blue) (detected by both longitudinal and cross-sectional GWAS).

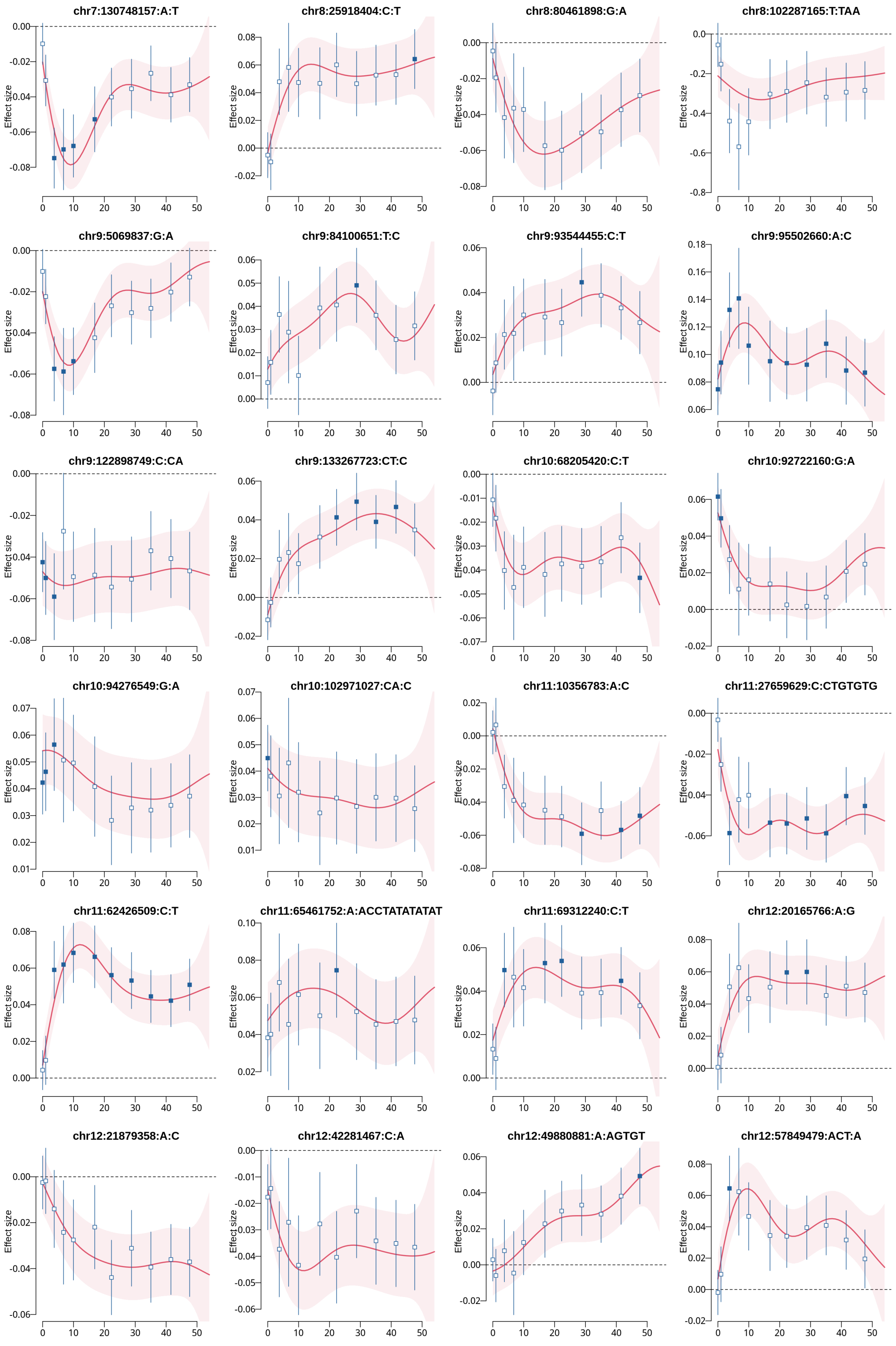


Figure 2 cont. The longitudinal QTL effect (red) and the cross-sectional QTL effects (blue) (detected by both longitudinal and cross-sectional GWAS).

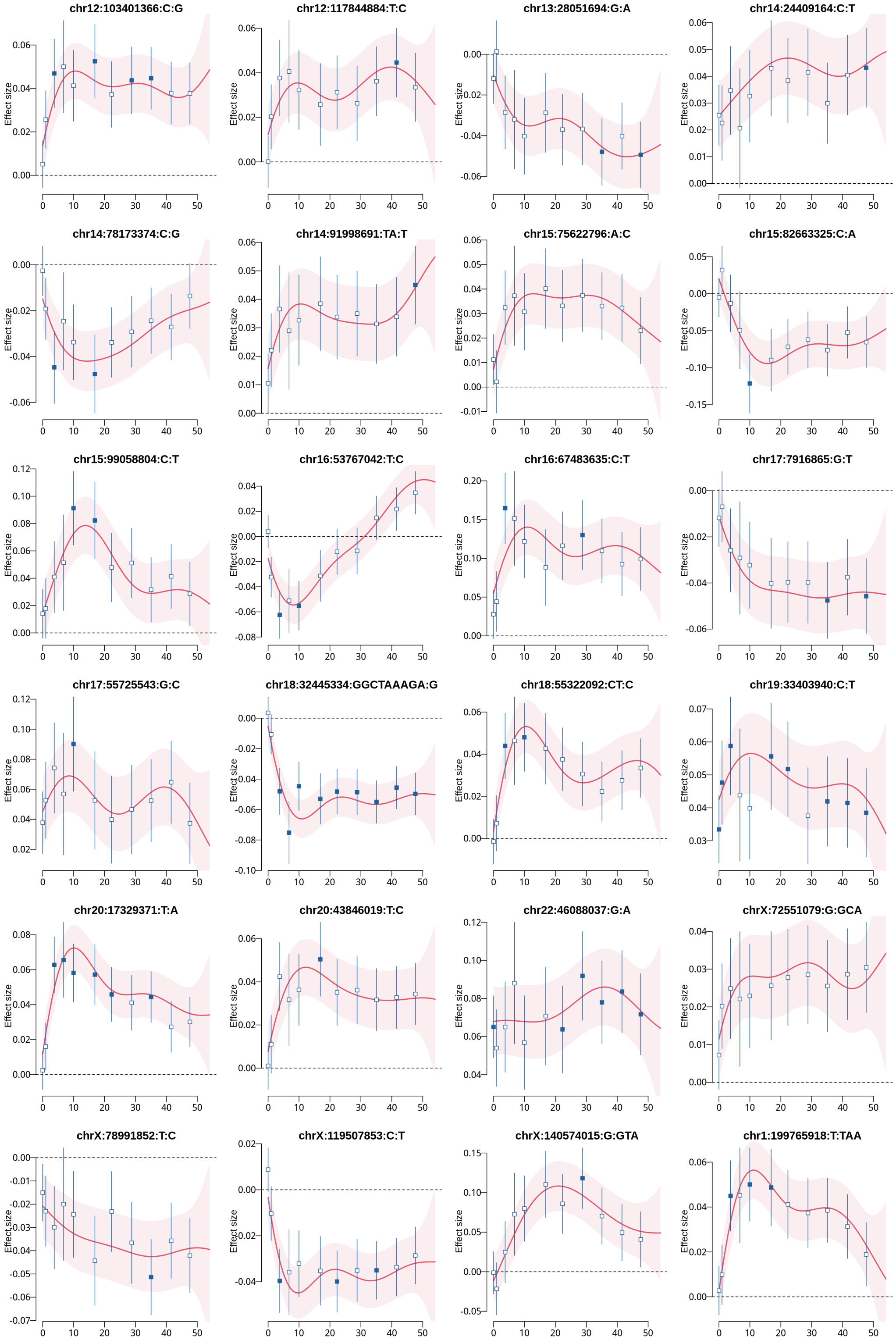


Figure 2 cont. The longitudinal QTL effect (red) and the cross-sectional QTL effects (blue) (detected by both longitudinal and cross-sectional GWAS).

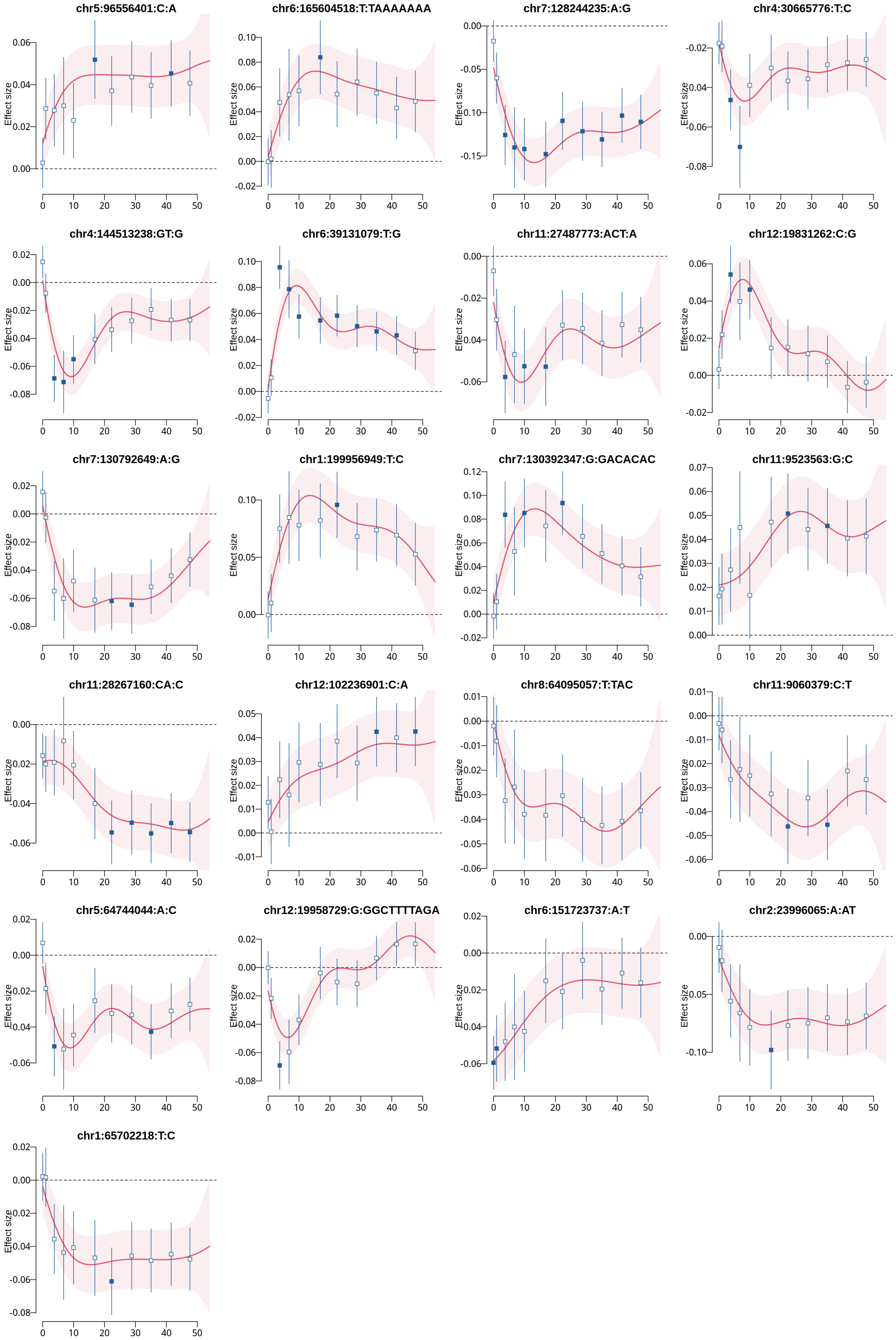


Figure 2 cont. The longitudinal QTL effect (red) and the cross-sectional QTL effects (blue) (detected by both longitudinal and cross-sectional GWAS).

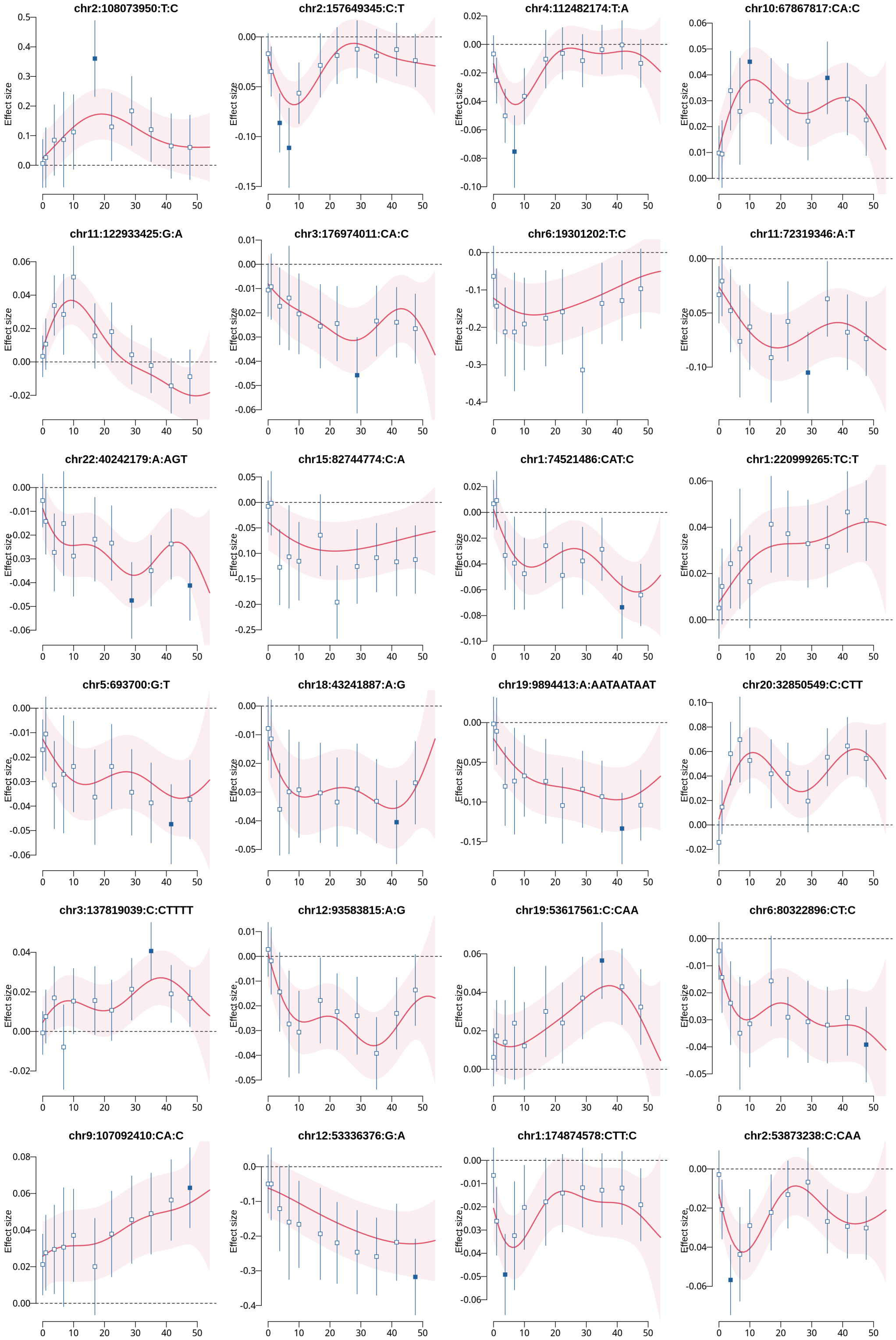


Figure 3. The longitudinal QTL effect (red) and the cross-sectional QTL effects (blue) (detected only by cross-sectional GWAS).

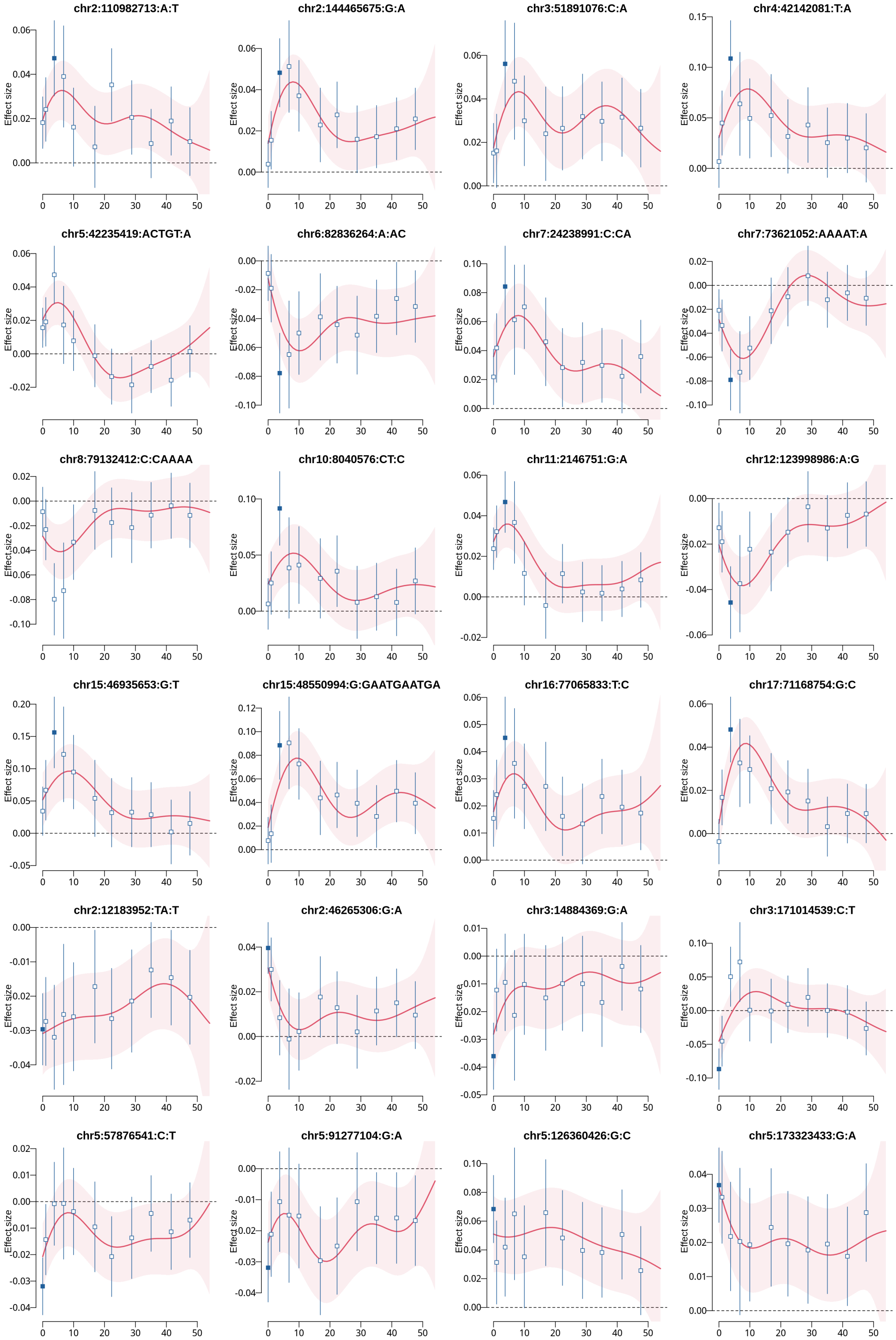


Figure 3 cont. The longitudinal QTL effect (red) and the cross-sectional QTL effects (blue) (detected only by cross-sectional GWAS).

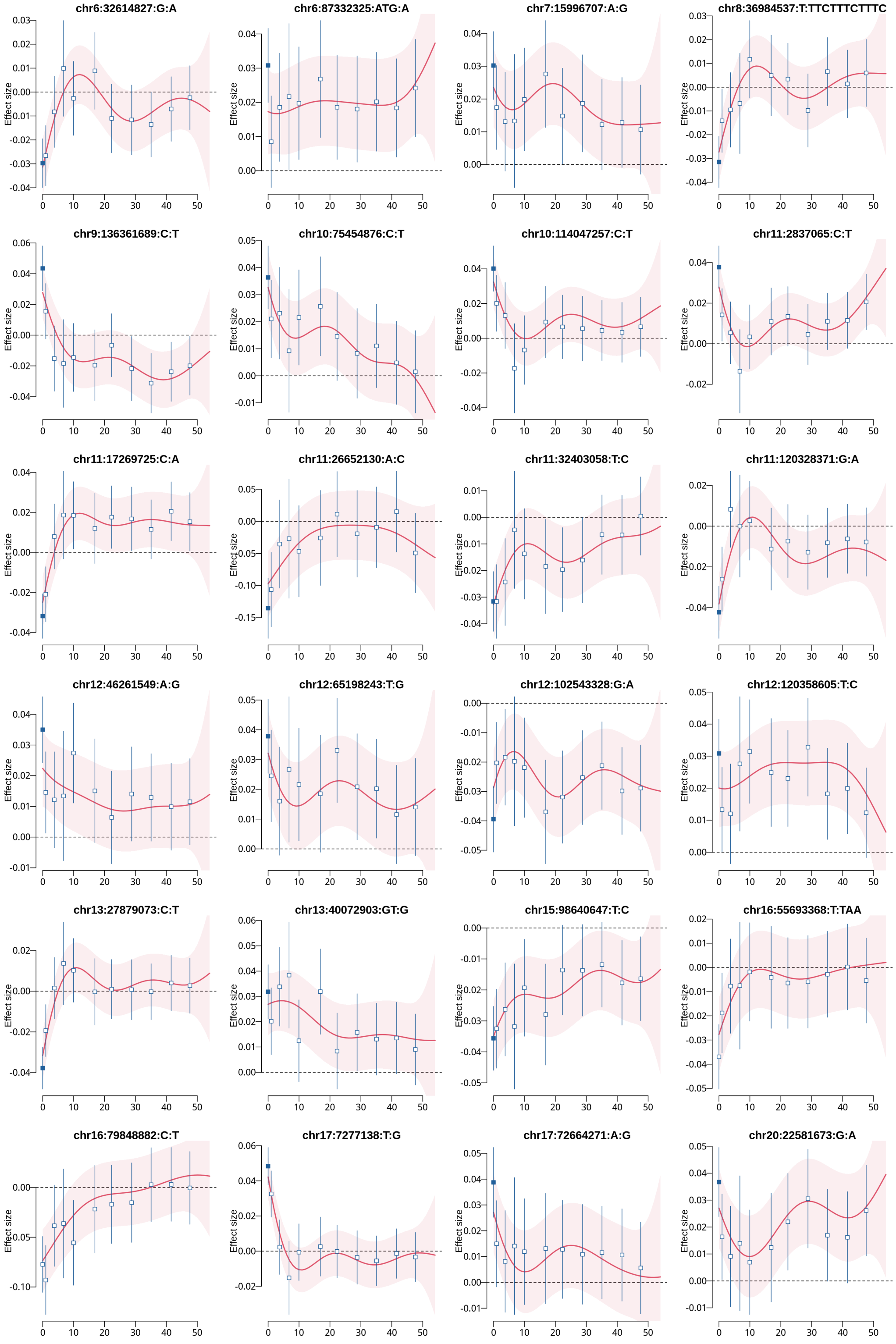


Figure 3 cont. The longitudinal QTL effect (red) and the cross-sectional QTL effects (blue) (detected only by cross-sectional GWAS).

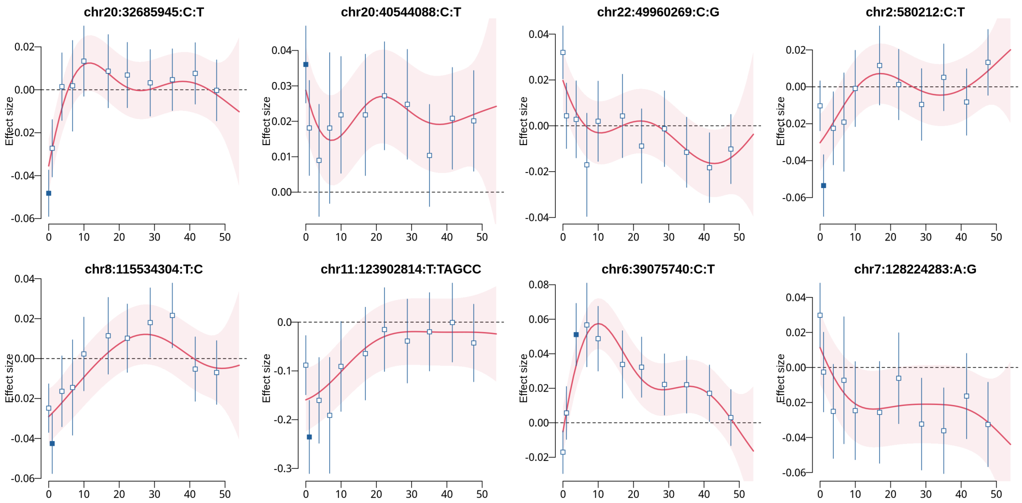


Figure 3 cont. The longitudinal QTL effect (red) and the cross-sectional QTL effects (blue) (detected only by cross-sectional GWAS).

### 2 Appendices

#### 2.1 Derivation of $Z$ scores for the simple linear regression with overlapping samples

Let us denote a response $y_i$ and a covariate $x_i$ for the individual $i$ ($i = 1, \dots, N$). Consider a simple linear regression model

$$y_i = \beta x_i + \varepsilon_i, \qquad \varepsilon_i \sim \mathcal{N}(0, \sigma^2).$$

Using all $N$ samples, the estimator of $\beta_1$ will be

$$\hat{\beta}_1^{(N)} = \beta_1 + \frac{\sum_i (x_i - \bar{x})\varepsilon_i}{\sum_i (x_i - \bar{x})^2}.$$

Using a downsampled subset $S$ of size $M = \rho^2 N$, the estimator will be

$$\hat{\beta}_1^{(M)} = \beta_1 + \frac{\sum_{i \in S} (x_i - \bar{x}_S)\varepsilon_i}{\sum_{i \in S} (x_i - \bar{x}_S)^2}.$$

The corresponding standard errors are then given by

$$\mathrm{SE}(\hat{\beta}_1^{(N)}) = \frac{\sigma}{\sqrt{\sum_i (x_i - \bar{x})^2}}, \qquad \mathrm{SE}(\hat{\beta}_1^{(M)}) = \frac{\sigma}{\sqrt{\sum_{i \in S} (x_i - \bar{x}_S)^2}}.$$

Hence, the *Z-scores* (standardized estimators) are defined by

$$Z^{(N)} = \frac{\hat{\beta}_1^{(N)}}{\mathrm{SE}(\hat{\beta}_1^{(N)})} = \frac{1}{\sigma} \sum_i w_i \varepsilon_i + \frac{\beta_1}{\mathrm{SE}(\hat{\beta}_1^{(N)})}, \qquad Z^{(M)} = \frac{\hat{\beta}_1^{(M)}}{\mathrm{SE}(\hat{\beta}_1^{(M)})} = \frac{1}{\sigma} \sum_i v_i \varepsilon_i + \frac{\beta_1}{\mathrm{SE}(\hat{\beta}_1^{(M)})},$$

where

$$w_i = \frac{x_i - \bar{x}}{\sqrt{\sum_j (x_j - \bar{x})^2}}, \qquad v_i = \begin{cases} \dfrac{x_i - \bar{x}_S}{\sqrt{\sum_{j \in S} (x_j - \bar{x}_S)^2}}, & i \in S, \\ 0, & i \notin S. \end{cases}$$

Since both are linear functions of the same Gaussian noise, we have

$$\mathrm{Cor}(Z^{(n)}, Z^{(m)}) = \sum_i w_i v_i.$$

Here we not that

$$\mathbb{E}[Z^{(N)}] = \frac{\beta_1}{\mathrm{SE}(\hat{\beta}_1^{(N)})}, \qquad \mathbb{E}[Z^{(M)}] = \frac{\beta_1}{\mathrm{SE}(\hat{\beta}_1^{(M)})},$$

and

$$\mathrm{Var}[Z^{(N)}] = \sum_i w_i^2 = 1, \qquad \mathrm{Var}[Z^{(M)}] = \sum_i v_i^2 = 1.$$

We then assume $x_i$ are i.i.d. with $\mathbb{E}[x_i] = 0$ and $\mathrm{Var}(x_i) = \tau^2$. When a random fraction $\rho^2 = M/N$ of samples is retained, centering effects are negligible, and we approximate

$$w_i \approx \frac{x_i}{\sqrt{\sum_j x_j^2}}, \qquad v_i \approx \frac{x_i I_i}{\sqrt{\sum_{j \in S} x_j^2}},$$

where $I_i = 1$ if $i \in S$, otherwise 0. Taking expectations, we have

$$\mathbb{E}[\mathrm{Cor}(Z^{(N)}, Z^{(M)})] \simeq \mathbb{E}\left[ \frac{\sum_i x_i^2 I_i}{\sqrt{(\sum_i x_i^2)(\sum_i x_i^2 I_i)}} \right].$$

Since $\mathbb{E}[I_i] = M/N = \rho^2$, we finally have

$$\mathbb{E}[\mathrm{Cor}(Z^{(N)}, Z^{(M)})] \approx \frac{\rho^2 \sum_i x_i^2}{\sqrt{(\sum_i x_i^2)(\rho^2 \sum_i x_i^2)}} = \rho.$$

Likewise

$$\mathbb{E}_x \left[ \frac{\mathbb{E}_\varepsilon[Z^{(M)}]}{\mathbb{E}_\varepsilon[Z^{(N)}]} \right] = \mathbb{E}_x \left[ \frac{\mathrm{SE}(\hat{\beta}_1^{(N)})}{\mathrm{SE}(\hat{\beta}_1^{(M)})} \right] \simeq \mathbb{E}_x \left[ \sqrt{\frac{\sum_i x_i^2 I_i}{\sum_i x_i^2}} \right] \approx \sqrt{\frac{\rho^2 \sum_i x_i^2}{\sum_i x_i^2}} = \rho.$$

#### 2.2 Derivation of noncentral $\chi^2$ distribution

We want to find the conditional distribution of $Z_2^2$ given $Z_1^2 = 1 + \mu^2$, such that

$$(Z_1, Z_2) \sim N\left( \begin{pmatrix} \mu \\ \mu\rho \end{pmatrix}, \begin{pmatrix} 1 & \rho \\ \rho & 1 \end{pmatrix} \right).$$

From the properties of the bivariate normal distribution:

$$Z_2 \mid Z_1 = z_1 \sim N(\rho z_1,\, 1 - \rho^2),$$

the squared variable follows a scaled noncentral chi-square distribution with one degree of freedom:

$$Z_2^2 \mid Z_1 = z_1 \;\sim\; (1 - \rho^2)\, \chi_1'^2 \left( \frac{\rho^2 z_1^2}{1 - \rho^2} \right),$$

where $\chi_1'^2(\lambda)$ denotes the noncentral chi-square distribution with 1 degree of freedom and noncentrality parameter $\lambda$. Since the conditional distribution depends only on $z_1^2$, the sign of $z_1$ is irrelevant. Therefore we have

$$Z_2^2 \mid (Z_1^2 = 1 + \mu^2) \;\sim\; (1 - \rho^2)\, \chi_1'^2 \left( \frac{\rho^2 (1 + \mu^2)}{1 - \rho^2} \right).$$

Note that, the conditional expectation is

$$\mathbb{E}[Z_2^2 \mid Z_1^2 = x] = (1 - \rho^2) + \rho^2 x,$$

hence, for $x = 1 + \mu^2$, we have

$$\mathbb{E}[Z_2^2 \mid Z_1^2 = 1 + \mu^2] = (1 - \rho^2) + \rho^2 (1 + \mu^2).$$

#### 2.3 Estimation of polynomial-model associations missed by GP screening

We estimated the expected number of genome-wide significant associations under the polynomial model that could have been missed because polynomial-model testing was restricted to variants with $P_G < p_{\text{screen}}$, where $p_{\text{screen}} = 10^{-4}$. For variants tested by both methods, the GP and polynomial-model $P$-values were converted to one-degree-of-freedom chi-square statistics,

$$T_{G,i} = F_{\chi_1^2}^{-1}(1 - P_{G,i}), \qquad T_{P,i} = F_{\chi_1^2}^{-1}(1 - P_{P,i}),$$

and their empirical relationship was estimated as

$$T_{P,i} = a + b T_{G,i} + \varepsilon_i.$$

Let $r_T = \mathrm{Cor}(T_G, T_P)$. Under the latent representation $T_G = Z_G^2$ and $T_P = Z_P^2$, the latent normal correlation was approximated by

$$\rho = \sqrt{\max(r_T, 0)},$$

using $\mathrm{Cor}(Z_G^2, Z_P^2) = \rho^2$ for correlated standard normal variables under the central $\chi_1^2$ model.

For each variant excluded by GP screening, the GP noncentrality parameter was estimated by moment matching as

$$\widehat{\lambda}_{G,i} = \max(T_{G,i} - 1, 0)\,,$$

and the corresponding polynomial-model noncentrality was approximated using the fitted slope as

$$\widehat{\lambda}_{P,i} = \max\Big(b\widehat{\lambda}_{G,i}, 0\Big)\,.$$

We then assumed

$$\begin{pmatrix} X_{G,i} \\ X_{P,i} \end{pmatrix} \sim \mathcal{N}_2\left[\begin{pmatrix} 0 \\ 0 \end{pmatrix}, \begin{pmatrix} 1 & \rho \\ \rho & 1 \end{pmatrix}\right],$$

with

$$T_{G,i} = \left(X_{G,i} + \sqrt{\widehat{\lambda}_{G,i}}\right)^2, \qquad T_{P,i} = \left(X_{P,i} + \sqrt{\widehat{\lambda}_{P,i}}\right)^2.$$

Let

$$c_G = F_{\chi_1^2}^{-1}(1 - p_{\text{screen}})\,, \qquad c_P = F_{\chi_1^2}^{-1}(1 - p_{\text{sig}})\,,$$

where $p_{\text{sig}} = 5 \times 10^{-8}$. Define

$$\ell_{G,i} = -\sqrt{c_G} - \sqrt{\widehat{\lambda}_{G,i}}, \qquad u_{G,i} = \sqrt{c_G} - \sqrt{\widehat{\lambda}_{G,i}},$$

and

$$\ell_{P,i} = -\sqrt{c_P} - \sqrt{\widehat{\lambda}_{P,i}}, \qquad u_{P,i} = \sqrt{c_P} - \sqrt{\widehat{\lambda}_{P,i}}.$$

Then

$$A_i = \Pr(T_{G,i} \le c_G) = \Phi(\nu_{G,i}) - \Phi(\ell_{G,i})\,,$$

where $\Phi$ is the standard normal cumulative distribution function. If $\Phi_2(x, y; \rho)$ denotes the standard bivariate normal cumulative distribution function with correlation $\rho$, the probability that both statistics are below their respective thresholds is

$$\begin{aligned} B_i = {} & \Pr(T_{G,i} \le c_G,\ T_{P,i} \le c_P) \\ = {} & \Phi_2(\nu_{G,i}, \nu_{P,i}; \rho) - \Phi_2(\ell_{G,i}, \nu_{P,i}; \rho) \\ & - \Phi_2(\nu_{G,i}, \ell_{P,i}; \rho) + \Phi_2(\ell_{G,i}, \ell_{P,i}; \rho)\,. \end{aligned}$$

The conditional probability that a variant reaches genome-wide significance under the polynomial model despite failing the GP screen is therefore

$$q_i = \Pr(T_{P,i} > c_P \mid T_{G,i} \le c_G) = \frac{A_i - B_i}{A_i}.$$

The expected number of missed variants is

$$\mathrm{E}(N_{\text{miss}}) = \sum_{i=1}^{N} q_i.$$

##### 2.4 Cross-trait LD Score Regression and Environmental Correlation

In bivariate LD Score regression, the expectation of the product of $z$-scores from two GWASs (trait 1 and trait 2) at SNP $j$ is modeled as

$$E[z_{1j}z_{2j}] = \frac{\sqrt{N_1 N_2}}{M} \rho_g \, l_j + \text{Intercept},$$

where $z_{1j}, z_{2j}$ are $z$-statistics for SNP $j$ in the two GWASs, $l_j$ is the LD score of SNP $j$, $M$ is the number of SNPs, $N_1, N_2$ are sample sizes, $\rho_g$ is the genetic covariance between the two traits, and the "Intercept" captures the non-genetic covariance (sample overlap, population structure, etc.). If a fraction of the samples is shared between the two GWASs, the intercept term can be expressed as

$$\text{Intercept} = \frac{N_{\text{overlap}}}{\sqrt{N_1 N_2}} \rho_p,$$

where $N_{\text{overlap}}$ is the number of overlapping individuals, and $\rho_p = \text{Cov}(Y_1, Y_2)$ is the phenotypic covariance (i.e., phenotypic correlation when standardized). Thus, the full model becomes

$$E[z_{1j}z_{2j}] = \frac{\sqrt{N_1 N_2}}{M} r_g \sqrt{h_1^2 h_2^2} \, l_j + r_p \frac{N_{\text{overlap}}}{\sqrt{N_1 N_2}}.$$

Here, the first term represents the contribution from shared genetic effects (the slope), and the second term (Intercept) represents the contribution from shared environment or sample overlap.

Let each standardized phenotype be decomposed as

$$Y_i = G_i + E_i, \quad \text{Var}(G_i) = h_i^2, \quad \text{Var}(E_i) = 1 - h_i^2,$$

for the trait $i = 1$ and 2, the phenotypic covariance can be written as

$$\text{Cov}(Y_1, Y_2) = \text{Cov}(G_1, G_2) + \text{Cov}(E_1, E_2).$$

Dividing by standard deviations gives

$$r_p = r_g \sqrt{h_1^2 h_2^2} + r_e \sqrt{(1 - h_1^2)(1 - h_2^2)}.$$

Thus, rearranging the above equation yields the environmental correlation as follows:

$$r_e = \frac{r_p - r_g \sqrt{h_1^2 h_2^2}}{\sqrt{(1 - h_1^2)(1 - h_2^2)}}.$$

Note here that, the intercept of cross-trait LDSC is given by `gcov_int` in the output file.

### References


*Cuomo Anna S E, Heinen Tobias, Vagiaki Danai, Horta Danilo, Marioni John C, Stegle Oliver*. CellRegMap: a statistical framework for mapping context-specific regulatory variants using scRNA-seq // Mol. Syst. Biol. VIII 2022. 18, 8. e10663.

*Kumasaka Natsuhiko, Rostom Raghd, Huang Ni, Polanski Krzysztof, Meyer Kerstin B, Patel Sharad, Boyd Rachel, Gomez Celine, Barnett Sam N, Panousis Nikolaos I, Schwartzentruber Jeremy, Ghoussaini Maya, Lyons Paul A, Calero-Nieto Fernando J, Göttgens Berthold, Barnes Josephine L, Worlock Kaylee B, Yoshida Masahiro, Nikolić Marko Z, Stephenson Emily, Reynolds Gary, Haniffa Muzlifah, Marioni John C, Stegle Oliver, Hagai Tzachi, Teichmann Sarah A*. Mapping interindividual dynamics of innate immune response at single-cell resolution // Nat. Genet. VI 2023. 55, 6. 1066–1075.

*Titsias M*. Variational learning of inducing variables in sparse Gaussian processes // Artificial Intelligence and Statistics. 2009.
