## Supplementary Notes 3 for "Genome-wide association study on longitudinal and cross-sectional traits of child health and development in a Japanese population"

### Norwegian Mother, Father and Child Cohort Study (MoBa) - cohort description

#### Ethics declarations

Informed consent was obtained from all study participants. The administrative board of the Norwegian Mother, Father and Child Cohort Study (MoBa) led by the Norwegian Institute of Public
Health (NIPH) approved the study protocol. The establishment of MoBa and initial data collection was based on a license from the Norwegian Data Protection Agency and approval from The Regional Committee for Medical Research Ethics. The MoBa cohort is currently regulated by the Norwegian Health Registry Act. The study was approved by The Regional Committee for Medical Research Ethics (#2012/67).


#### Acknowledgments

We thank the Norwegian Institute of Public Health (NIPH) for establishing and maintaining MoBa. We are grateful to all the families in Norway who are taking part in the ongoing MoBa cohort study.

Genotying in MoBa was part of the HARVEST collaboration, supported by the Research Council of Norway (#229624). We also thank the NORMENT Centre for providing genotype data, funded by the Research Council of Norway (#223273), South East Norway Health Authorities and Stiftelsen Kristian Gerhard Jebsen, and in collaboration with deCODE Genetics. We further thank the Center for Diabetes Research, the University of Bergen for providing genotype data and performing initial quality control and imputation of the data funded by the ERC AdG project SELECTionPREDISPOSED, Stiftelsen Kristian Gerhard Jebsen, Trond Mohn Foundation, the Research Council of Norway, the Novo Nordisk Foundation, the University of Bergen, and the Western Norway Health Authorities. We thank the MoBaPsychGen team, led by Elizabeth Corfield, for establishing and sharing quality controlled genotype data, supported by funding from the South-Eastern Norway Regional Health Authority (#2021045; #2020022; #2022083; 2018058). We thank the many people in Norway and abroad who contributed to the development of this resource.

Quality control, phasing, imputation, and harmonization of genotypes from the MoBa cohort were performed using digital labs in HUNT Cloud at the Norwegian University of Science and Technology (NTNU), Trondheim, Norway. We are grateful for the support from the HUNT Cloud community.

#### Data Availability

Data from the Norwegian Mother, Father and Child Cohort Study used in this study are managed by the Norwegian Institute of public health and can be made available to researchers, provided approval from the Regional Committees for Medical and Health Research Ethics (REC), compliance with the EU General Data Protection Regulation (GDPR) and approval from the data owners. The consent given by the participants does not open for storage of data on an individual level in repositories or journals. Researchers who want access to data sets for replication should apply through helsedata.no. Access to data sets requires approval from The Regional Committee for Medical and Health Research Ethics in Norway and an agreement with MoBa. The sample sizes for EU_core_EU are given by:

| Timepoint | raw_bmi n | raw_bmi + covariates n |
|---|---|---|
| 6w | 46125 | 45881 |
| 3m | 64339 | 64041 |
| 6m | 65455 | 65151 |
| 8m | 55053 | 54786 |
| 1y | 56882 | 56607 |
| 16m | 42663 | 42451 |
| 2y | 42627 | 42417 |
| 3y | 43495 | 43281 |
| 5y | 34942 | 34775 |
